# Influenza forecasting for the French regions by using EHR, web and climatic data sources with an ensemble approach ARGONet

**DOI:** 10.1101/19009795

**Authors:** Canelle Poirier, Yulin Hswen, Guillaume Bouzillé, Marc Cuggia, Audrey Lavenu, John S Brownstein, Thomas Brewer, Mauricio Santillana

## Abstract

Effective and timely disease surveillance systems have the potential to help public health officials design interventions to mitigate the effects of disease outbreaks. Currently, healthcare-based disease monitoring systems in France offer influenza activity information that lags real-time by 1 to 3 weeks. This temporal data gap introduces uncertainty that prevents public health officials from having a timely perspective on the population-level disease activity. Here, we present a machine-learning modeling approach that produces real-time estimates and short-term forecasts of influenza activity for the 12 continental regions of France by leveraging multiple disparate data sources that include, Google search activity, real-time and local weather information, flu-related Twitter micro-blogs, electronic health records data, and historical disease activity synchronicities across regions. Our results show that all data sources contribute to improving influenza surveillance and that machine-learning ensembles that combine all data sources lead to accurate and timely predictions.

**Author summary:** The role of public health is to protect the health of populations by providing the right intervention to the right population at the right time. In France and all around the world, Influenza is a major public health problem. Traditional surveillance systems produce estimates of influenza-like illness (ILI) incidence rates, but with one-to three-week delay. Accurate real-time monitoring systems of influenza outbreaks could be useful for public health decisions. By combining different data sources and different statistical models, we propose an accurate and timely forecasting platform to track the flu in France at a spatial resolution that, to our knowledge, has not been explored before.

## Introduction

Influenza is a major public health problem causing up to 5 million severe cases and 500,000 deaths per year worldwide [1–3]. In France alone, the epidemic of 2018-2019 caused 9,500 deaths. During epidemic peaks, large increases of visits to general practitioners and to emergency departments are observed and often lead to disruptions to healthcare delivery and thus increase the risk of undesirable outcomes in patients with influenza infections. To reduce the impact of influenza outbreaks in the population and to better design timely public health interventions, surveillance systems that produce accurate real-time and short-term forecasts of disease activity may prove to be instrumental.

In France, an important influenza monitoring system was implemented by the Sentinelles network in 1984 [4, 5]. This system centralizes information obtained from a group of volunteer (1314 in 2018) general practitioners and (116 in 2018) pediatricians that each week report the proportion of patients with Influenza-Like-Illness (ILI, any acute respiratory infection with fever ≥ 38 °C, cough and onset within the last 10 days) seeking medical attention. Data collection, processing, aggregation and distribution processes of this information, at the national and regional levels, introduce up to three weeks delays in the availability of flu activity information. This temporal data gap prevents public health officials from having the most up-to-date epidemiological information, and thus leads to the design of interventions that do not take into consideration recent changes in disease activity [2, 6]. For example, if estimates were available in real-time, information campaigns and vaccination prevention could be deployed earlier and and could lead to grater impact. Additionally, healthcare facilities could be better prepared to respond to unexpected increases in the flux of high-risk patient during time periods of increased disease activity.

With the motivation to alleviate this time delay, mathematical modeling and machine learning approaches have been proposed to produce disease estimates in real time and ahead of healthcare-based surveillance systems in multiple nations around the world. Most of these studies have been designed and tested in developed nations, such as the United States and France, where information on disease outbreaks has been collected historically for decades [2]. Numerous research studies have been conducted on the use of traditional statistical methods, like temporal series or compartmental methods, as well as the inclusion of disparate data sources such as meteorological or demographic data to track flu activity, as discussed in Nsoesie et al. 2014 and Yang and Shamman 2014 [7, 8]. And in recent years, multiple more studies have emerged exploring the use of Internet-based data sources that capture aspects of human behavior and environmental factors to track the spread of diseases. With over 3.2 billion web users, data flows from the internet are huge and of all types. Some studies have used data from Google [2, 3, 9–11], Twitter [12–14] or Wikipedia [15–18] to monitor flu specifically.

One of the first and most prominent studies on the use of internet data for monitoring influenza epidemics is Google Flu Trends (GFT) [19]. This web-based platform, created in 2009 and designed and deployed by Google, used the volume of selected Google search terms to estimate ILI activity in real time. GFT led to multiple prediction errors during the 2009 H1N1 Flu Pandemic (due to changes in people’s search behaviour as a result of the exceptional nature of the pandemic) and later produced large overestimations during the 2012-2013 US flu season (due to the announcement of a pandemic that finally did not appear). These events led to eventual discontinuation of this disease monitoring platform [20]. Since then, multiple research teams have proposed improved methodologies that are capable of extracting information more efficiently from flu-related Google searches and produce improved flu estimates. Among these methods, the work of Shihao Yang et al. [2] explored a penalized regression methodology that combines historical flu activity with Google search activity dynamically, called ARGO, to better predict flu.

Additional data sources have been explored to monitor flu activity such as clinicians’ searches, electronic health records (EHR), crowd-sourced flu monitoring apps [21–23]. Among these, electronic health records have been shown to track flu accurately and timely in the US and France [6, 24–26]. Specifically, in United States, Santillana et al. [6] showed that a model leveraging EHR data and a machine learning algorithms was capable to monitor flu activity in multiple spatial resolutions that included the regional level. In France, Poirier et al. [24] similarly showed multiple statistical models that incorporate EHR and Internet-search data, can yield accurate ILI incidence rates in real time at the national level.

In early 2019, Fred S. Lu et al. [27] extended the ARGO methodology to accurately track flu activity in multiple states of the United States. In their approach, they included Google search data, EHRs and historical flu trends. They developed also a spatial network approach, called Net, to capture the synchronicity observed historically in flu activity between each states. Finally, by dynamically combining estimates from ARGO and Net, they showed that an ensemble approach, named ARGONet, led to improved results.

### Our contribution

In this study, we propose a forecasting platform that combines multiple data sources and statistical models to track flu activity in France at a spatial resolution that, to our knowledge, has not been explored before. Our forecasting platform produces accurate region-specific real-time and short-term flu activity forecasts for the 12 continental French regions, by leveraging national-level flu-related Google searches, electronic health records data, Twitter data, and region-specific climate data. Additionally, historical synchronicities across regions are captured with a Network model. A machine learning ensemble approach is proposed to improve predictions by dynamically combining estimates from these two distinct approaches. Near real-time estimates as well as one- and two-week ahead forecasts are presented.

## Materials and methods

### Data sources

#### Sentinelles network data

We obtained weekly ILI incidence rates (per 100000 inhabitants) for the French regions (12) from the French Sentinelles network (websenti.u707.jussieu.fr/sentiweb). We retrieved these data in August 2018 from 05 January 2004 to 13 March 2017. We considered these data as the gold standard and as our task for our prediction models.

#### Google Data

We obtained the frequency per week of the 100 most correlated internet queries (if correlation ≥ 0.60) by French users from Google Correlate (https://www.google.com/trends/correlate. Because our prediction period spans 05 January 2015 to 20 February 2017, we utilized the ILI signal for each French region, from January 2004 to December 2014 to obtain the most highly correlated search terms using the tool Google Correlate. In this way, we obtained different search terms for each individual region. The signals obtained correspond to queries performed by French users at the national level. We retrieved Google Correlate data in August 2018 for the period going from 05 January 2004 to 13 March 2017.

#### Electronic Health Record Data

We retrieved EHR data from the clinical data warehouse (CDW) of Rennes University Hospital (France),This CDW, called eHOP, integrates structured (laboratory test results, prescriptions, ICD-10 diagnoses) and unstructured (discharge letter, pathology reports, operative reports) patients’ data. It includes data from 1.2 million inpatients and outpatients and 45 million documents that correspond to 510 million structured elements. eHOP consists of a powerful search engine system that can identify patients with specific criteria by querying unstructured data with keywords, or structured data with querying codes based on terminologies.

The first approach to obtain eHOP data connected with ILI was to perform different manual queries to retrieve patients who had at least one document in their EHR that matched the following search criteria: (1) Queries directly connected with flu or ILI with the keywords “flu” or “ILI”; (2) Queries connected with flu symptoms with the keywords “fever”, “pyrexia”, “body aches” or “muscular pain”; (3) Queries connected with flu drugs with the keyword “Tamiflu”; (4) Queries with the ICD-10 terminology; (5) Queries connected with flu tests, positive or negative results.

In total, we performed 34 manual queries. For each query, the eHOP search engine returned all documents containing the chosen keywords (often, several documents for one patient and one stay). For query aggregation, we kept the oldest document for one patient and one stay and then calculated, for each week, the number of stays with at least one document mentioning the keyword contained in the query.

From the CDW eHOP, we built a database containing the time series constructed from the structured data. In all, we have 1 335 347 time series. As Google Correlate, the Pearson correlation between each signal of each region and the time series from the database was calculated. In this way, for each region, the second approach was to retrieve the 100 most correlated signals to ILI signal. Because our test period is from 05 January 2015 to 20 February 2017, we calculated the correlation between January 2004 and December 2014.

As a result, for each region, we obtained 134 variables from the CDW eHOP where there are at least 34 variables common to all regions (manual queries). We retrieved retrospective data in August 2018 for the period going from 03 January 2005 to 13 March 2017. This study was approved by the local Ethics Committee of Rennes Academic Hospital (approval number 16.69).

#### Weather Data

We obtained region-specific weather data from the French climatological website Info Climat (https://www.infoclimat.fr). It has been shown in several studies that humidity is correlated with the spread of influenza. [28]. In the absence of humidity data on the Climat website, we retrieved precipitation and temperatures data. This choice was made knowing that both variables, [29, 30] and temperature and precipitation can be used as a proxy for humidity since they are directly related by the Clausius–Clapeyron relation. [31] We obtained temperatures and precipitations per day for the largest city of each region, and calculated the weekly mean for both temperature and precipitation. We retrieved climatic data in August 2018 for the time period going from 07 January 2008 to 13 March 2017.

#### Twitter Data

Geotag tweets were extracted as the national scale for France from Boston Children’s Hospital Geotweet dataset with the following keywords pertaining to influenza (“grippe”, “grippé”, “syndrome grippal”, “fièvre”, “toux”, “congestion”, “malade”, “faiblesse”, “courbatures”, “tamiflu”, “la crève”). From there, we aggregated tweets to get weekly counts. In this way, we obtained 11 variables from Twitter. We retrieved Twitter data in December 2018 for the period going from 30 December 2013 to 13 March 2017.

### Statistical models

#### The ARGO model

The ARGO model is a regularized regression dynamically calibrated weekly using the LASSO method [32] to combine multiple external data sources with historical flu information. We performed the LASSO regression with the R package caret and the associated function fit with the method glmnet [33, 34]. We optimized the shrinkage parameter lambda via a 10-fold cross-validation. To test the stationarity and whiteness of residuals, we used Dickey Fuller’s and Box-Pierce’s tests available from the R packages tseries and stats [35]. The formulation of our model is :

- Real time estimates:

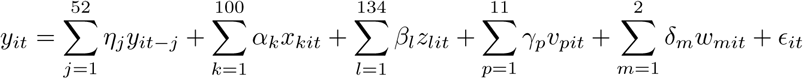

- One-week ahead forecast:

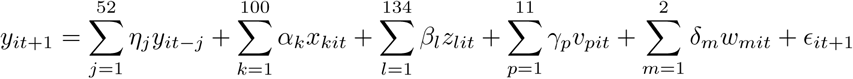

- Two-week ahead forecast:

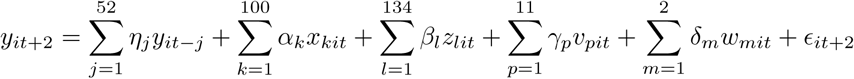

where *y*_*it*_ corresponding to the flu incidence rate at time *t* for the region 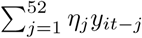 corresponding to the historical flu incidence rates for the region *i*, 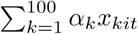 corresponding to Google data for the region *i*, 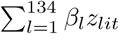 corresponding to hospital data for the region *i*, 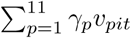 corresponding to Twitter data, 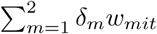 corresponding to climatic data for the region *i, ϵ*_*t*_ corresponding to residuals.

We applied this model for each region. The model was dynamically recalibrated every week by incorporating all data available. In this way, the size of our training dataset increases every week. We obtained estimates from January 2011 to March 2017.

#### The Net model

The Net model is a LASSO model dynamically calibrated weekly and using the relationship between the regions to know how synchronicity could improve forecasts. Indeed, Figure S1 (Heatmap of pairwise correlations between all regions) shows that the flu incidence rates of the different areas are correlated. For each region, we used historical data of all regions and estimates obtained with ARGO model for all regions expected the region to be predicted.

The formulation of our model is :

- Real time estimates:

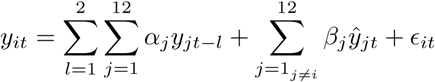

- One-week ahead forecast:

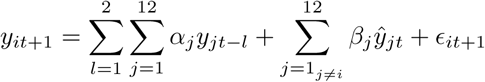

- Two-week ahead forecast:

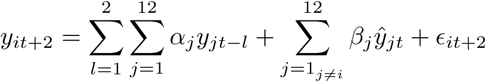

where *y*_*it*_ corresponding to the flu incidence rate at time *t* for the region *i*, 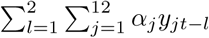 corresponding to two weeks of historical flu incidence rates for all regions, 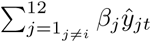 corresponding to ARGO predictions for all regions excepted the region *i* to be predicted and *ϵ*_*t*_ corresponding to residuals.

We applied this model for each region. We used a two years’ training dataset. We obtained estimates from January 2013 to March 2017.

#### The ARGONet model

The ARGONet model is an ensemble approach combining the predictive power of ARGO and Net models. For this model we tested three methods :

- The first is, ARGONet’s estimate is the ARGO estimate if ARGO model gives the lowest mean error in the previous K estimates compared to Net model. Otherwise, ARGONet’s estimate is the Net estimate. The value of K can be 1, 2, 3 or 4.
- The second is, ARGONet’s estimate is the mean between ARGO’s estimate and Net’s estimate.
- The third is, ARGONet’s estimate is the result of a linear regression between ARGO’s estimate and Net’s estimate. We trained the linear regression model on a period of two years.

#### The Baseline Autoregressive model

To assess the importance of external data sources, we built an autoregressive model of order 52 (AR(52)). We used the LASSO regression with the previous 52 weeks of ILI incidence rates to predict the current week and the two weeks after.

- Real time estimates:

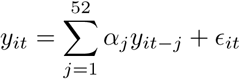

- One-week ahead forecast:

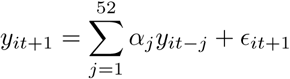

- Two-week ahead forecast:

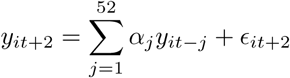

where *y*_*it*_ corresponding to the flu incidence rate at time *t* for the region *i*, 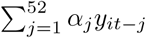 corresponding to the previous 52 weeks, *ϵ*_*t*_ corresponding to residuals.

We applied this model for each region. We used a six years’ training dataset. The model was dynamically recalibrated every week.

### Evaluation

Our test period consists on 115 weeks starting from January 2015 to March 2017.

#### Metrics

To assess the performance of the models, we compared estimates to the official incidence rates from the Sentinelles network by calculating two metrics : the mean squared error (MSE) and the Pearson correlation coefficient (PCC).

- 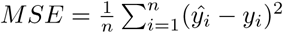
- 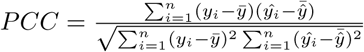

where *ŷ*_*i*_ is the predicted value for the week *i*, 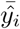 is the mean of predicted values, *y*_*i*_ the real value for the week *i*, 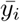 is the mean of real values.

We also estimated the relative efficiency of ARGONet model compared to the autoregressive model with 95% confidence interval (CI) by using a Bootstrap method. A relative efficiency, calculated by 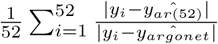 bigger than 1, suggests increased predictive power of ARGONet compared to the autoregressive model. The CI and relative efficiency have been computed based on 100 Bootstrap samples of length 52. The 52 weeks were randomly selected from estimates from January 2015 to February 2017.

#### Comparisons

First, we assessed the importance of adding external data sources by comparing :

- MSE and PCC of the autoregressive model and the ARGO model including historical data plus the 10 most correlated variables from hospital data and Google data. The individual contribution of hospital data and Google data has already been shown in a previous study [24]. But, we added in appendices, two comparisons: A comparison with the 10 most correlated variables from hospital data and a comparison with the 10 most correlated variables from Google data.
- MSE and PCC of the autoregressive model and the ARGO model including historical data plus climatic data.
- MSE and PCC of the autoregressive model and the ARGO model including historical data plus Twitter data.

Second, we compared the autoregressive model, ARGO model (including all the data sources), Net model and ARGONet model.

## Results

### Evaluation of Data Sources as Predictors

In order to assess the predictive value of each and all external data source, we compared ARGO models that incrementally included external data sources with a baseline autoregressive model, AR(52), model that only uses historical information as input. As shown in the next sections, we found that all external data sources improve flu estimates, specially in the one- and two-week ahead forecasts.

#### EHR Data and Google Data

Our first modeling experiment involved comparing ARGO models that use Google search and EHR data simultaneously with the baseline AR(52) in all French regions. A detailed analysis on the individual contribution of Google data and EHR data into predictions, separately, is provided for completeness in the supplementary materials. Our findings suggest that each of these data sources individually improves predictions in all time-horizons. This is consistent with the findings of a previous study conducted at the national-level and the French region of Brittany [24], where both Google and EHR information were found meaningful, but EHR data was shown to possess a stronger predictive power.

The join contribution of both EHR and Google data on predictions is presented below. In real time (Table 1), in terms of correlation and error metrics, estimates produced using EHR data and Google data improve the accuracy for all the regions. The combination of both sources lead to correlation improvements of up to 5% and decreases in error of up to 30% for the region Bretagne.

**Table 1.** Real time estimate - MSE and PCC for ARGO models including only historical data (AR(52)) and the 10 most correlated variables from hospital and Google data, for the period starting from January 2015 to March 2017

|  | Auv. | Bour. | Bre. | Cen. | Gd Est | Ht Fra. | Ile Fra. | Norm. | Aqui. | Occi. | Loi. | Pro. |
| --- | --- | --- | --- | --- | --- | --- | --- | --- | --- | --- | --- | --- |
| **MSE** |  |  |  |  |  |  |  |  |  |  |  |  |
| AR(52) | 0.083 | 0.203 | 0.167 | 0.161 | 0.096 | 0.169 | 0.129 | 0.241 | 0.161 | 0.108 | 0.368 | 0.141 |
| ARGO | **0.058** | **0.143** | **0.098** | **0.080** | **0.067** | **0.128** | **0.101** | **0.174** | **0.120** | **0.041** | **0.305** | **0.074** |
| **PCC** |  |  |  |  |  |  |  |  |  |  |  |  |
| AR(52) | 0.958 | 0.898 | 0.916 | 0.919 | 0.952 | 0.915 | 0.935 | 0.879 | 0.919 | 0.946 | 0.815 | 0.929 |
| ARGO | **0.971** | **0.928** | **0.950** | **0.960** | **0.966** | **0.935** | **0.949** | **0.912** | **0.939** | **0.980** | **0.846** | **0.963** |

For One-week ahead estimate (Table 2), estimates obtained with EHR and Google data are more accurate or comparable for 11 of the 12 regions. The combination of both sources lead to correlation improvements of up to 15% and decreases in error of up to 45% for the region Bourgogne.

**Table 2.**
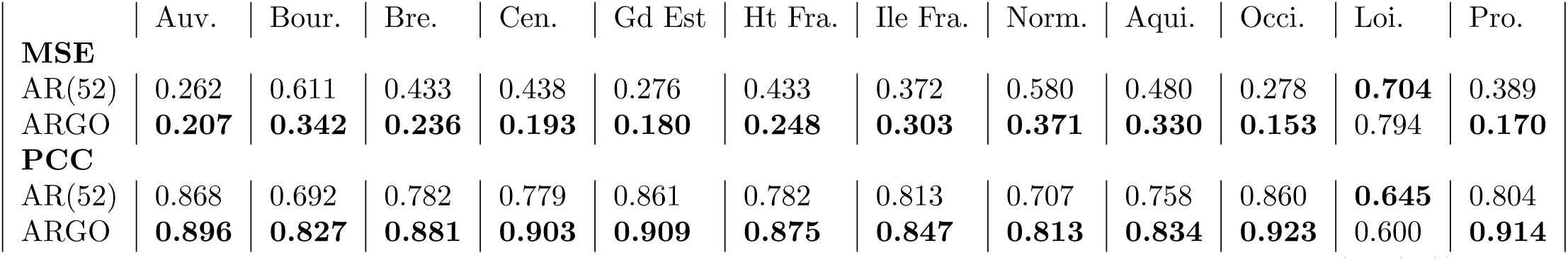
One-week ahead estimate - MSE and PCC for ARGO models including only historical data (AR(52)) and the 10 most correlated variables from hospital and Google data, for the period starting from January 2015 to March 2017

For two-week ahead predictions (Table 3), estimates obtained with EHR and Google data are more accurate for all the regions. The combination of both sources lead to correlation improvements of up to 30% and decreases in error of up to 60% for the region Centre.

**Table 3.** Two-week ahead estimate - MSE and PCC for ARGO models including only historical data (AR(52)) and the 10 most correlated variables from hospital and Google data, for the period starting from January 2015 to March 2017

|  | Auv. | Bour. | Bre. | Cen. | Gd Est | Ht Fra. | Ile Fra. | Norm. | Aqui. | Occi. | Loi. | Pro. |
| --- | --- | --- | --- | --- | --- | --- | --- | --- | --- | --- | --- | --- |
| **MSE** |  |  |  |  |  |  |  |  |  |  |  |  |
| AR(52) | 0.534 | 0.947 | 0.665 | 0.776 | 0.530 | 0.679 | 0.618 | 0.914 | 0.775 | 0.533 | 0.935 | 0.596 |
| ARGO | **0.357** | **0.580** | **0.439** | **0.332** | **0.330** | **0.351** | **0.446** | **0.568** | **0.555** | **0.299** | **0.666** | **0.282** |
| **PCC** |  |  |  |  |  |  |  |  |  |  |  |  |
| AR(52) | 0.731 | 0.522 | 0.665 | 0.609 | 0.733 | 0.658 | 0.688 | 0.539 | 0.610 | 0.731 | 0.528 | 0.699 |
| ARGO | **0.820** | **0.708** | **0.779** | **0.832** | **0.834** | **0.823** | **0.775** | **0.714** | **0.720** | **0.849** | **0.664** | **0.858** |

#### Climatic Data

When combining climatic data with historical activity via ARGO was shown to consistently improve prediction results across all regions (Table 4). However, this improvement is lower than the one observed with EHR and Google data. Indeed, climatic data lead to correlation improvements of 2% and decreases in error of 7% for the region Pays de la Loire.

**Table 4.** Real time estimate - MSE and PCC for ARGO models including only historical data (AR(52)) and only climatic data, for the period starting from January 2015 to March 2017

|  | Auv. | Bour. | Bre. | Cen. | Gd Est | Ht Fra. | Ile Fra. | Norm. | Aqui. | Occi. | Loi. | Pro. |
| --- | --- | --- | --- | --- | --- | --- | --- | --- | --- | --- | --- | --- |
| **MSE** |  |  |  |  |  |  |  |  |  |  |  |  |
| AR(52) | 0.083 | 0.203 | 0.167 | 0.161 | 0.096 | 0.169 | 0.129 | 0.241 | 0.161 | 0.108 | 0.368 | 0.141 |
| ARGO | **0.076** | **0.196** | **0.154** | **0.160** | **0.091** | **0.156** | **0.122** | **0.230** | **0.149** | **0.096** | **0.342** | **0.131** |
| **PCC** |  |  |  |  |  |  |  |  |  |  |  |  |
| AR(52) | 0.958 | 0.898 | 0.916 | 0.919 | 0.952 | 0.915 | 0.935 | 0.879 | 0.919 | 0.946 | 0.815 | 0.929 |
| ARGO | **0.962** | **0.901** | **0.922** | **0.919** | **0.954** | **0.921** | **0.939** | **0.884** | **0.925** | **0.951** | **0.828** | **0.934** |

For one-week ahead estimate (Table 5), in term of correlation and error, results obtained with Climatic data are better or comparable for 11 of the 12 regions. Climatic data lead to correlation improvements of up to 5% and decreases in error of up to 12% for the region Bourgogne-Franche-Comté.

**Table 5.** One-week ahead estimate - MSE and PCC for ARGO models including only historical data (AR(52)) and only climatic data, for the period starting from January 2015 to March 2017

|  | Auv. | Bour. | Bre. | Cen. | Gd Est | Ht Fra. | Ile Fra. | Norm. | Aqui. | Occi. | Loi. | Pro. |
| --- | --- | --- | --- | --- | --- | --- | --- | --- | --- | --- | --- | --- |
| **MSE** |  |  |  |  |  |  |  |  |  |  |  |  |
| AR(52) | **0.262** | 0.611 | 0.433 | 0.438 | 0.276 | 0.433 | 0.372 | 0.580 | 0.480 | **0.278** | 0.704 | 0.389 |
| ARGO | 0.264 | **0.542** | **0.379** | **0.421** | **0.264** | **0.381** | **0.346** | **0.516** | **0.422** | 0.294 | **0.657** | **0.357** |
| **PCC** |  |  |  |  |  |  |  |  |  |  |  |  |
| AR(52) | **0.868** | 0.692 | 0.782 | 0.779 | 0.861 | 0.782 | 0.813 | 0.707 | 0.758 | **0.860** | 0.645 | 0.804 |
| ARGO | 0.867 | **0.726** | **0.809** | **0.788** | **0.867** | **0.808** | **0.825** | **0.740** | **0.787** | 0.852 | **0.669** | **0.820** |

For two-week ahead estimate (Table 6), results obtained with Climatic data are better for all the regions. Climatic data lead to correlation improvements of up to 20% and decreases in error of up to 30% for the region Bourgogne-Franche-Comté.

**Table 6.**
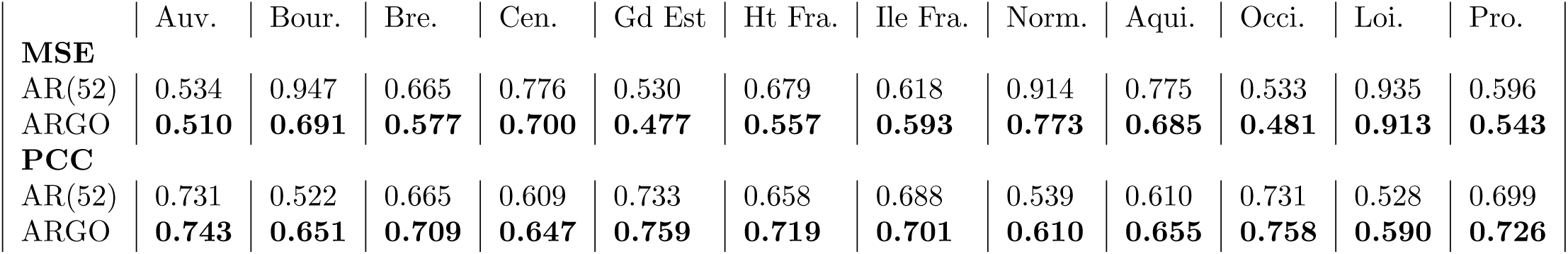
Two-week ahead estimates - MSE and PCC for ARGO models including only historical data (AR(52)) and only climatic data, for the period starting from January 2015 to March 2017

|  | Auv. | Bour. | Bre. | Cen. | Gd Est | Ht Fra. | Ile Fra. | Norm. | Aqui. | Occi. | Loi. | Pro. |
| --- | --- | --- | --- | --- | --- | --- | --- | --- | --- | --- | --- | --- |
| **MSE** |  |  |  |  |  |  |  |  |  |  |  |  |
| AR(52) | 0.534 | 0.947 | 0.665 | 0.776 | 0.530 | 0.679 | 0.618 | 0.914 | 0.775 | 0.533 | 0.935 | 0.596 |
| ARGO | **0.510** | **0.691** | **0.577** | **0.700** | **0.477** | **0.557** | **0.593** | **0.773** | **0.685** | **0.481** | **0.913** | **0.543** |
| **PCC** |  |  |  |  |  |  |  |  |  |  |  |  |
| AR(52) | 0.731 | 0.522 | 0.665 | 0.609 | 0.733 | 0.658 | 0.688 | 0.539 | 0.610 | 0.731 | 0.528 | 0.699 |
| ARGO | **0.743** | **0.651** | **0.709** | **0.647** | **0.759** | **0.719** | **0.701** | **0.610** | **0.655** | **0.758** | **0.590** | **0.726** |

#### Twitter Data

Overall, we found that national-level flu-related Twitter data improves prediction results for all regions.

In real time (Table 7), we see that Twitter data improves results for 8 out of the 12 regions. Twitter data lead to correlation improvements of 2% and decreases in error of 30% for the regions Occitanie and Pays de la Loire.

**Table 7.** Real time estimate - MSE and PCC for ARGO models including only historical data (AR(52)) and only Twitter data, for the period starting from January 2015 to March 2017

|  | Auv. | Bour. | Bre. | Cen. | Gd Est | Ht Fra. | Ile Fra. | Norm. | Aqui. | Occi. | Loi. | Pro. |
| --- | --- | --- | --- | --- | --- | --- | --- | --- | --- | --- | --- | --- |
| **MSE** |  |  |  |  |  |  |  |  |  |  |  |  |
| AR(52) | 0.083 | **0.203** | **0.167** | 0.161 | 0.096 | **0.169** | 0.129 | **0.241** | 0.161 | 0.108 | 0.368 | 0.141 |
| ARGO | **0.078** | 0.212 | 0.170 | **0.137** | **0.091** | 0.170 | **0.109** | 0.257 | **0.160** | **0.078** | **0.337** | **0.133** |
| **PCC** |  |  |  |  |  |  |  |  |  |  |  |  |
| AR(52) | 0.958 | **0.898** | **0.916** | 0.919 | 0.952 | **0.915** | 0.935 | **0.879** | 0.919 | 0.946 | 0.815 | 0.929 |
| ARGO | **0.960** | 0.893 | 0.914 | **0.931** | **0.954** | 0.914 | **0.945** | 0.871 | **0.919** | **0.961** | **0.830** | **0.933** |

For one-week ahead estimate (Table 8), estimates obtained with Twitter data are more accurate for all the regions. Twitter data lead to correlation improvements of 10% and decreases in error of 20% for the region Pays de la Loire.

**Table 8.** One-week ahead estimate - MSE and PCC for ARGO models including only historical data (AR(52)) and only Twitter data, for the period starting from January 2015 to March 2017

|  | Auv. | Bour. | Bre. | Cen. | Gd Est | Ht Fra. | Ile Fra. | Norm. | Aqui. | Occi. | Loi. | Pro. |
| --- | --- | --- | --- | --- | --- | --- | --- | --- | --- | --- | --- | --- |
| **MSE** |  |  |  |  |  |  |  |  |  |  |  |  |
| AR(52) | 0.262 | 0.611 | 0.433 | 0.438 | 0.276 | 0.433 | 0.372 | 0.580 | 0.480 | 0.278 | 0.704 | 0.389 |
| ARGO | **0.236** | **0.570** | **0.355** | **0.331** | **0.251** | **0.422** | **0.305** | **0.548** | **0.376** | **0.232** | **0.589** | **0.330** |
| **PCC** |  |  |  |  |  |  |  |  |  |  |  |  |
| AR(52) | 0.868 | 0.692 | 0.782 | 0.779 | 0.861 | 0.782 | 0.813 | 0.707 | 0.758 | 0.860 | 0.645 | 0.804 |
| ARGO | **0.881** | **0.712** | **0.821** | **0.833** | **0.873** | **0.787** | **0.846** | **0.723** | **0.811** | **0.883** | **0.703** | **0.834** |

For two-week ahead estimate (Table 9), results obtained with Twitter data are more accurate for all the regions. Twitter data lead to correlation improvements of 15% and decreases in error of 20% for the region Bourgogne-Franche-Comté.

**Table 9.** Two-week ahead estimate - MSE and PCC for ARGO models including only historical data (AR(52)) and only Twitter data, for the period starting from January 2015 to March 2017

|  | Auv. | Bour. | Bre. | Cen. | Gd Est | Ht Fra. | Ile Fra. | Norm. | Aqui. | Occi. | Loi. | Pro. |
| --- | --- | --- | --- | --- | --- | --- | --- | --- | --- | --- | --- | --- |
| **MSE** |  |  |  |  |  |  |  |  |  |  |  |  |
| AR(52) | 0.534 | 0.947 | 0.665 | 0.776 | 0.530 | 0.679 | 0.618 | 0.914 | 0.775 | 0.533 | 0.935 | 0.596 |
| ARGO | **0.503** | **0.764** | **0.490** | **0.603** | **0.466** | **0.624** | **0.571** | **0.874** | **0.624** | **0.417** | **0.866** | **0.507** |
| **PCC** |  |  |  |  |  |  |  |  |  |  |  |  |
| AR(52) | 0.731 | 0.522 | 0.665 | 0.609 | 0.733 | 0.658 | 0.688 | 0.539 | 0.610 | 0.731 | 0.528 | 0.699 |
| ARGO | **0.746** | **0.615** | **0.753** | **0.696** | **0.765** | **0.685** | **0.712** | **0.559** | **0.685** | **0.790** | **0.563** | **0.744** |

### Evaluation of Statistical Models

Here, we compare the predictive performance of four different modeling approaches AR(52), ARGO, Net, and ARGONet for three time horizons: real-time, one-week and two-week ahead estimates. Figure 1 displays the ranking of each model for each time horizon of prediction across regions during the out-of-sample evaluation time period (January 2015 to March 2017). If a model is ranked in the 1st position, it means that it led to the best prediction results in terms of error (MSE) and in most cases this was also the case in terms of correlation. As displayed in Figure 1, ARGONet is the most accurate model, ranking either 1st or 2nd in all regions for real-time estimates, and ranking 1st in both the one- and two-week prediction horizons. Further details about each model’s performance are shown in Figures S2 to S13.

**Fig 1.**
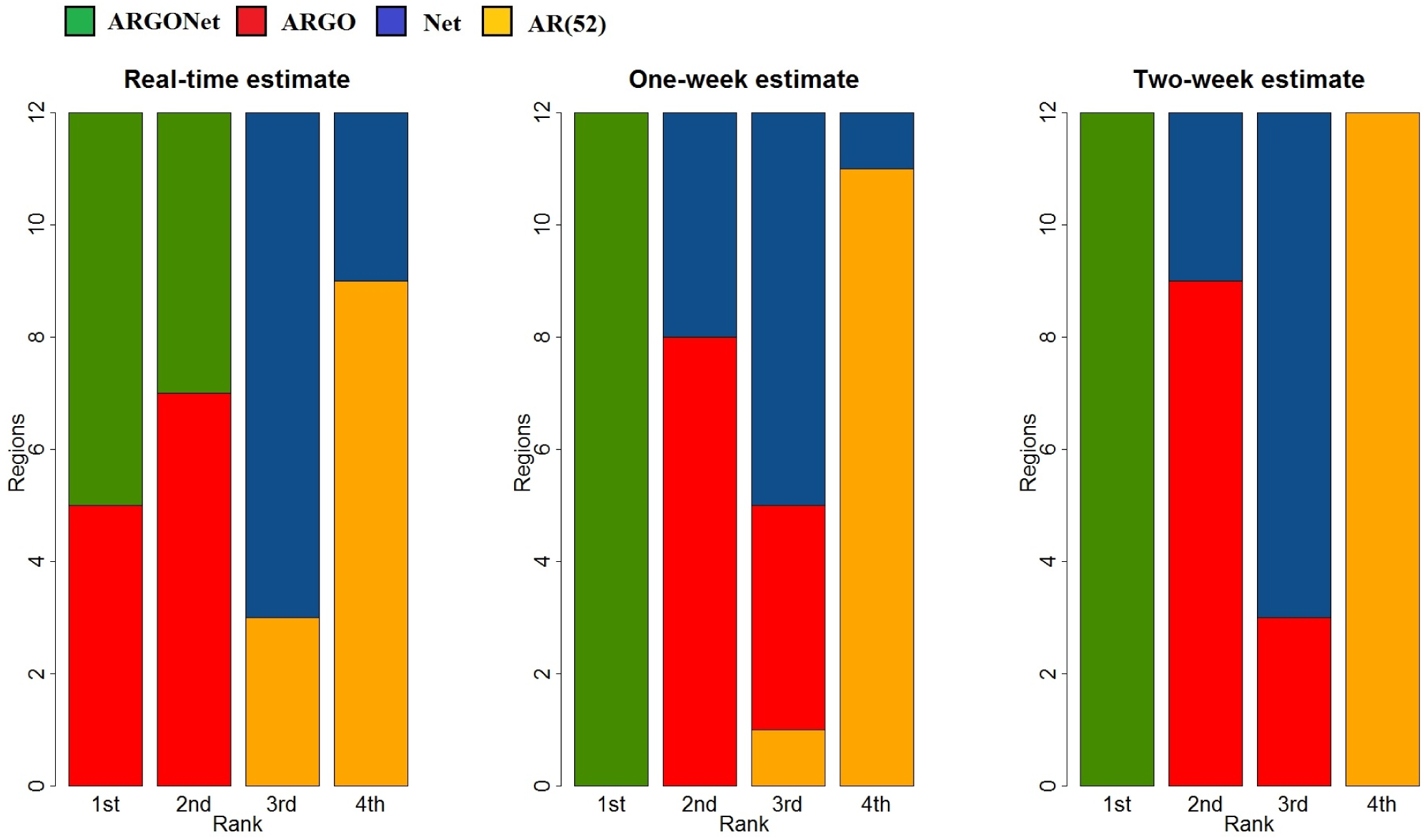
Ranks obtained by each model over the 12 French regions for PCC and MSE.

#### Real-time estimates

Figure 2 and Table 10 summarize results obtained with AR(52), ARGO, Net and ARGONet models for the period starting from January 2015 to March 2017, for the 12 regions. Over this time period, the 90% confidence interval (CI) of the best correlation is [0.915;0.971] with a median value equal to 0.950. The 90% CI of the relative error is [0.057;0.169] with a median value equal to 0.096 which implies a reduction of the error from 5% to 17% thanks to our models.

**Fig 2.**
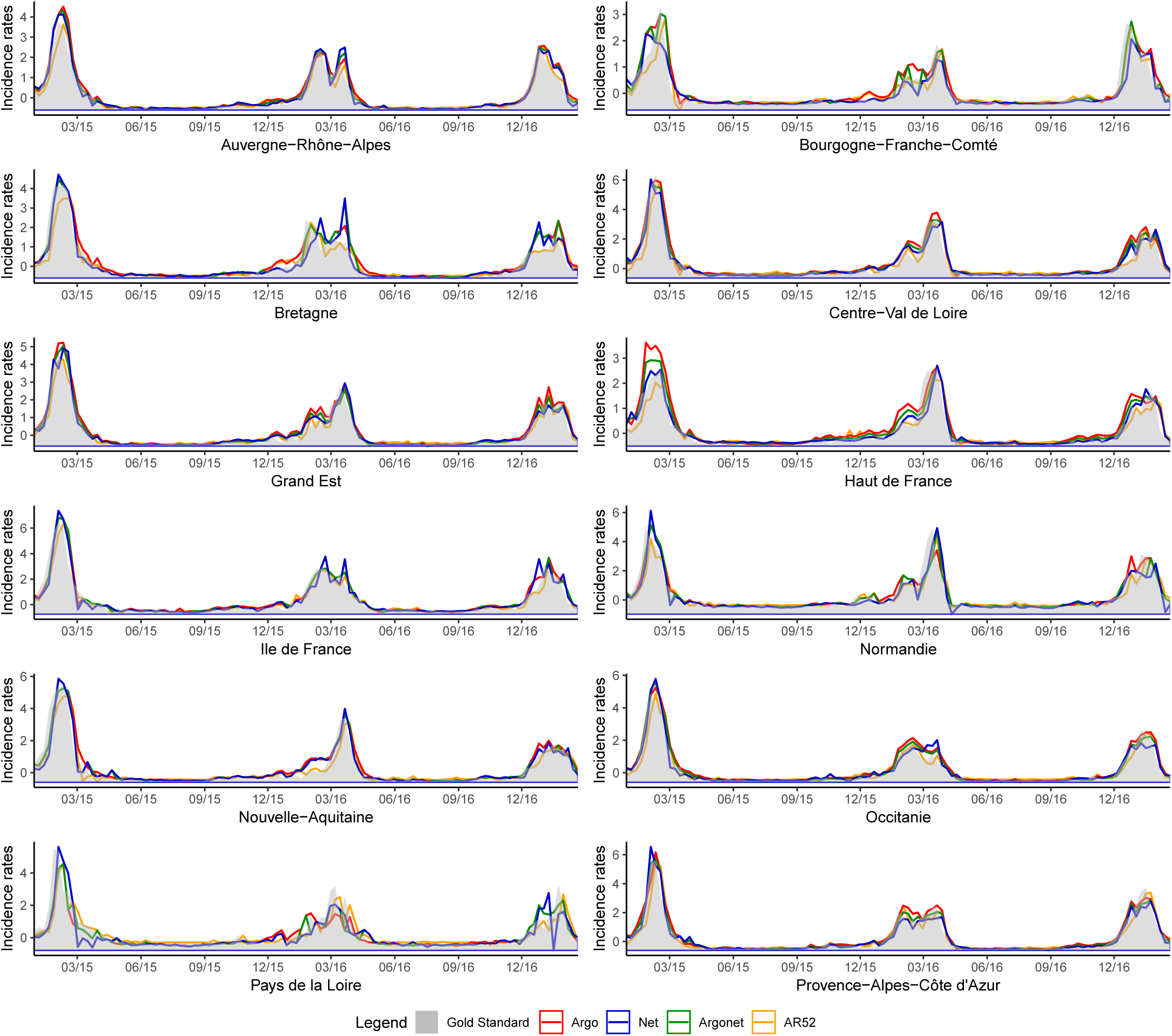
Real-time estimate obtained with ARGO, Net and ARGONet models from January 2015 to March 2017.

**Table 10.** PCC and MSE for real-time estimate for all french regions for the period starting from January 2015 to March 2017

|  | Auv. | Bour. | Bre. | Cen. | Gd Est | Ht Fra. | Ile Fra. | Norm. | Aqui. | Occi. | Loi. | Pro. |
| --- | --- | --- | --- | --- | --- | --- | --- | --- | --- | --- | --- | --- |
| **MSE** |  |  |  |  |  |  |  |  |  |  |  |  |
| AR(52) | 0.083 | 0.203 | 0.167 | 0.161 | 0.096 | 0.169 | 0.129 | 0.241 | 0.161 | 0.108 | 0.368 | 0.141 |
| Argo | 0.059 | **0.138** | **0.098** | **0.083** | **0.067** | 0.121 | **0.098** | **0.169** | 0.118 | **0.042** | 0.284 | 0.072 |
| Net | 0.071 | 0.215 | 0.186 | 0.115 | 0.074 | 0.132 | 0.142 | 0.237 | 0.126 | 0.087 | 0.337 | 0.085 |
| K=1 | **0.057** | 0.154 | 0.134 | 0.095 | 0.064 | 0.128 | 0.120 | 0.180 | **0.094** | 0.057 | 0.309 | 0.078 |
| K=2 | **0.057** | 0.155 | 0.131 | 0.107 | 0.062 | 0.137 | 0.118 | **0.176** | **0.096** | 0.058 | 0.308 | **0.068** |
| K=3 | 0.059 | 0.159 | 0.126 | 0.108 | **0.060** | 0.133 | 0.118 | 0.224 | 0.108 | 0.058 | **0.250** | 0.077 |
| K=4 | 0.066 | 0.160 | 0.141 | 0.103 | 0.071 | 0.174 | 0.108 | 0.188 | 0.110 | 0.049 | 0.260 | 0.086 |
| Mean | **0.057** | 0.153 | 0.125 | **0.088** | **0.061** | **0.108** | 0.110 | 0.180 | 0.112 | 0.052 | **0.258** | **0.060** |
| Lm | **0.068** | **0.147** | **0.102** | 0.096 | 0.062 | **0.118** | **0.103** | 0.243 | 0.124 | **0.042** | 0.316 | 0.069 |
| **PCC** |  |  |  |  |  |  |  |  |  |  |  |  |
| AR(52) | 0.958 | 0.898 | 0.916 | 0.919 | 0.952 | 0.915 | 0.935 | 0.879 | 0.919 | 0.946 | 0.815 | 0.929 |
| Argo | 0.970 | **0.930** | **0.951** | **0.958** | **0.966** | 0.939 | **0.951** | **0.915** | 0.941 | **0.979** | 0.857 | 0.964 |
| Net | 0.964 | 0.892 | 0.906 | 0.942 | 0.963 | 0.933 | 0.928 | 0.881 | 0.936 | 0.956 | 0.830 | 0.957 |
| K=1 | **0.971** | 0.922 | 0.932 | 0.952 | 0.968 | 0.935 | 0.939 | 0.909 | **0.952** | 0.971 | 0.844 | 0.961 |
| K=2 | **0.971** | 0.922 | 0.934 | 0.946 | 0.969 | 0.931 | 0.941 | **0.911** | **0.952** | 0.971 | 0.845 | **0.966** |
| K=3 | 0.970 | 0.920 | 0.936 | 0.946 | **0.970** | 0.933 | 0.940 | 0.887 | 0.946 | 0.971 | **0.874** | 0.961 |
| K=4 | 0.967 | 0.919 | 0.929 | 0.948 | 0.964 | 0.912 | 0.945 | 0.905 | 0.944 | 0.975 | 0.869 | 0.956 |
| Mean | **0.971** | 0.923 | 0.937 | **0.955** | **0.969** | **0.946** | 0.944 | 0.909 | 0.943 | 0.974 | **0.870** | **0.970** |
| Lm | 0.966 | **0.926** | **0.948** | 0.952 | 0.969 | **0.940** | **0.948** | 0.878 | 0.938 | **0.979** | 0.841 | 0.965 |

Figure 3 confirms, region by region, that the best PCC and MSE are mostly obtained with ARGONet for real-time predictions. In this time-horizon, ARGO shows good performance.

**Fig 3.**
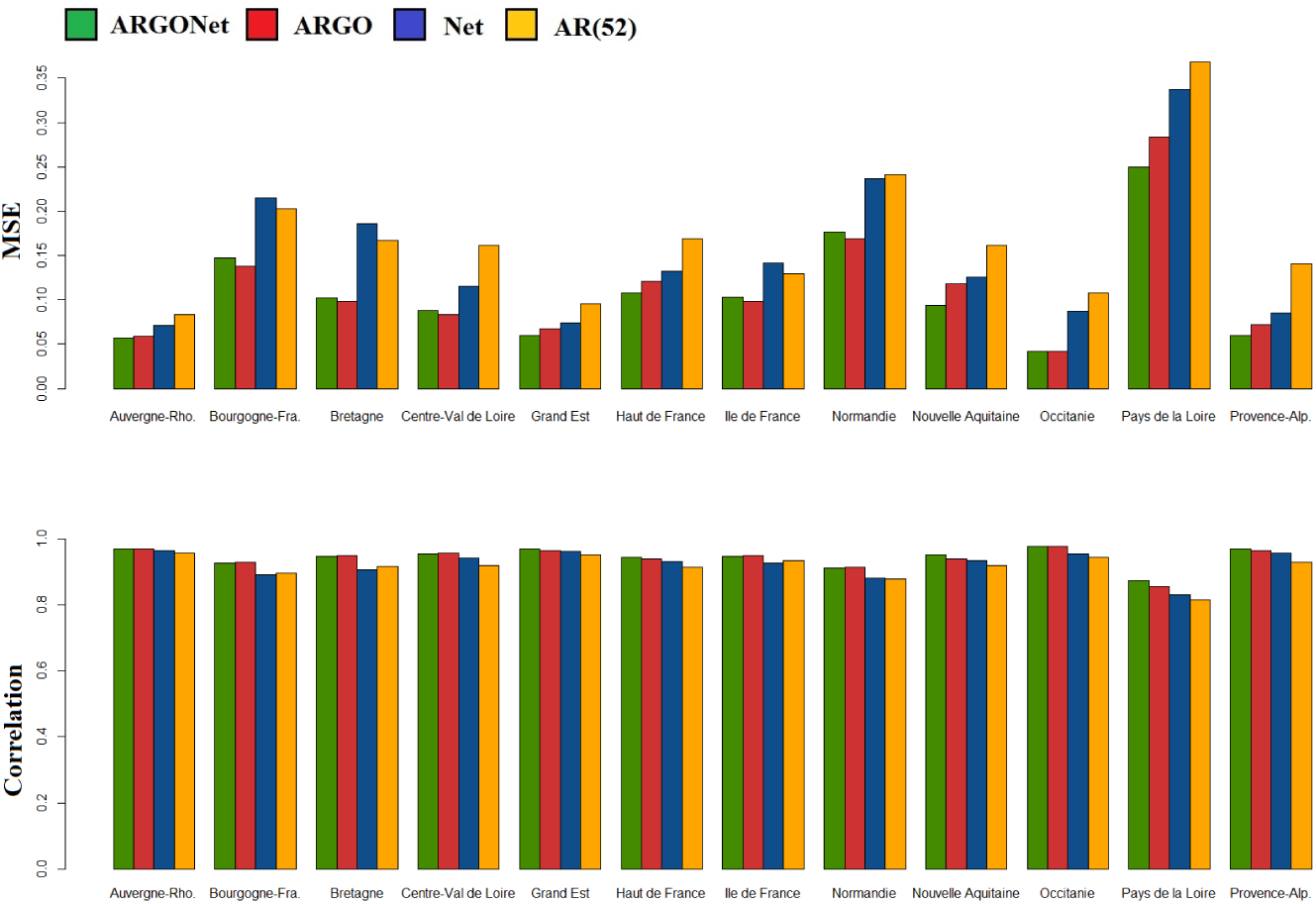
PCC and MSE obtained for real-time estimate with ARGO, Net and ARGONet models.

For 5 regions the best model is ARGO with the highest PCC and the lowest MSE. For the other 7 regions, the best model is ARGONet. For the one- and two-week time horizons, Figures 7, 11, 14 and 15 confirm that ARGONet outperforms all other models.

**Fig 4.**
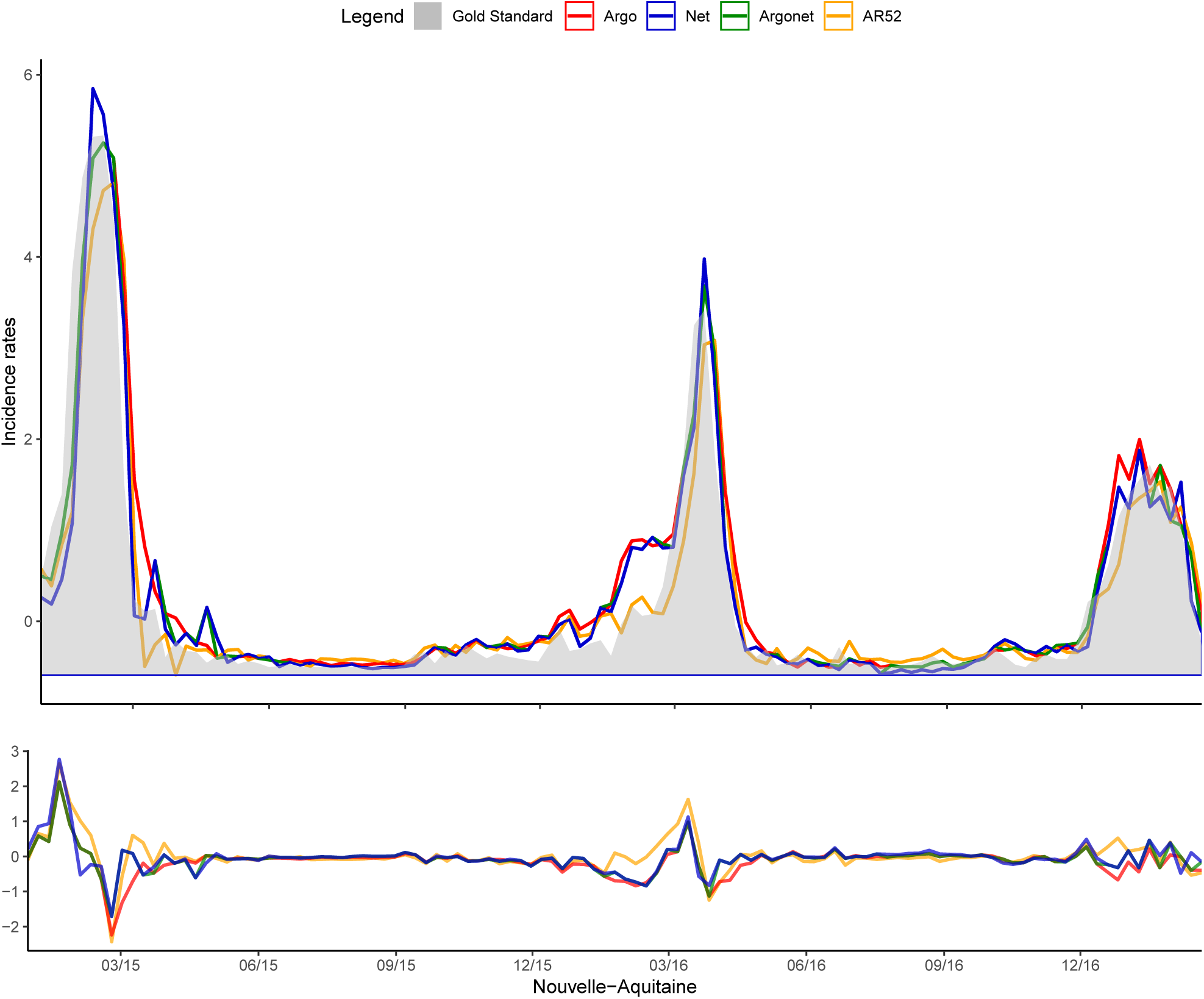
Nouvelle-Aquitaine Real time estimate.

**Fig 5.**
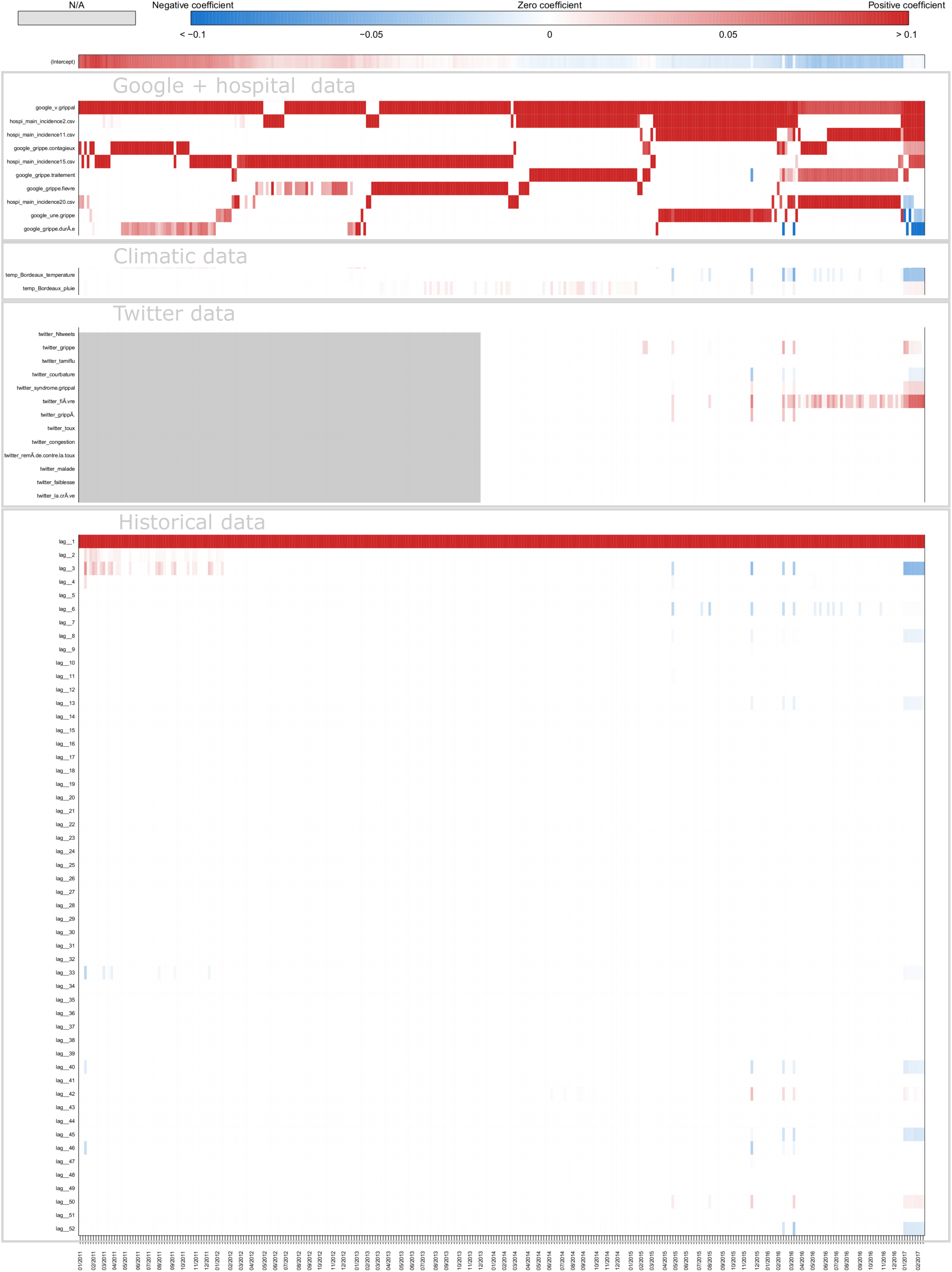
Coefficients Nouvelle-Aquitaine Real-time estimate.

**Fig 6.**
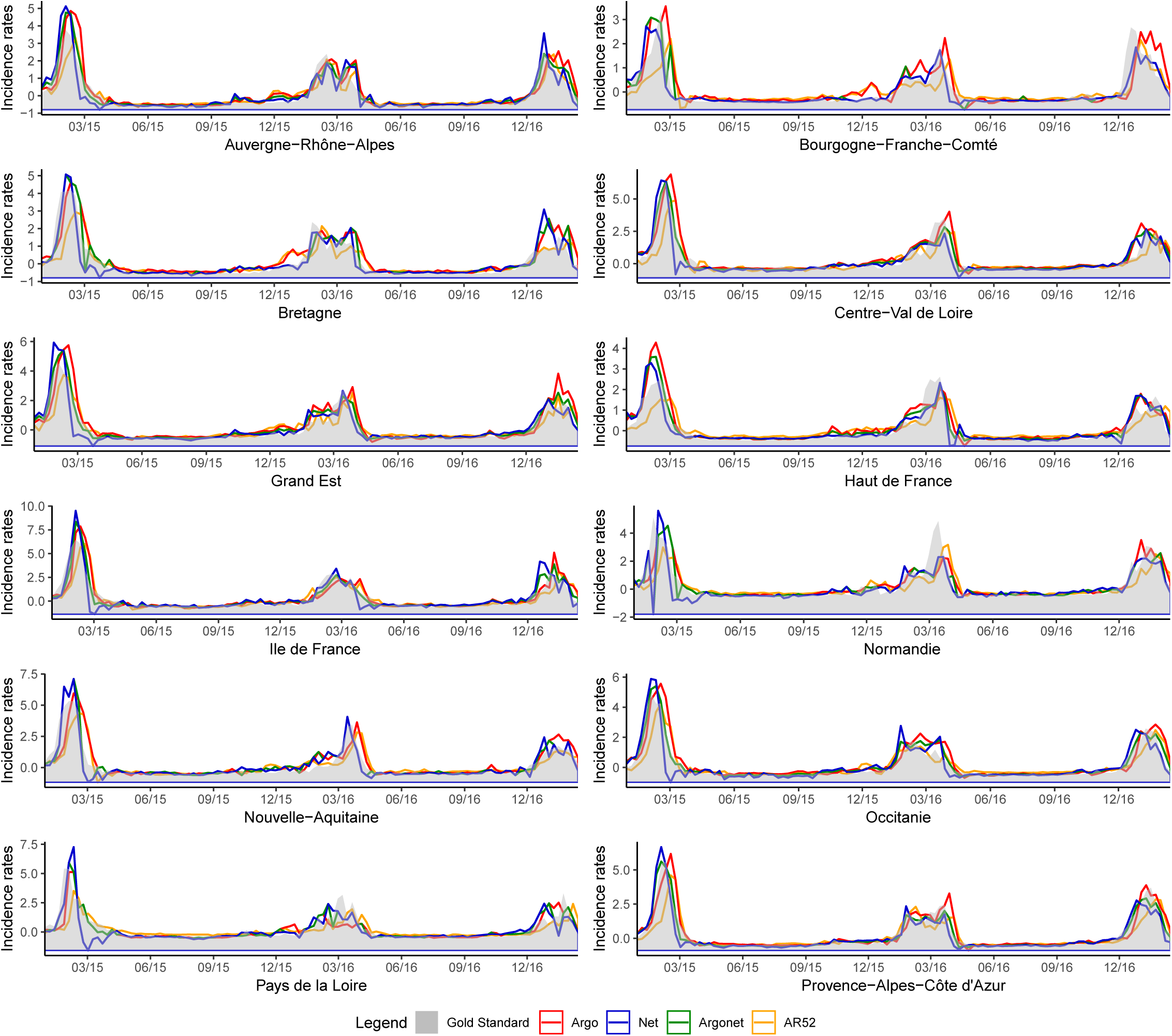
One-week ahead estimate obtained with ARGO, Net and ARGONet models from January 2015 to March 2017.

**Fig 7.**
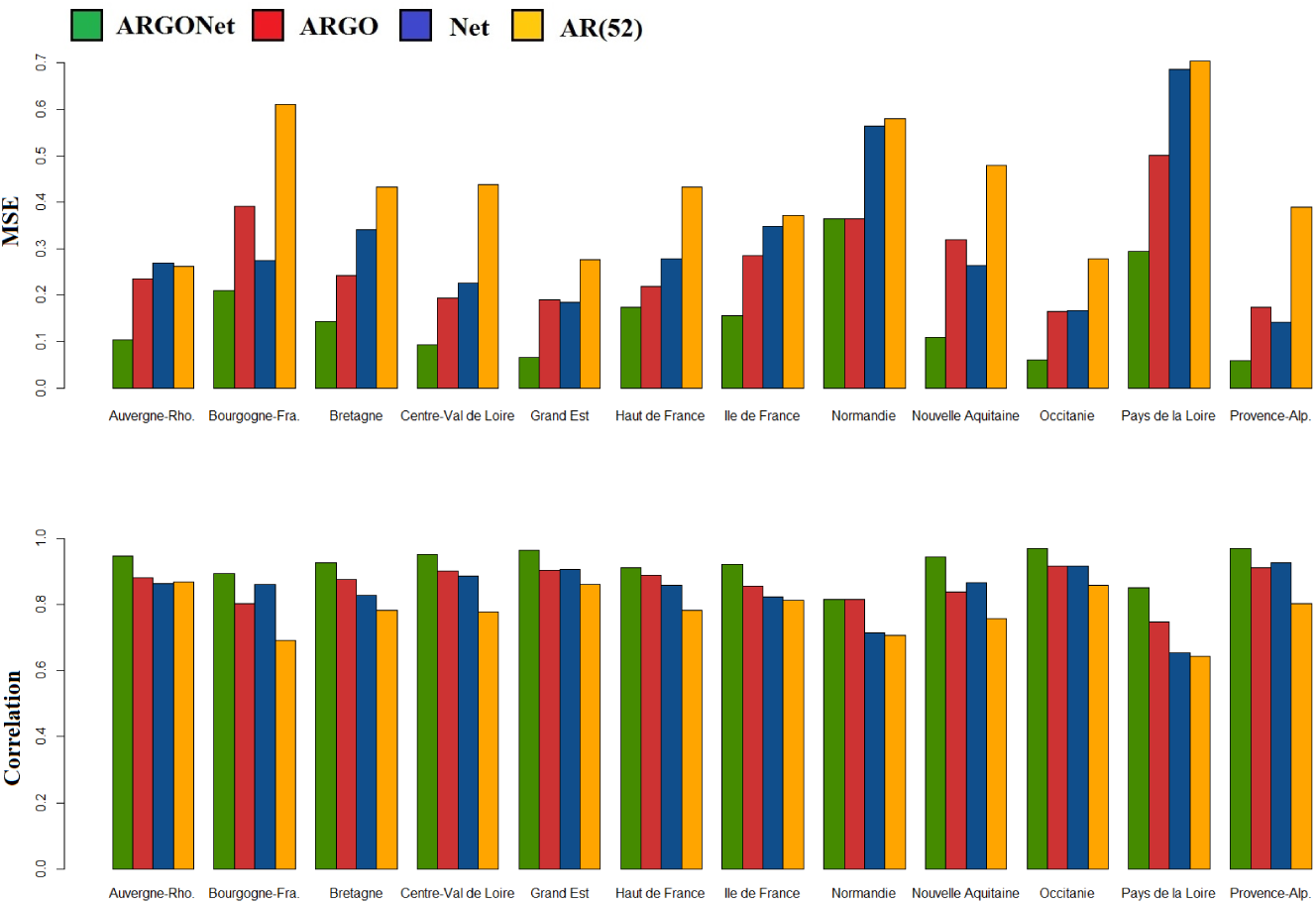
PCC and MSE obtained for one-week ahead estimate with ARGO, Net and ARGONet models.

**Fig 8.**
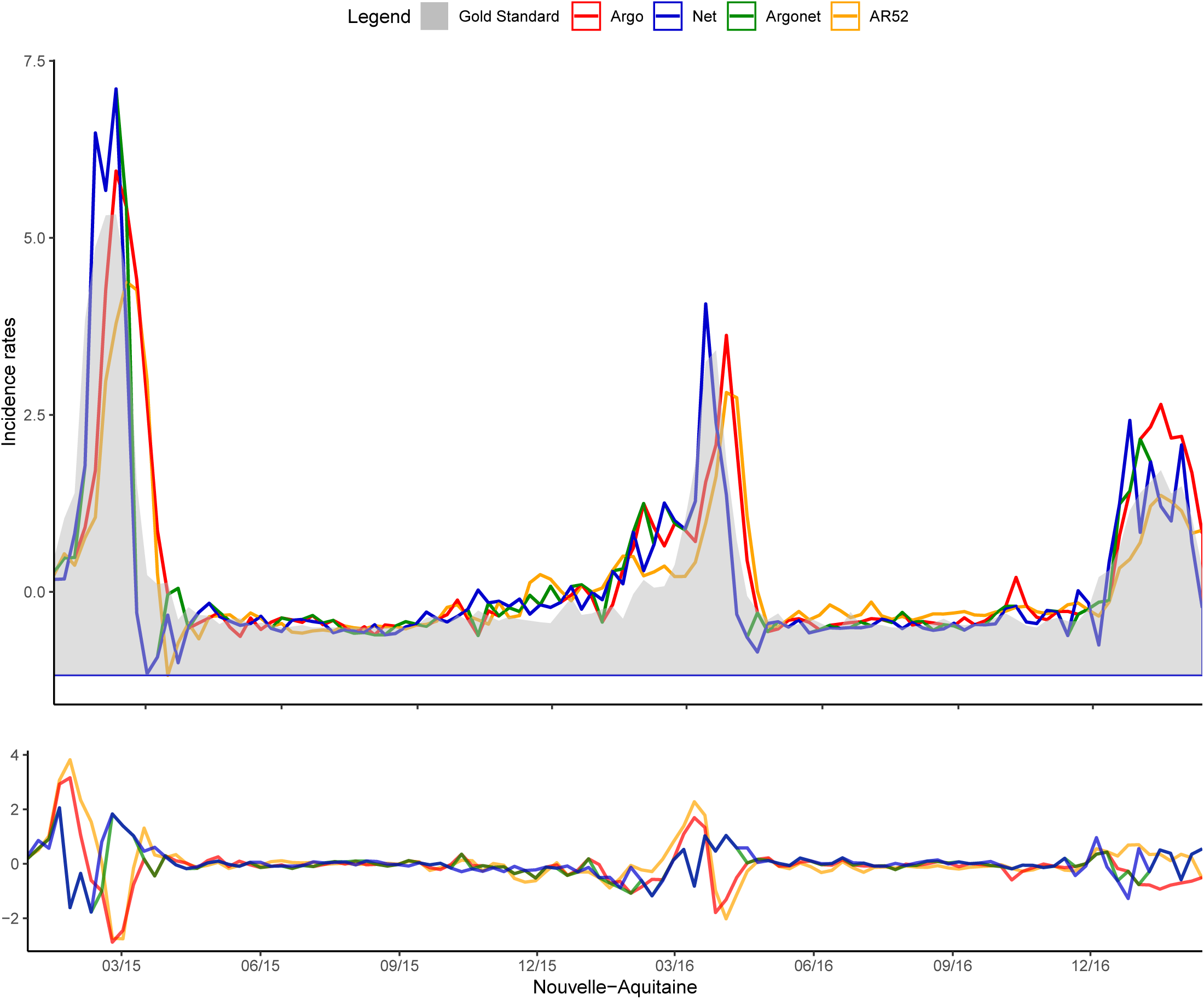
Nouvelle-Aquitaine one-week ahead estimate.

**Fig 9.**
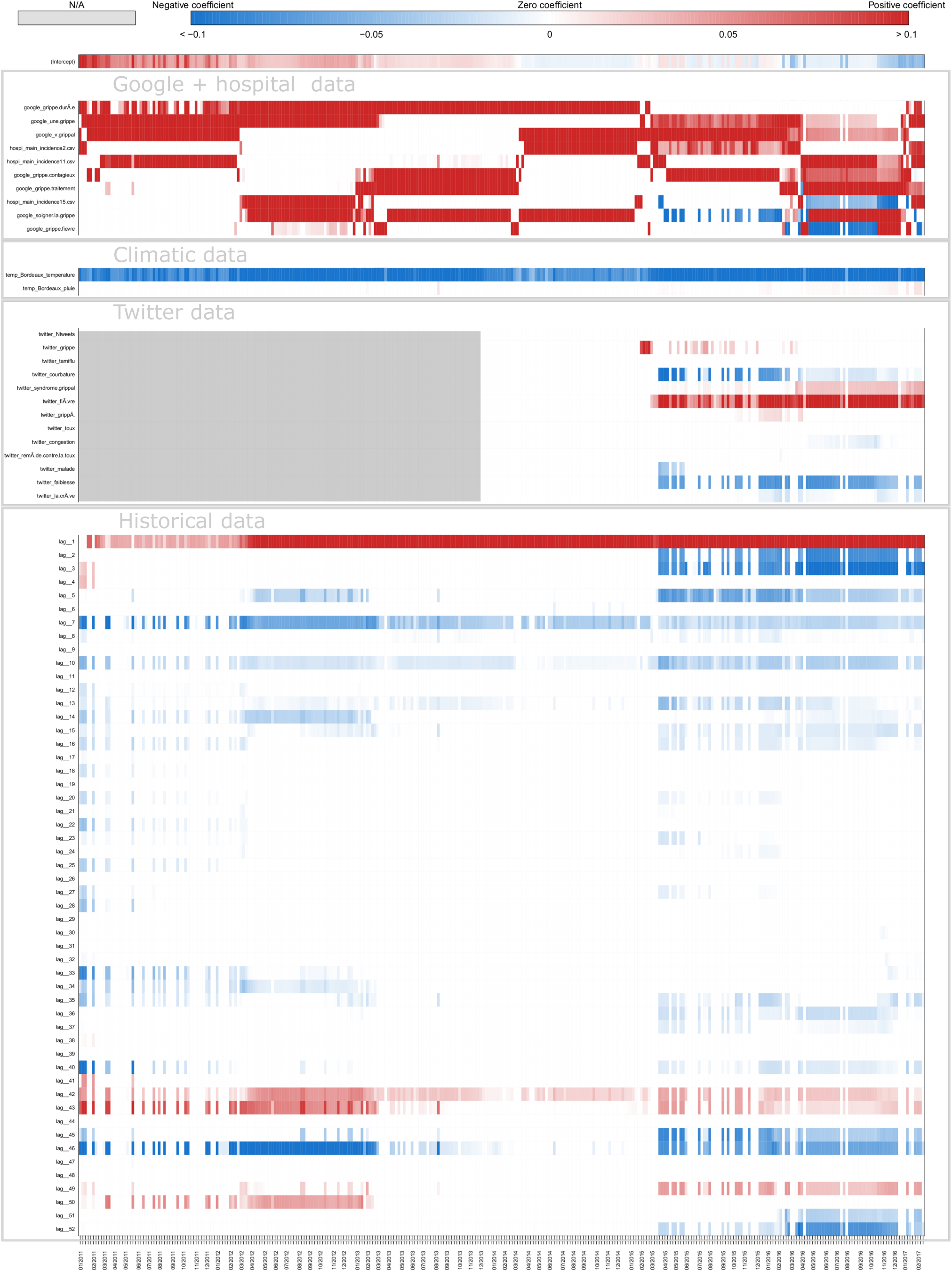
Coefficients Nouvelle-Aquitaine one-week ahead estimate.

**Fig 10.**
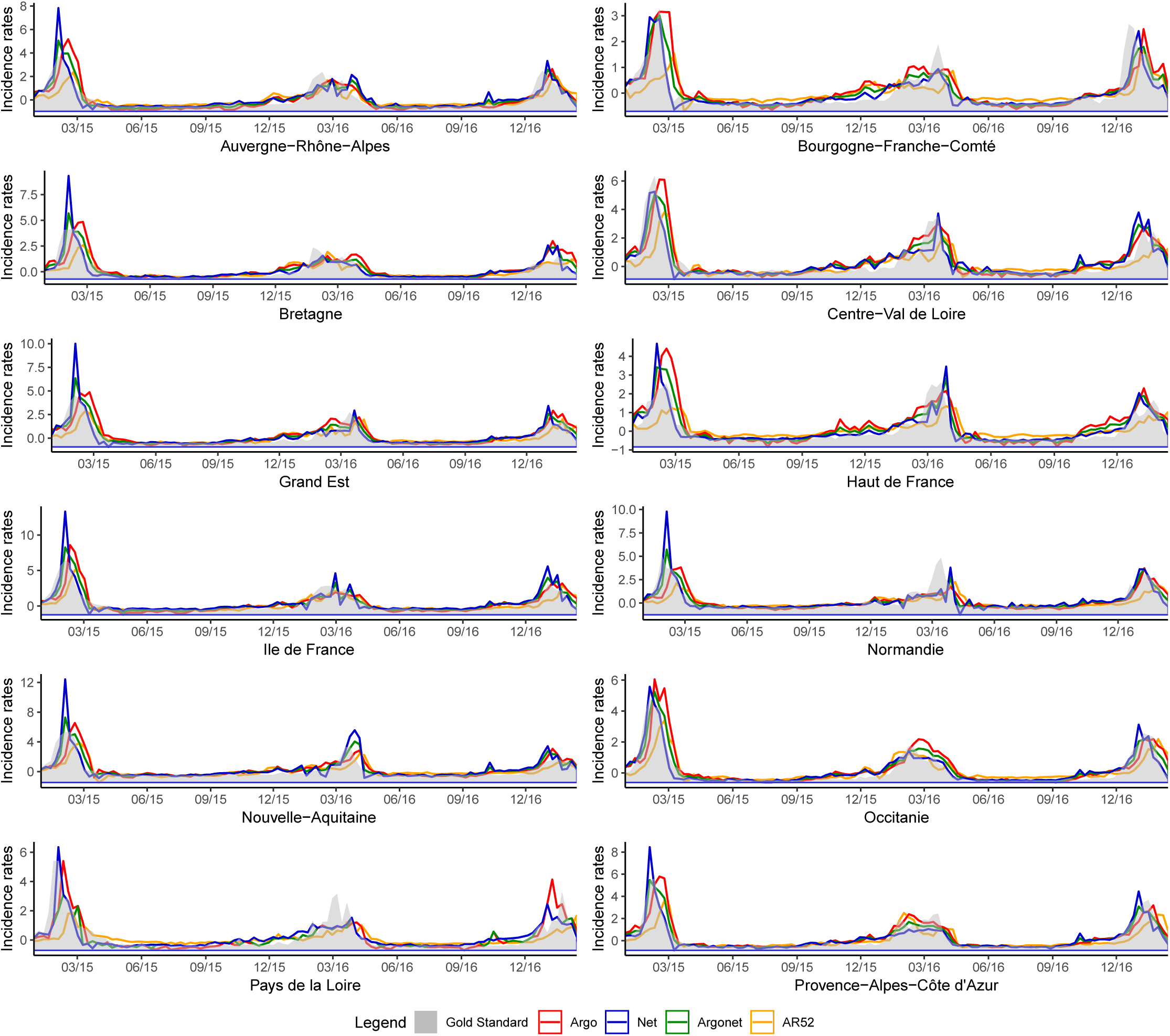
Two-week ahead estimate obtained with ARGO, Net and ARGONet models from January 2015 to March 2017.

**Fig 11.**
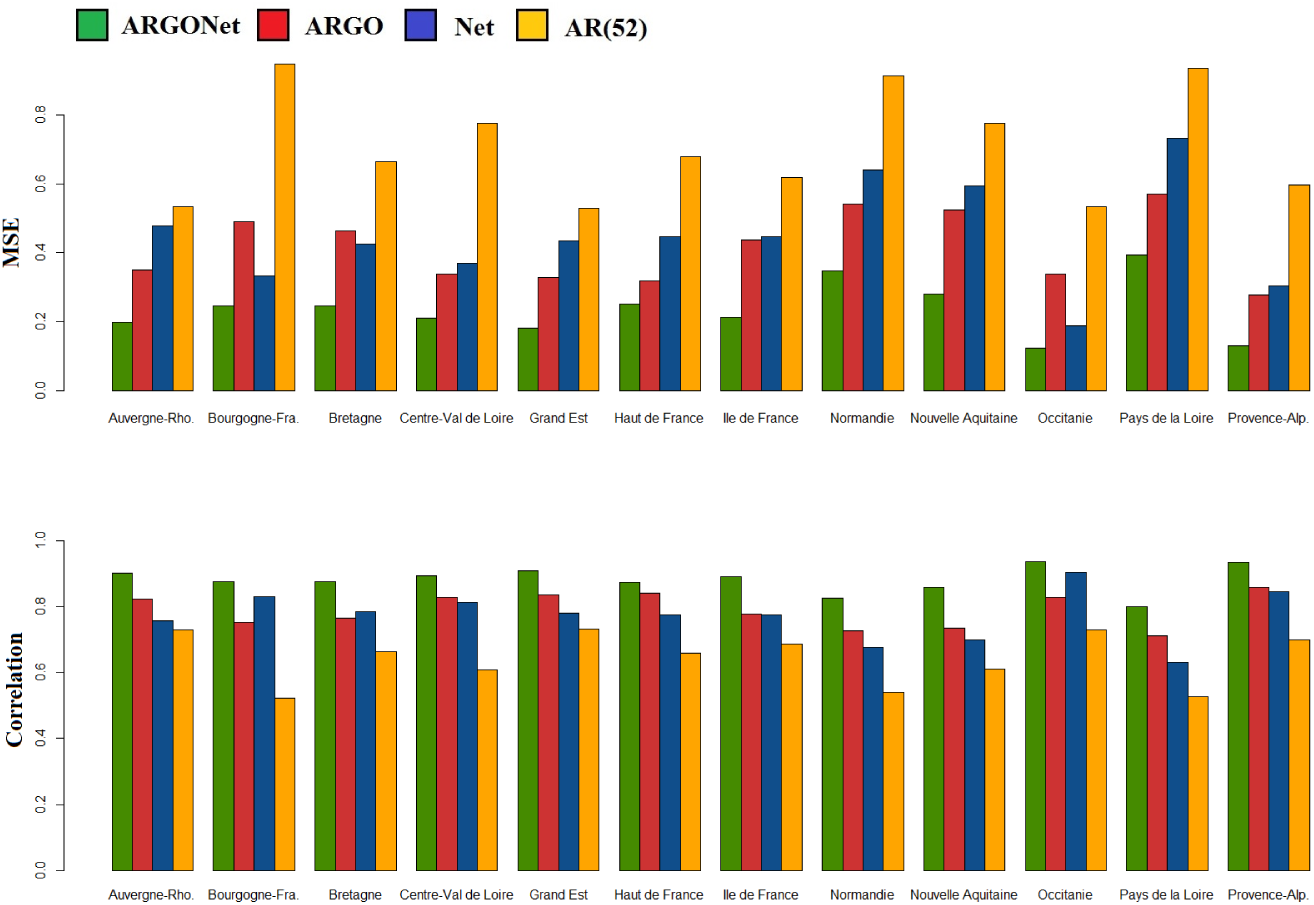
PCC and MSE obtained for two-week ahead estimate with ARGO, Net and ARGONet models.

**Fig 12.**
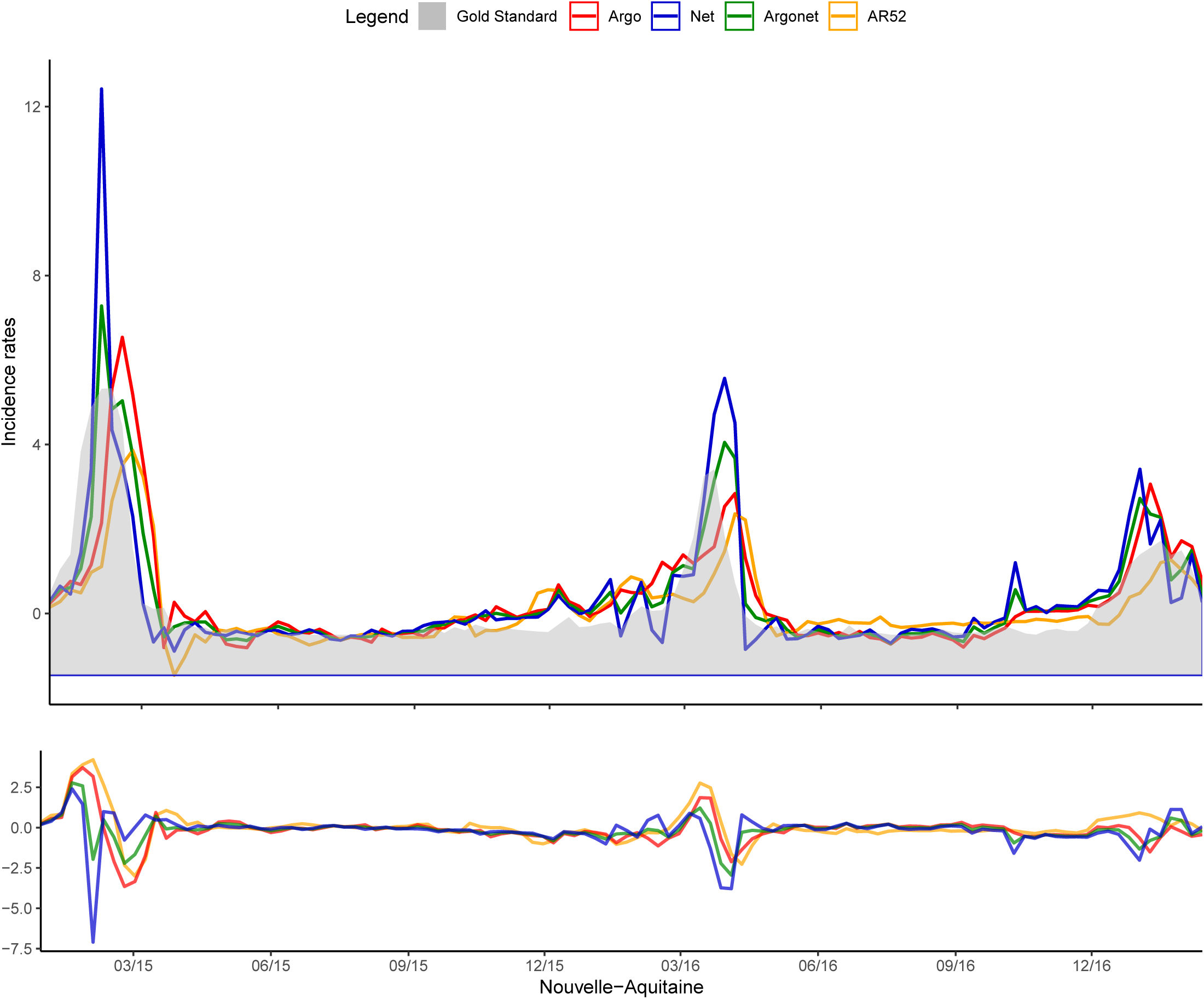
Nouvelle-Aquitaine Two-week ahead estimate.

**Fig 13.**
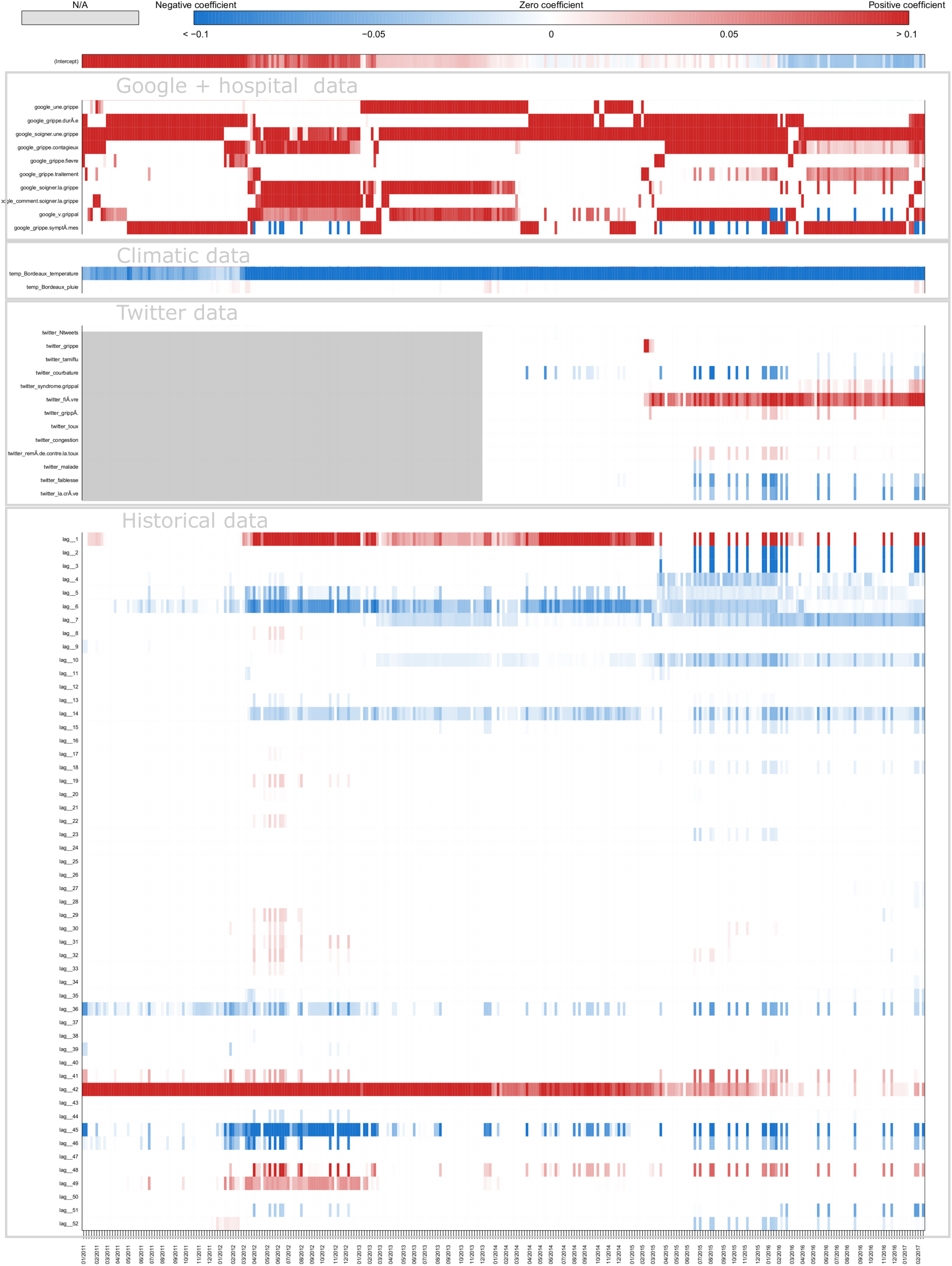
Coefficients Nouvelle-Aquitaine Two-week ahead estimate.

**Fig 14.**
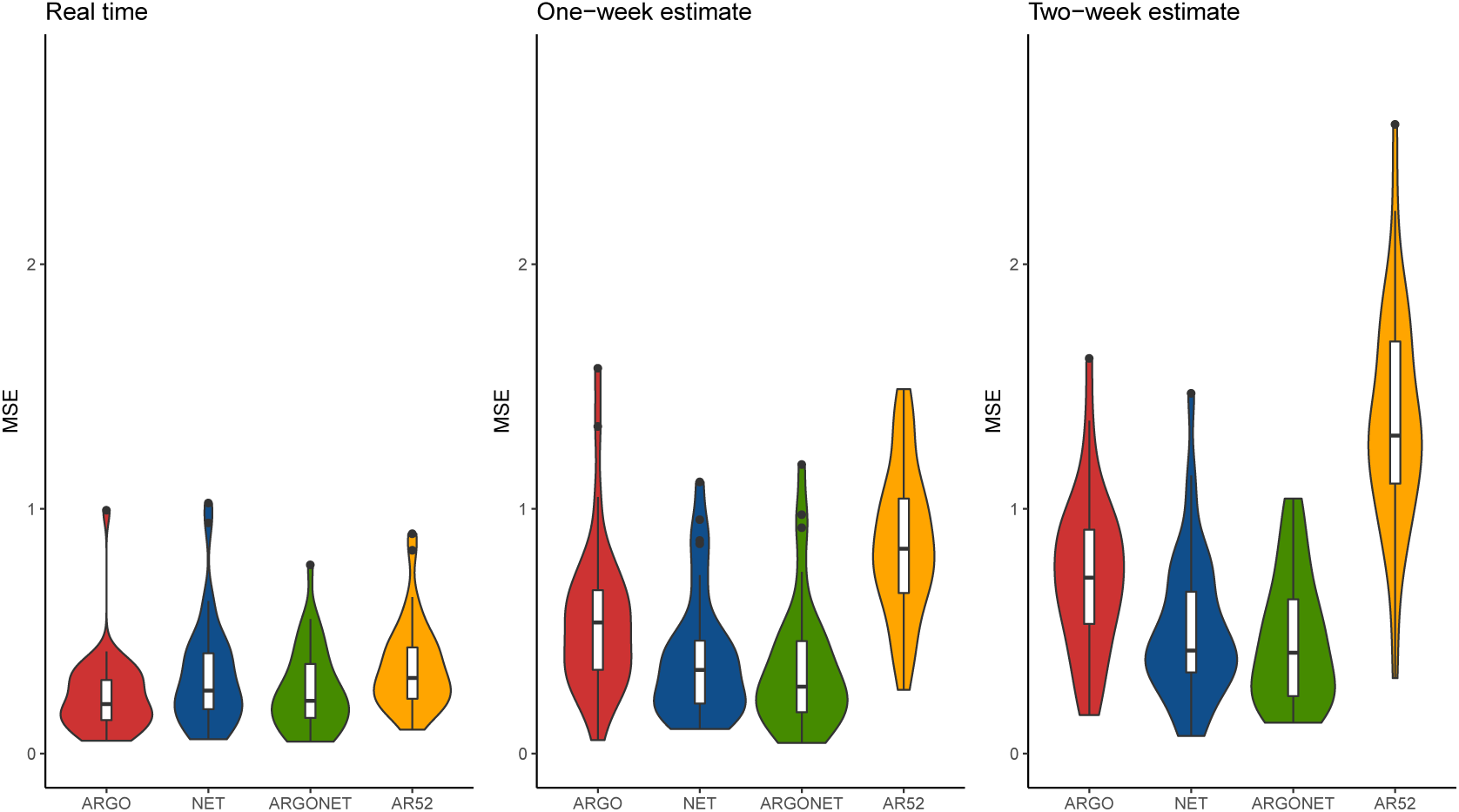
Error distribution.

**Fig 15.**
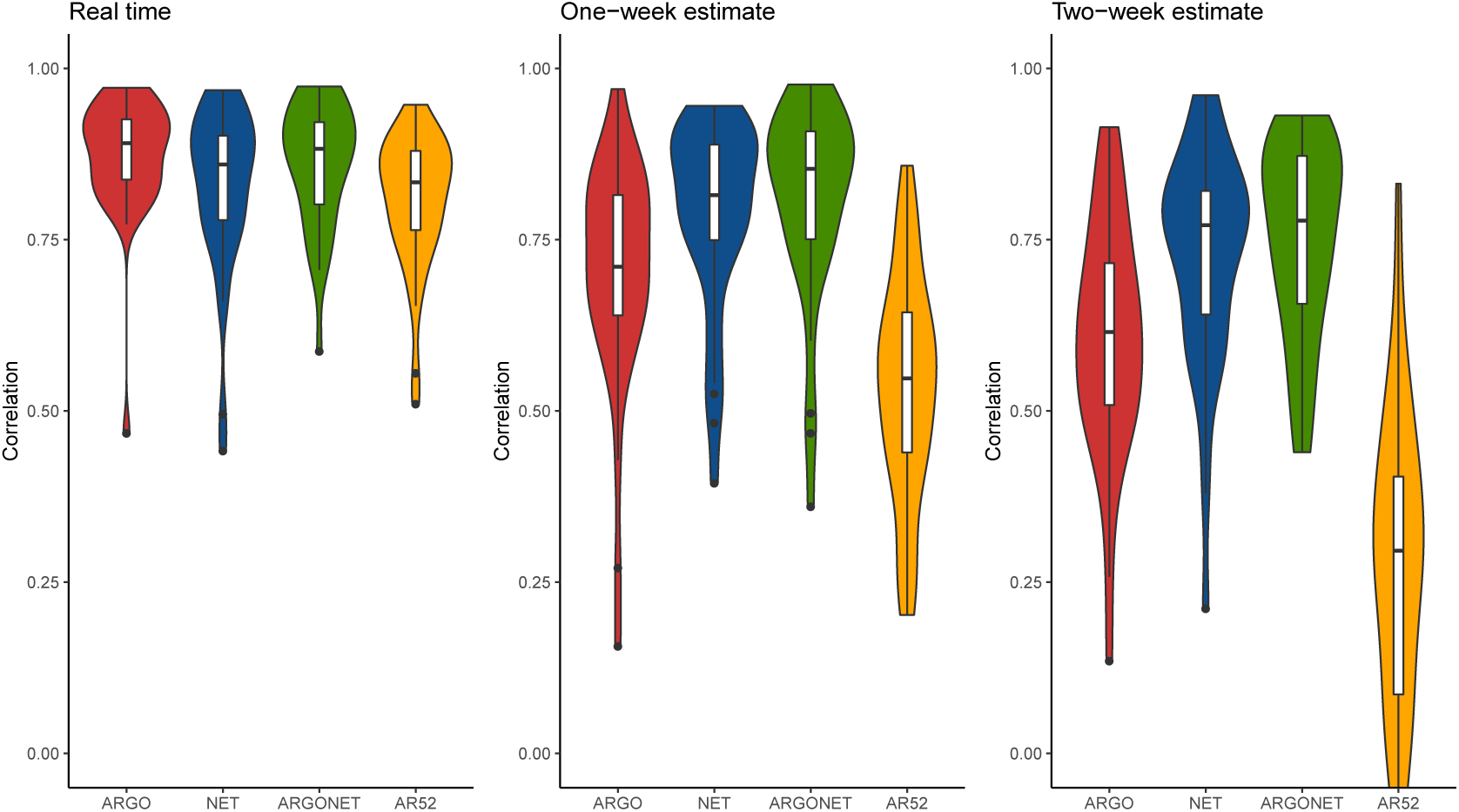
Correlation distribution.

To assess the statistical significance of the improved prediction power of ARGONet, we constructed a 95% confidence interval for the relative efficiency of ARGONet compared to the autoregressive model (the error of ARGONet is in the denominator). Table 11 shows that in real-time, the improvement obtained thanks to the ARGONet model compared to the autoregressive model is statistically significant for all regions. Depending on the region, ARGONet allows to reduce the error by 15% to 60%.

**Table 11.** Real-time estimate - Relative efficiency being bigger than 1 suggests increased predictive power of ARGONet compared to the autoregressive model

| Region | Relative efficiency | 95% CI |
| --- | --- | --- |
| Auv. | 1.43 | [1.35;1.52] |
| Bour. | 1.38 | [1.29;1.48] |
| Bre. | 1.62 | [1.52;1.72] |
| Cen. | 1.75 | [1.65;1.86] |
| Gd Est | 1.78 | [1.67;1.89] |
| Ht Fra. | 1.89 | [1.71;2.07] |
| Ile Fra. | 1.18 | [1.13;1.24] |
| Norm. | 1.47 | [1.38;1.57] |
| Aqui. | 1.63 | [1.56;1.71] |
| Occi. | 2.43 | [2.27;2.59] |
| Loi. | 1.38 | [1.29;1.48] |
| Pro. | 2.41 | [2.14;2.69] |

Figure 4 and Figure 5 show a typical example of plot and heatmap obtained for estimates in real time. The heatmap allows to visualize coefficients used for ARGO model. On these plots, we can see that all models have estimates close to the gold standard. However, for the autoregressive model, there is a time lag more important. On the heatmap, we can see that ARGO model uses mostly 5 variables including 2 variables from Google Data, 2 variables from Hospital Data and 1 variable from Historical Data.

#### One-week ahead estimates

Figure 6 and Table 12 show results for one-week ahead forecasts for the time period January 2015-March 2017. Over this time period, the 90% CI of the best correlation is [0.852;0.970] with a median value equal to 0.936. The 90% CI of the relative error is [0.060;0.294] with a median value equal to 0.127 which implies a reduction of the error from 6% to 30% thanks to our models.

**Table 12.**
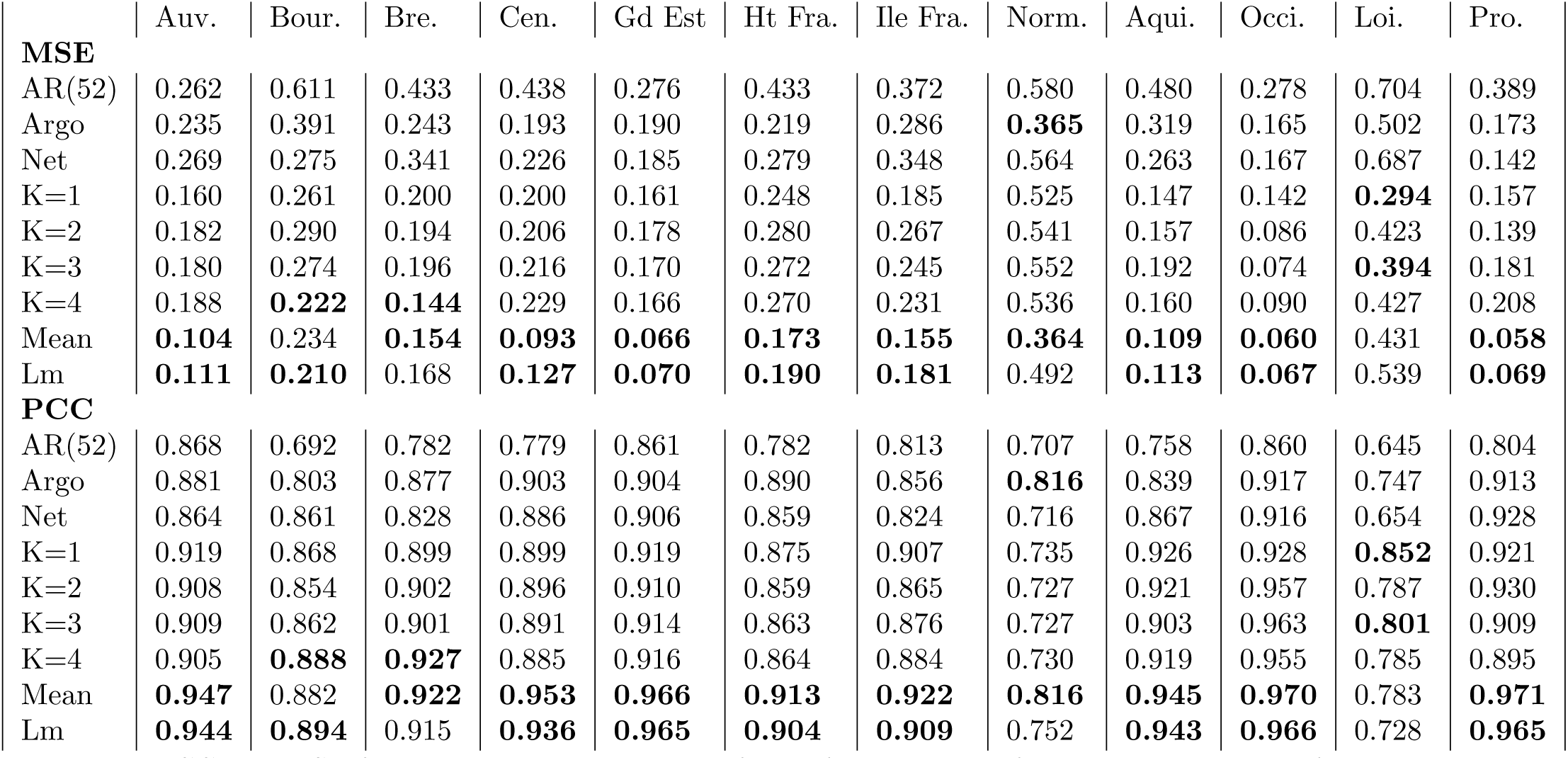
PCC and MSE for one-week ahead estimate for all french regions for the period starting from January 2015 to March 2017

**Table 13.** One-week ahead estimate - Relative efficiency being bigger than 1 suggests increased predictive power of ARGONet compared to the autoregressive model

| Region | Relative efficiency | 95% CI |
| --- | --- | --- |
| Auv. | 2.56 | [2.40;2.73] |
| Bour. | 3.56 | [3.04;3.68] |
| Bre. | 3.14 | [2.88;3.39] |
| Cen. | 4.95 | [4.36;5.54] |
| Gd Est | 4.24 | [3.83;4.64] |
| Ht Fra. | 2.90 | [2.65;3.16] |
| Ile Fra. | 2.39 | [2.17;2.61] |
| Norm. | 2.20 | [1.98;2.42] |
| Aqui. | 5.06 | [4.55;5.58] |
| Occi. | 4.60 | [4.13;5.07] |
| Loi. | 2.83 | [2.45;3.21] |
| Pro. | 7.79 | [6.85;8.73] |

For one-week ahead forecasts the best model is ARGONet. AR(52) is the model giving the worst results, but, in contrast to real-time results, ARGO and Net models are comparable. Indeed, for 4 regions, Net model allows to have better results than ARGO model. We can also observe these results on barplots (Figure 7) and on the distribution of correlation and error (Figures 14 and 15).

Table 12 shows that the improvement obtained thanks to the ARGONet model compared to the autoregressive model is statistically significant for all regions for one-week ahead estimate. Depending on the region, ARGONet allows to reduce the error by 55% to 87%.

Figure 8 shows one-week ahead estimate obtained for the french region Nouvelle-Aquitaine. On this plot, we can see that AR(52) and ARGO models still have a lag of one or two weeks. It is not the case for Net and ARGONet models. On this plot, estimates obtained with Net and ARGONet models are comparable. Figure 9, the heatmap shows that ARGO model uses mostly 9 variables including 2 variables from Google Data, 2 variables from Hospital Data, one variable from Climatic data and one variable from Historical data.

#### Two-week ahead estimate

Figure 10 and Table 14 show results for two-week ahead forecasts for the time period January 2015-March 2017. Over this time period, the 90% CI of the best correlation is [0.825;0.935] with a median value equal to 0.885. The 90% CI of the relative error is [0.129;0.347] with a median value equal to 0.229 which implies a reduction of the error from 13% to 35% thanks to our models. Like for real-time and one-week ahead forecasts, AR(52) is the model giving the worst estimates. For all french regions, the best model is ARGONet with the method using the mean between estimates obtained from ARGO and Net models.

**Table 14.** PCC and MSE for two-week ahead estimate for all french regions for the period starting from January 2015 to March 2017

|  | Auv. | Bour. | Bre. | Cen. | Gd Est | Ht Fra. | Ile Fra. | Norm. | Aqui. | Occi. | Loi. | Pro. |
| --- | --- | --- | --- | --- | --- | --- | --- | --- | --- | --- | --- | --- |
| **MSE** |  |  |  |  |  |  |  |  |  |  |  |  |
| AR(52) | 0.534 | 0.947 | 0.665 | 0.776 | 0.530 | 0.679 | 0.618 | 0.914 | 0.775 | 0.533 | 0.935 | 0.596 |
| Argo | 0.350 | 0.490 | 0.464 | 0.339 | 0.327 | 0.318 | 0.437 | 0.541 | 0.525 | 0.339 | 0.570 | 0.277 |
| Net | 0.479 | 0.334 | 0.424 | 0.369 | 0.434 | 0.446 | 0.446 | 0.640 | 0.594 | 0.188 | 0.732 | 0.304 |
| K=1 | 0.259 | 0.309 | **0.335** | 0.321 | **0.268** | 0.321 | 0.296 | 0.514 | **0.345** | 0.205 | 0.619 | 0.262 |
| K=2 | 0.286 | **0.275** | 0.339 | 0.305 | 0.274 | 0.285 | 0.284 | 0.551 | 0.369 | 0.190 | 0.497 | 0.217 |
| K=3 | 0.278 | 0.307 | 0.341 | 0.303 | 0.291 | 0.302 | 0.284 | 0.549 | 0.356 | 0.239 | **0.466** | 0.288 |
| K=4 | 0.283 | 0.368 | 0.344 | 0.333 | 0.297 | 0.292 | **0.282** | 0.514 | 0.461 | 0.197 | 0.496 | 0.278 |
| Mean | **0.197** | 0.284 | **0.246** | **0.209** | **0.180** | **0.251** | **0.211** | **0.347** | **0.281** | **0.123** | **0.393** | **0.129** |
| Lm | **0.227** | **0.246** | 0.408 | **0.293** | 0.330 | **0.268** | 0.298 | **0.506** | 0.488 | **0.186** | 0.508 | **0.165** |
| **PCC** |  |  |  |  |  |  |  |  |  |  |  |  |
| AR(52) | 0.731 | 0.522 | 0.665 | 0.609 | 0.733 | 0.658 | 0.688 | 0.539 | 0.610 | 0.731 | 0.528 | 0.699 |
| Argo | 0.823 | 0.753 | 0.766 | 0.829 | 0.835 | 0.840 | 0.779 | 0.727 | 0.735 | 0.829 | 0.712 | 0.860 |
| Net | 0.759 | 0.832 | 0.786 | 0.814 | 0.781 | 0.775 | 0.775 | 0.677 | 0.700 | 0.905 | 0.631 | 0.847 |
| K=1 | 0.869 | 0.844 | **0.831** | 0.838 | **0.865** | 0.838 | 0.850 | 0.740 | **0.826** | 0.897 | 0.688 | 0.868 |
| K=2 | 0.856 | **0.861** | 0.829 | 0.846 | 0.862 | 0.856 | 0.857 | 0.722 | 0.814 | 0.904 | 0.749 | 0.891 |
| K=3 | 0.860 | 0.845 | 0.828 | 0.847 | 0.853 | 0.848 | 0.857 | 0.723 | 0.820 | 0.879 | **0.765** | 0.855 |
| K=4 | 0.857 | 0.814 | 0.826 | 0.832 | 0.850 | 0.853 | **0.858** | 0.741 | 0.767 | 0.901 | 0.750 | 0.860 |
| Mean | **0.901** | 0.857 | **0.876** | **0.895** | **0.909** | **0.873** | **0.893** | **0.825** | **0.858** | **0.938** | **0.802** | **0.935** |
| Lm | **0.886** | **0.876** | 0.795 | **0.852** | 0.833 | **0.865** | 0.850 | **0.745** | 0.754 | **0.906** | 0.744 | **0.917** |

On Figure 11, barplots confirm that ARGONet is the best model for all regions in term of correlation and error. On the same way, for the distribution of PCC and MSE (Figure 14 and 15), ARGONet is the best model.

Table 15 shows that the improvement obtained thanks to the ARGONet model compared to the autoregressive model is statistically significant for all regions for two-week ahead estimate. Depending on the region, ARGONet allows to reduce the error by 60% to 82%.

**Table 15.** Two-week ahead estimate - Relative efficiency being bigger than 1 suggests increased predictive power of ARGONet compared to the autoregressive model

| Region | Relative efficiency | 95% CI |
| --- | --- | --- |
| Auv. | 2.56 | [2.39;2.73] |
| Bour. | 5.49 | [4.81;6.19] |
| Bre. | 3.30 | [2.99;3.62] |
| Cen. | 3.33 | [3.07;3.59] |
| Gd Est | 4.00 | [3.55;4.45] |
| Ht Fra. | 3.00 | [2.79;3.22] |
| Ile Fra. | 2.84 | [2.57;3.11] |
| Norm. | 2.74 | [2.50;2.98] |
| Aqui. | 3.29 | [2.95;3.64] |
| Occi. | 4.34 | [4.02;4.66] |
| Loi. | 2.48 | [2.19;2.77] |
| Pro. | 5.22 | [4.57;5.86] |

Figure 12 shows two-week ahead estimates for the region Nouvelle-Aquitaine. As for one-week ahead estimate, we can see that estimates obtained with AR(52) and ARGO models are still delayed. It is not the case for Net and ARGONet models. Nevertheless, unlike ARGONet model, Net model tends to overestimate the peaks. On the heatmap Figure 13, we can see that ARGO model uses mostly 9 variables, including 6 variables from Google Data, one variable from Climatic Data, two variables from Historical data.

## Discussion

We have introduced a machine learning ensemble methodology that combines multiple data sources and multiple statistical approaches to accurately track flu activity in the 12 continental regions of France. To the best of our knowledge, this is a spatial resolution for which no forecasting approaches have been explored before in France. Our methodology provides real-time estimates as well as one- and two-week ahead forecasts.

The success of our approach comes from the ability to dynamically identify the appropriate method and data sources to produce the best disease activity estimates for a given location and time horizon in a prospective way (out-of-sample). Specifically, we show that the ARGO model alone (one that does not incorporate flu activity from neighboring regions) yields accurate results for real-time estimates but fails to produce optimal predictions for longer-term time-horizons. We find that the Net model (one that leverages information from neighboring regions alone) leads to reasonable flu predictions but tends to overestimate epidemic peaks. The proposed ensemble approach, named ARGONet (that combines information from both ARGO and the Net model), an extension of a model proposed in the USA [27], produces forecasts with the lowest errors and highest correlation as captured by Figure 1. This machine-learning ensemble approach displays both accuracy and robustness to estimate ILI activity up to two-weeks ahead of time at the french regional level.

Prediction error reductions are observed when using ARGONet over its autoregressive counterpart (up to 50% across regions) in real-time predictions. Whereas the prediction performance of ARGONet and ARGO are comparable (Table 10) in this same task. As the time-horizon of prediction increases, the improvements of predictions are more evident, leading to up to 80% error reductions when comparing ARGONet with AR, and up to 60% error reduction of ARGONet over ARGO for one-week ahead predictions; and up to 80% (ARGONet vs AR) and 30% (ARGONet vs ARGO) respectively in two-week ahead predictions (Tables 11 and 12). Figures S2 through S13 show these results graphically. As expected, autoregressive approaches show “within-range” prediction values that consistently lag behind the observed disease activity and lead to under-predictions close to peak activity.

We find that all external data sources contribute to improving local flu estimates, when compared to the baseline autoregressive model, specially for longer-term forecasts. Indeed, for the two-week ahead estimates, the combination of EHR data and Google data lead to correlation improvements of up to 25% and decreases in error of up to 60%. For Climatic data, this improvement reaches 20% for correlation and 25% for the error. For Twitter data, it reaches 20% for both correlation and error. By analyzing heatmaps (Figures 5, 9 and 13 and in the Supplementary materials) obtained for ARGO models, we can see that the contributions of different predictors (data sources) change over time and time-horizon of prediction, but all data sources appear to posses predictive power. Indeed, the most important data sources are EHR data and Google data in real-time and for longer-term forecasts. Historical data is consistently used in real-time, but less used for longer-term forecasting. Conversely, Climatic data and Twitter data are used more prominently for longer-term forecasts than for real-time estimate.

The fact that we could only access EHR data from Rennes University Hospital, and thus from the Brittany region, prevented us from being able to quantify the added valued of region-specific EHR information on flu predictions in their respective region. This should be evaluated in future research efforts. On the other hand, we find interesting the fact that data from a hospital in Rennes can improve flu forecasting in other regions.

Indeed, tables S4 to S6 show that forecasts that include Rennes’ EHR information, up to two weeks, are more accurate for all the regions when compared to the baseline autoregressive model. Rennes’ EHR data appears to be more relevant for some regions than others. For example, it appears to be an important predictor in the Brittany region (which contains Rennes) as expected, as well as in Normandy, which shares a border with Brittany. For Occitanie, Rennes’ EHR data improves predictions, which is in alignment with the fact that historical information shows that flu activity tends to occur synchronously (with a correlation of 0.93) as seen in Figure S1. We hypothesize that having access to region-specific EHR data, from all the french regions, will lead to prediction improvements across the board.

Twitter data was collected at the National level given the sparsity of relevant flu-related Tweets at the regional level. This was the case as we only had access to the publicly available data shared by Twitter’s API that only allows users to view up to 5% of all Geo-coded Tweets (themselves a small fraction of about 5% of the total corpus of all Tweets). We also suspect that gaining access to higher volumes of Tweets at the regional level could improve our forecasts.

For climatic data, we only had a access to weekly local temperature and precipitation. Future studies may explore incorporating other climatic indicators known to be more directly related to the transmission of the virus, such as humidity [28].

To conclude, we have shown that Internet-based data sources can yield accurate influenza estimates in the 12 continental regions in France. Operational implementations of these methods may prove to be useful for public health officials in the face of public health threats. Our regional-level flu estimates may contribute to better management of patients’ flow in general practitioners’ offices and in hospitals, particularly emergency departments.

## Data Availability

All data cannot be shared publicly, in particular EHR data, due to the protection of
patient data.

## Acknowledgments

We would like to thank the French National Research Agency for partially funding this work inside the Integrating and Sharing Health Data for Research Project (Grant No. ANR-15-CE19-0024). We also thank the French Sentinelles network and Google and Twitter services for making their data publicly available. MS and CP were partially funded by the National Institute of General Medical Sciences of the National Institutes of Health under Award Number R01GM130668. The content is solely the responsibility of the authors and does not necessarily represent the official views of the National Institutes of Health

## Authors Contribution

C.P. and M.S. conceived the research. C.P. wrote the manuscript with support from M.S.. G.B extracted hospital data. Y.H and T.B. extracted Twitter data. All authors discussed the results and contributed to the final manuscript.

## Supplementary Material

**Fig S1.**
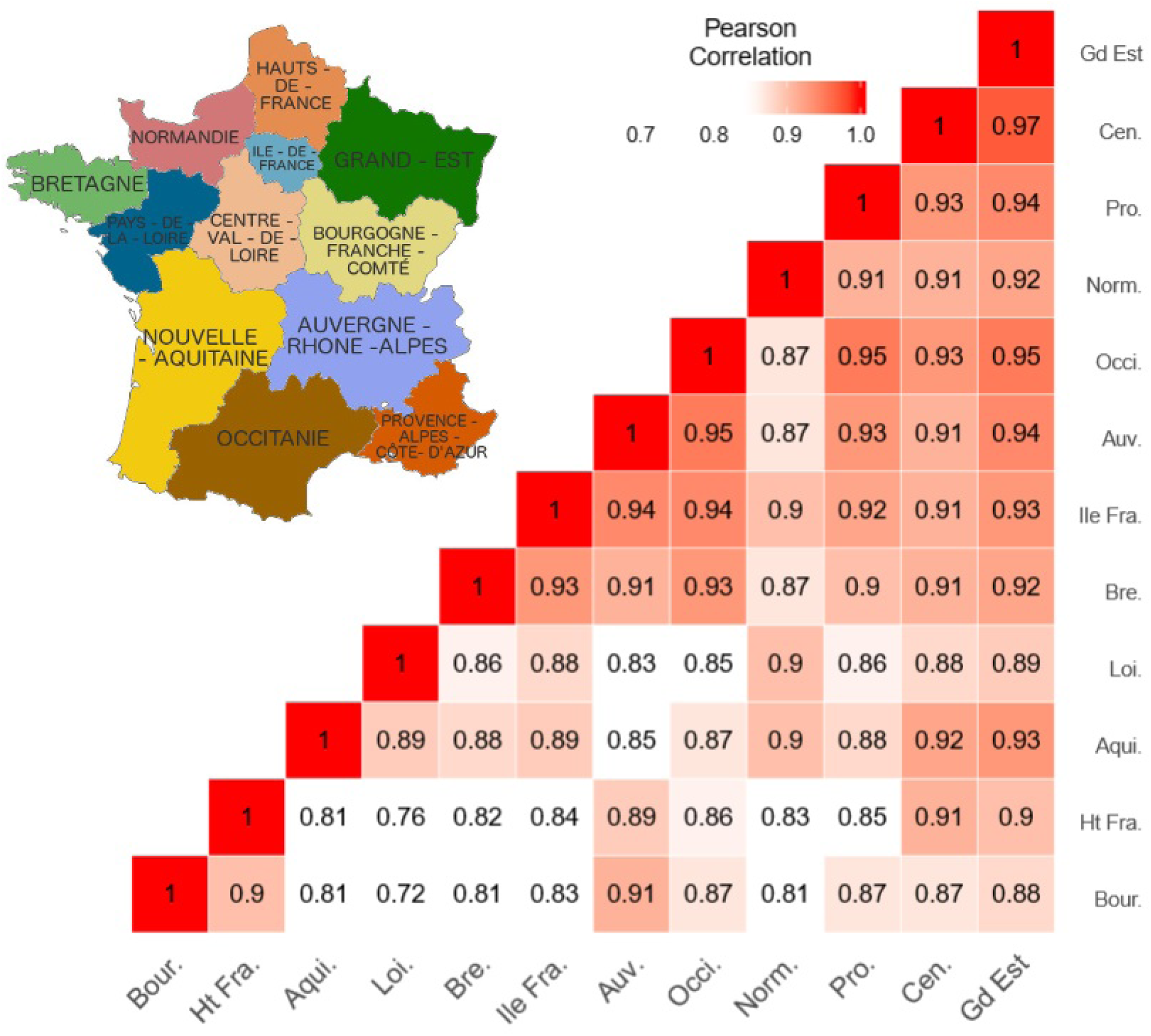
Correlation between French regions on the period starting from January 2013 to March 2017.

**Fig S2.**
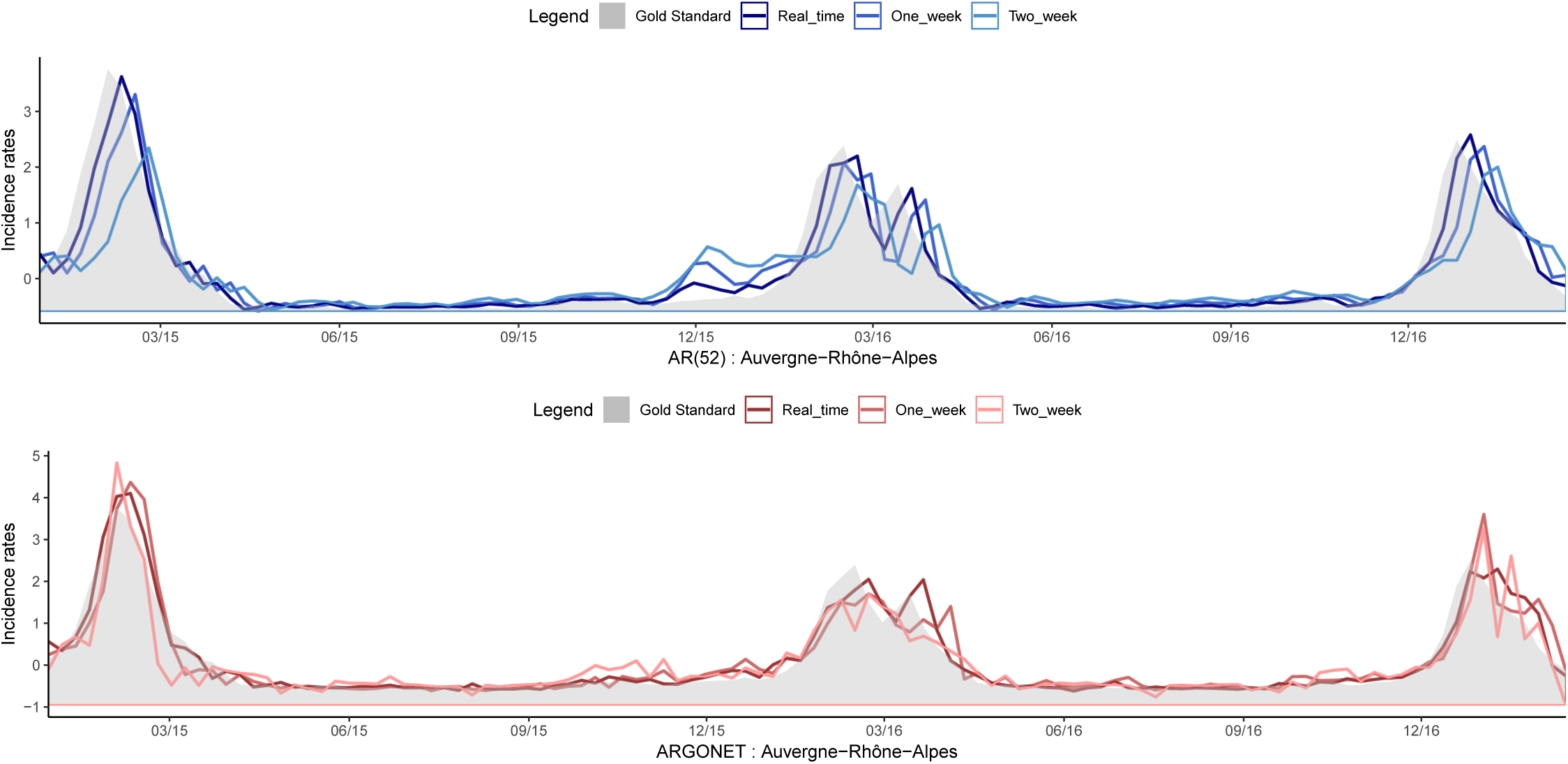
Evolution of Auvergne-Rhône-Alpes estimates over time for AR(52) and ARGONET models.

**Fig S3.**
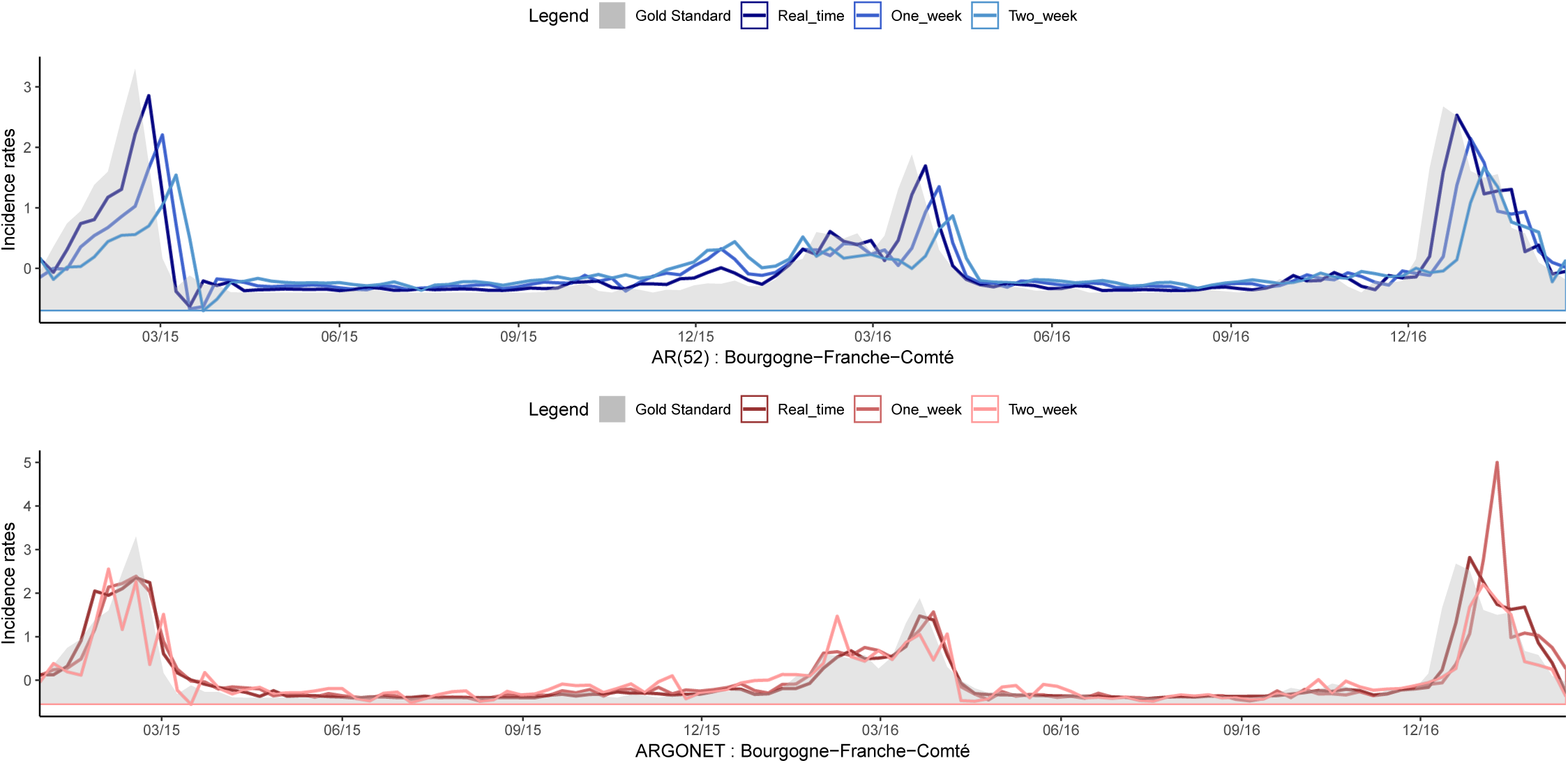
Evolution of Bourgogne-Franche-Comté estimates over time for AR(52) and ARGONET models.

**Fig S4.**
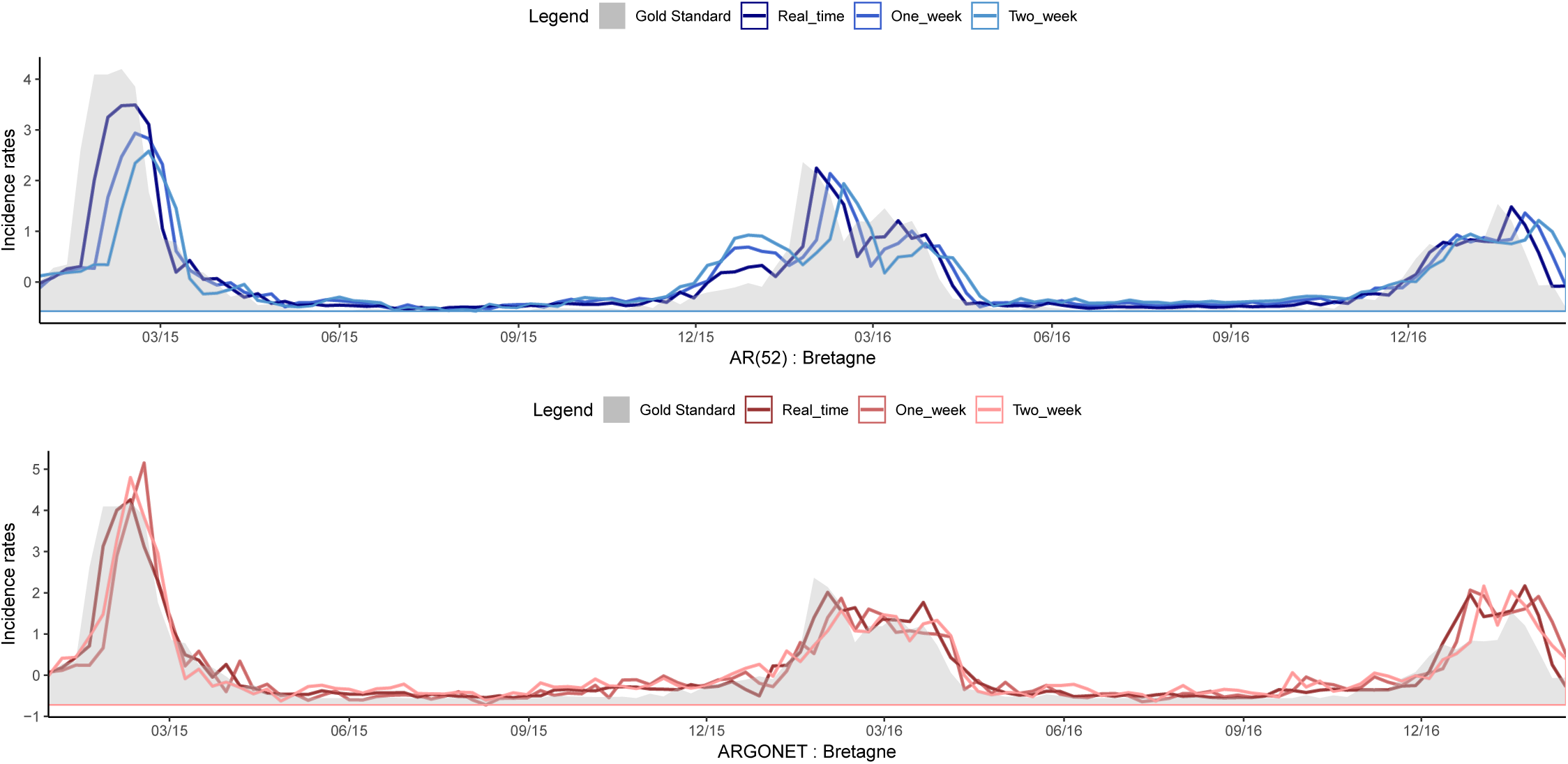
Evolution of Bretagne estimates over time for AR(52) and ARGONET models.

**Fig S5.**
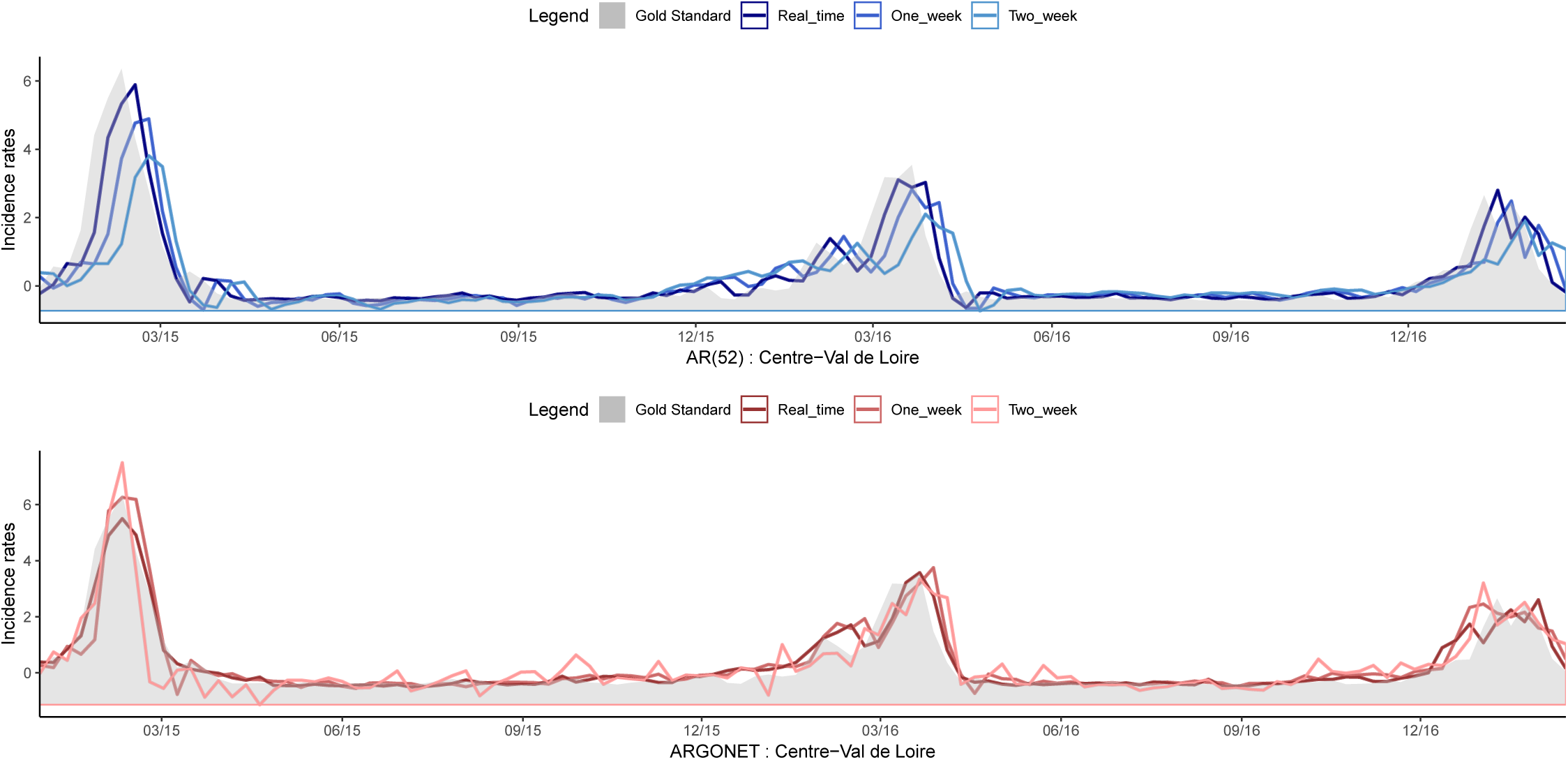
Evolution of Centre-Val de Loire estimates over time for AR(52) and ARGONET models.

**Fig S6.**
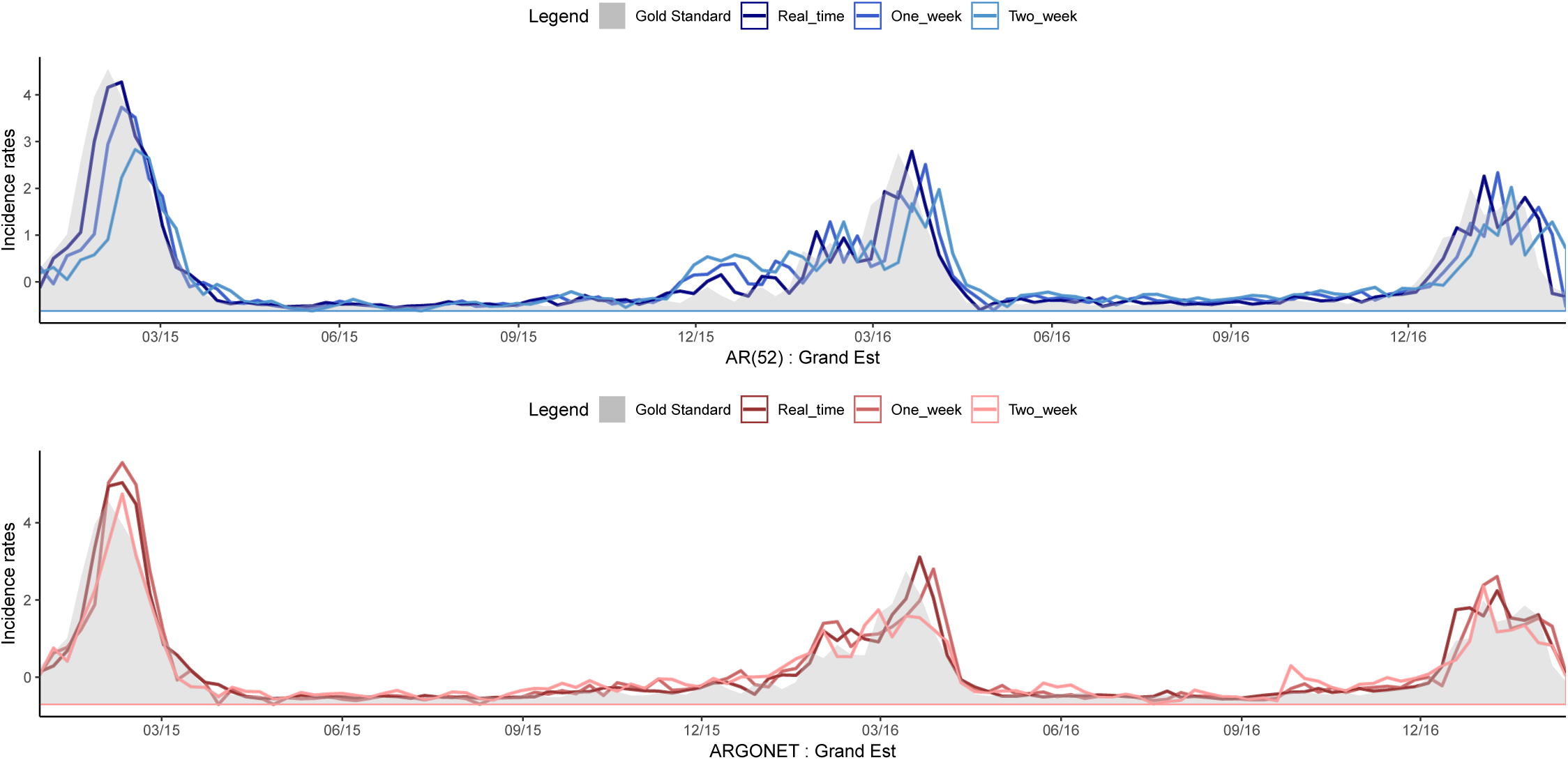
Evolution of Grand Est estimates over time for AR(52) and ARGONET models.

**Fig S7.**
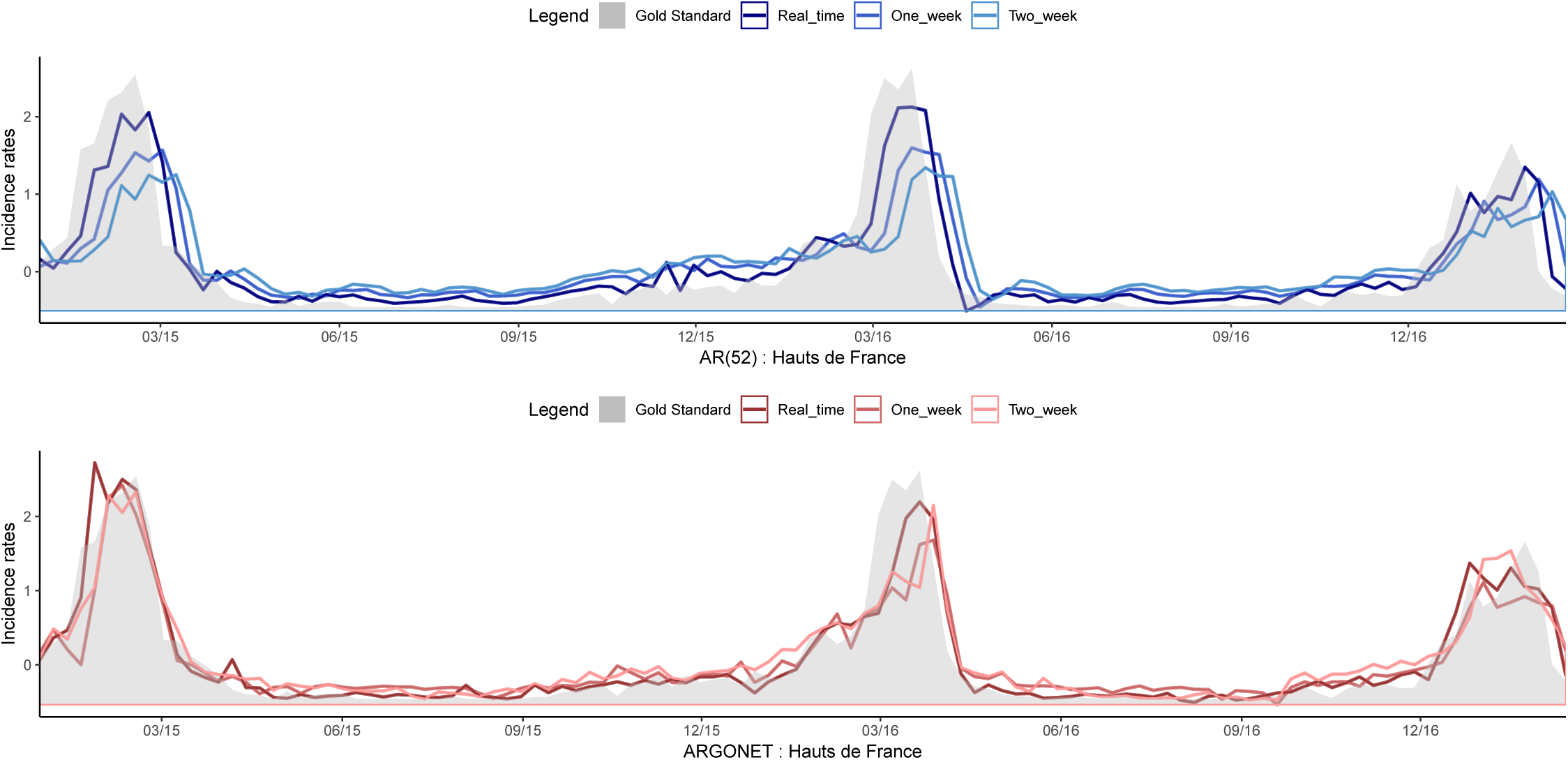
Evolution of Hauts de France estimates over time for AR(52) and ARGONET models.

**Fig S8.**
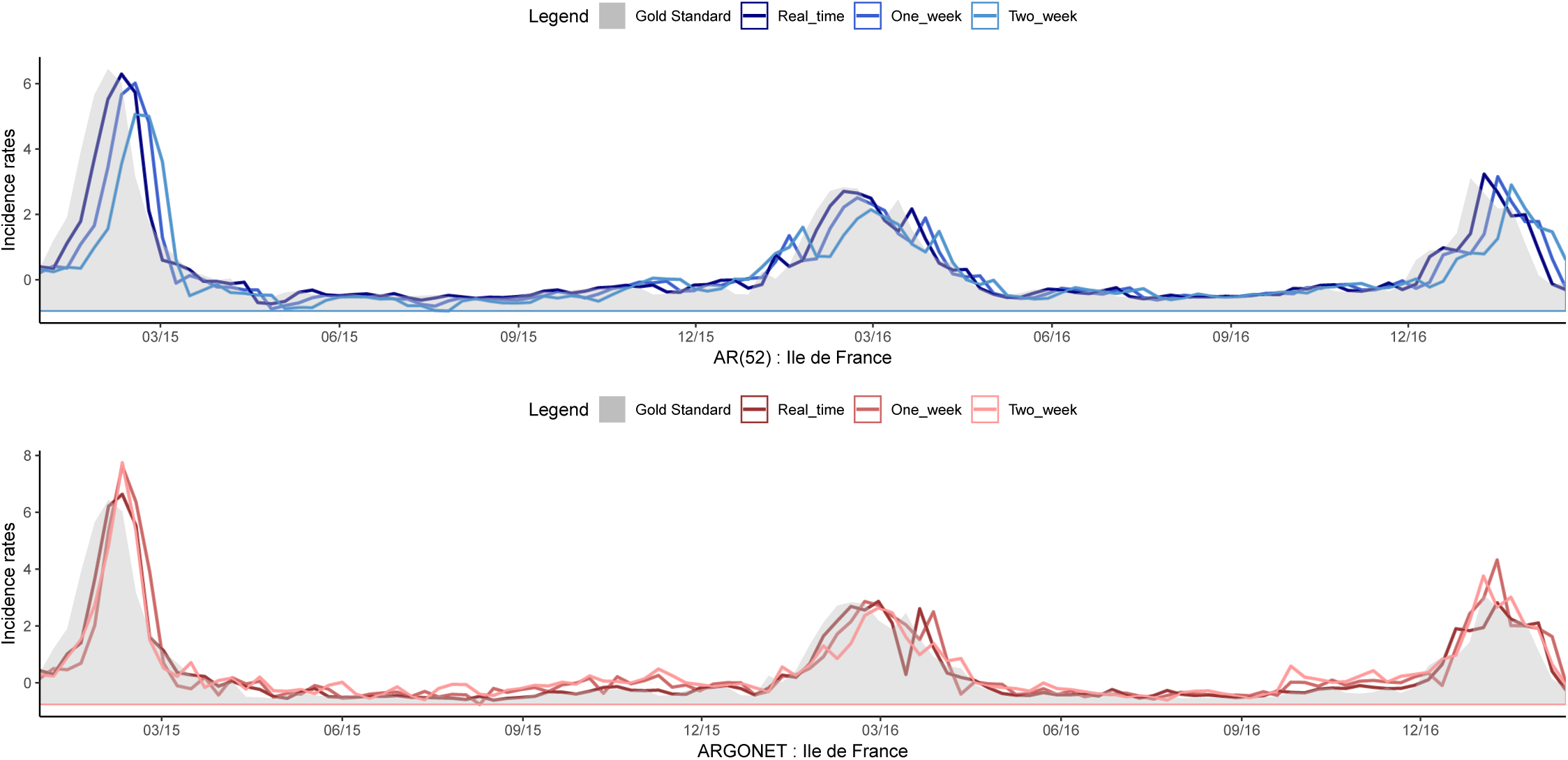
Evolution of Ile de France estimates over time for AR(52) and ARGONET models.

**Fig S9.**
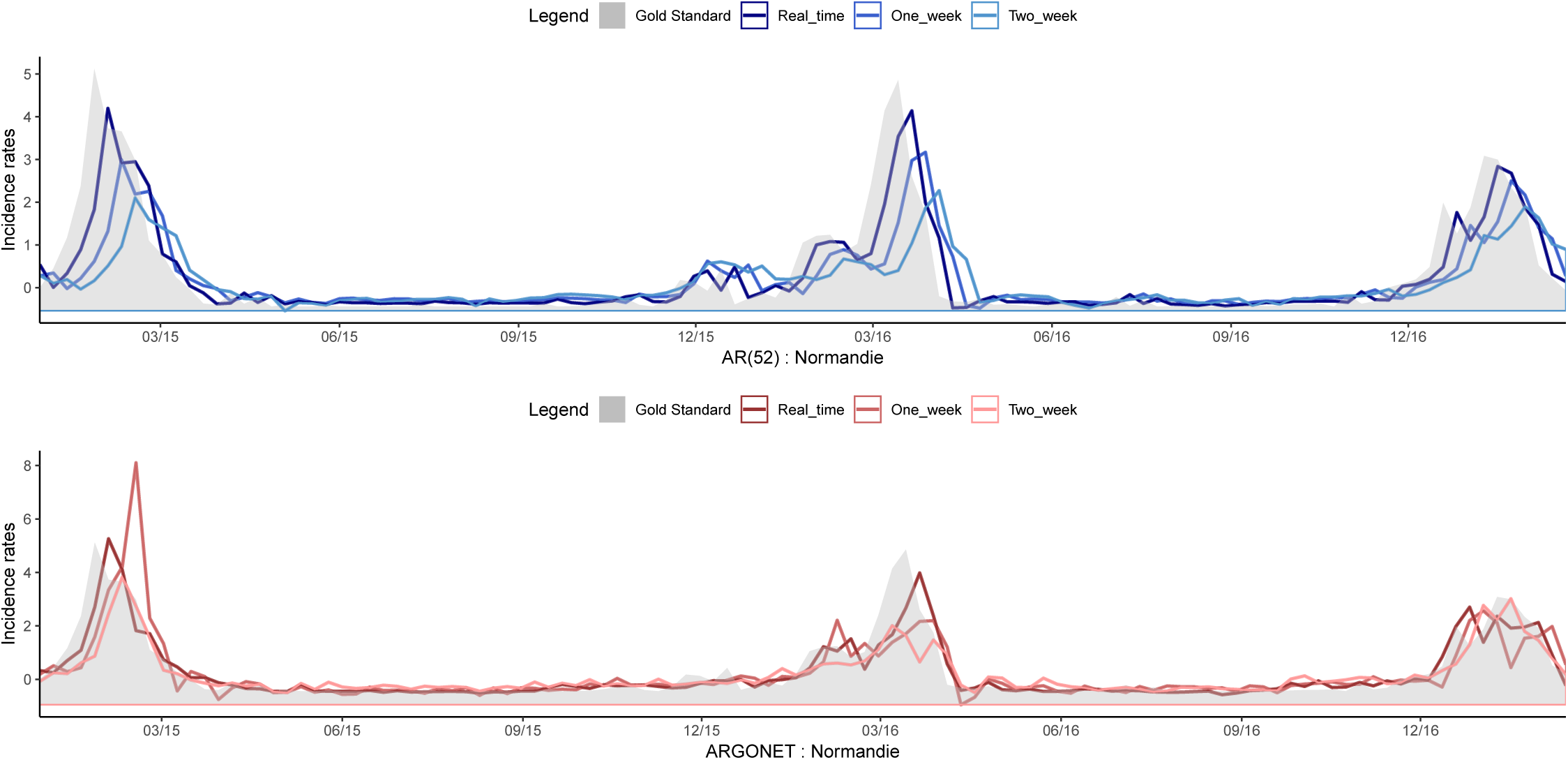
Evolution of Normandie estimates over time for AR(52) and ARGONet models.

**Fig S10.**
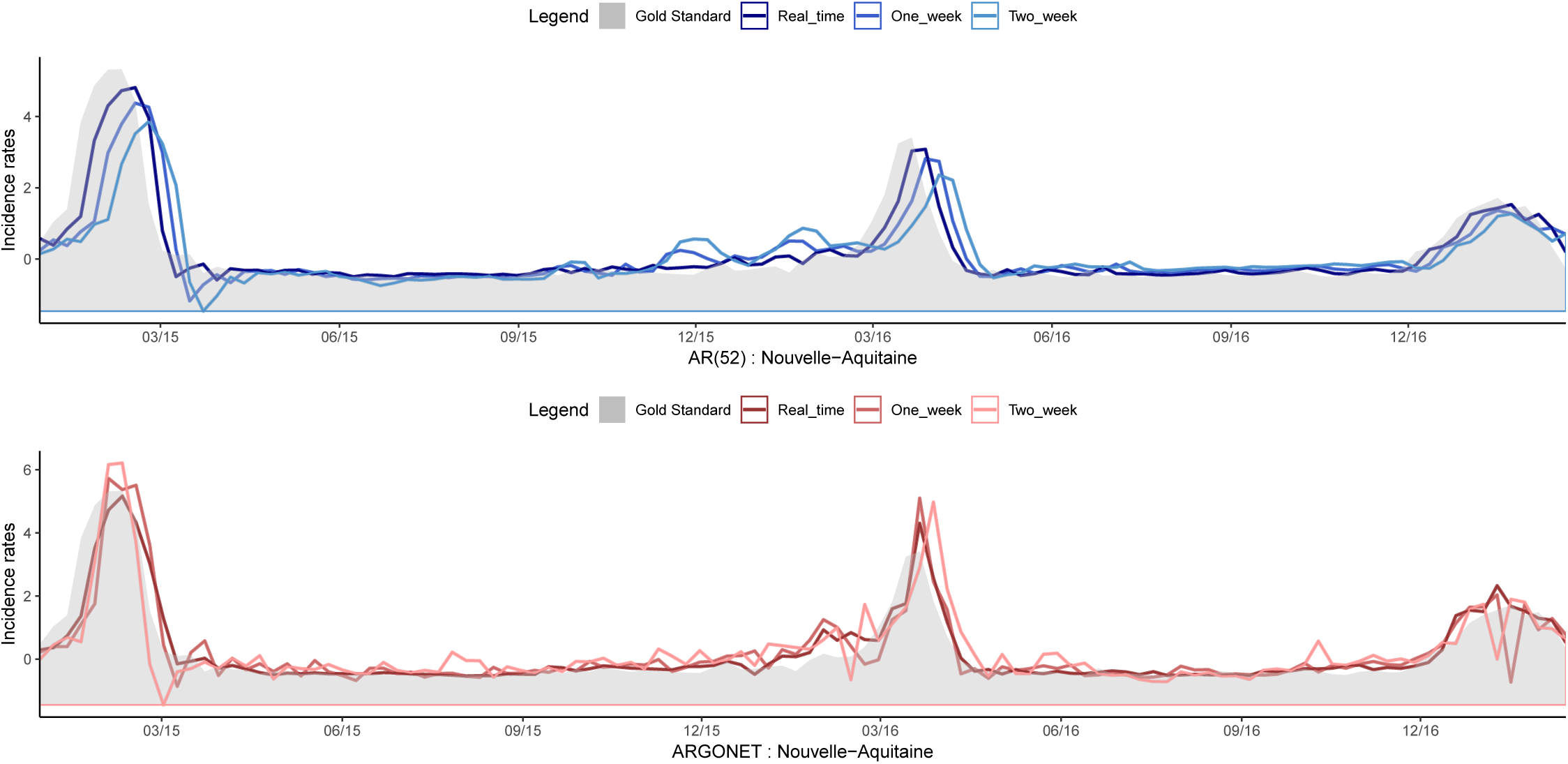
Evolution of Nouvelle-Aquitaine estimates over time for AR(52) and ARGONet models.

**Fig S11.**
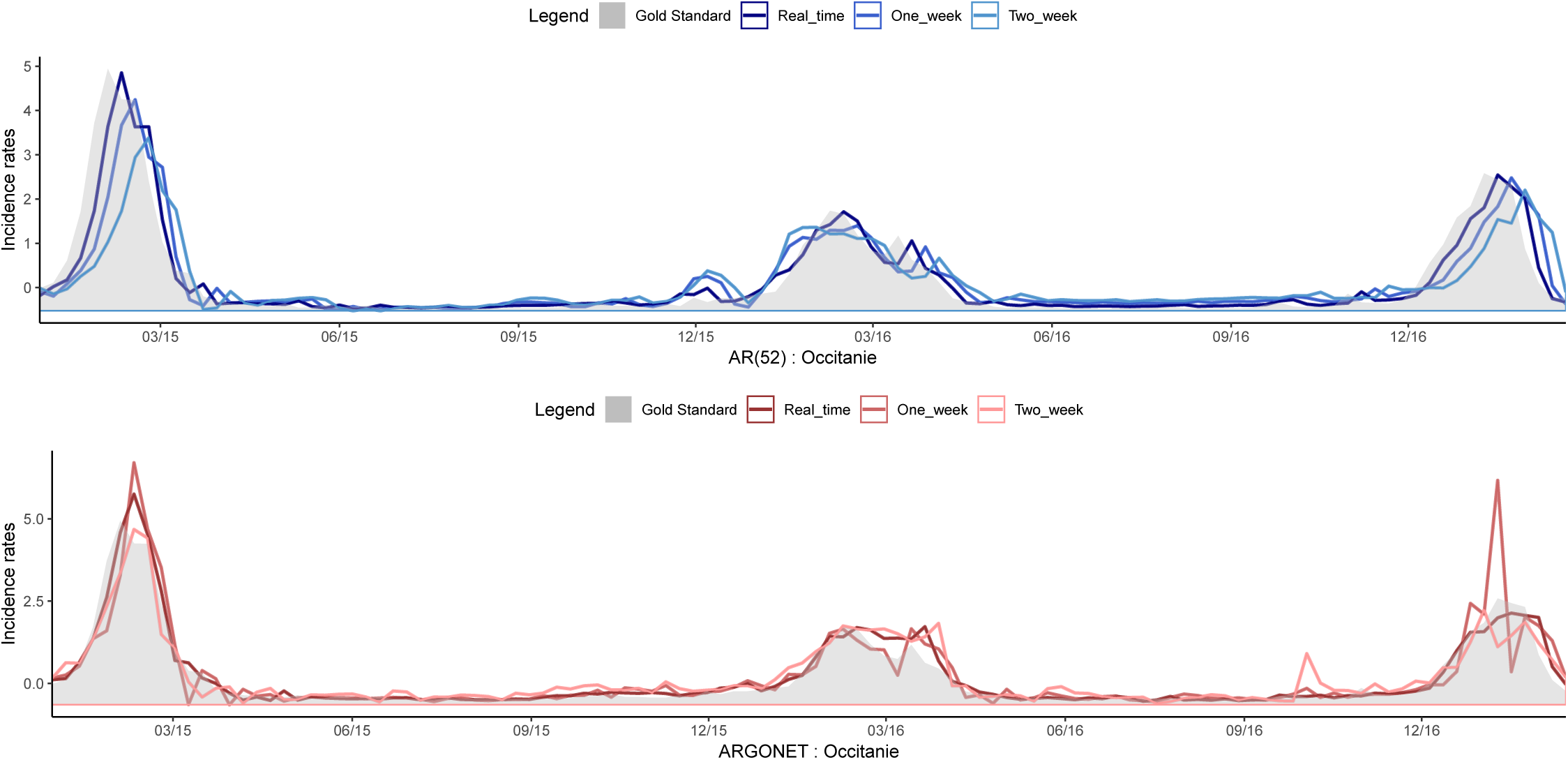
Evolution of Occitanie estimates over time for AR(52) and ARGONet models.

**Fig S12.**
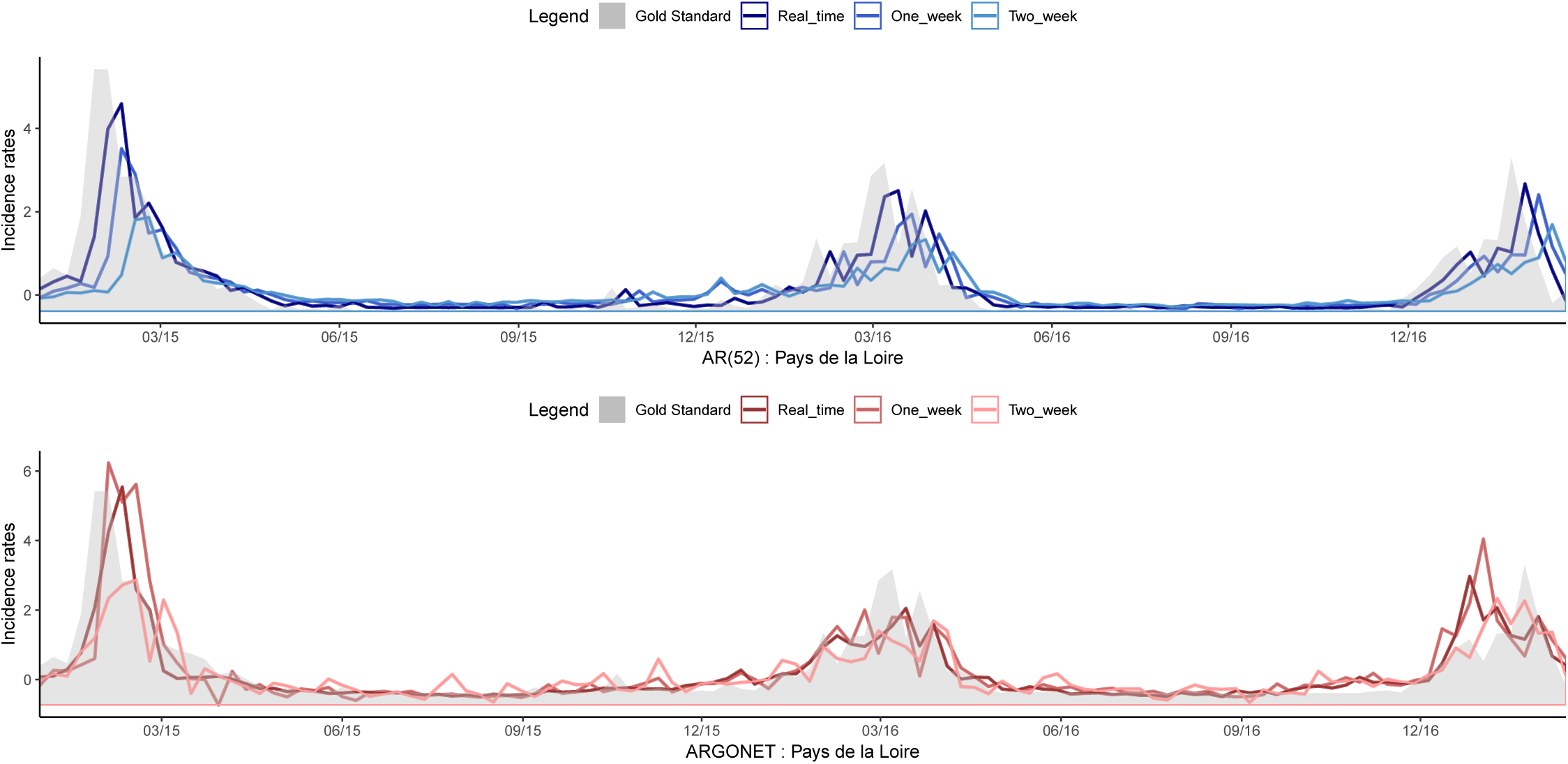
Evolution of Pays de la Loire estimates over time for AR(52) and ARGONet models.

**Fig S13.**
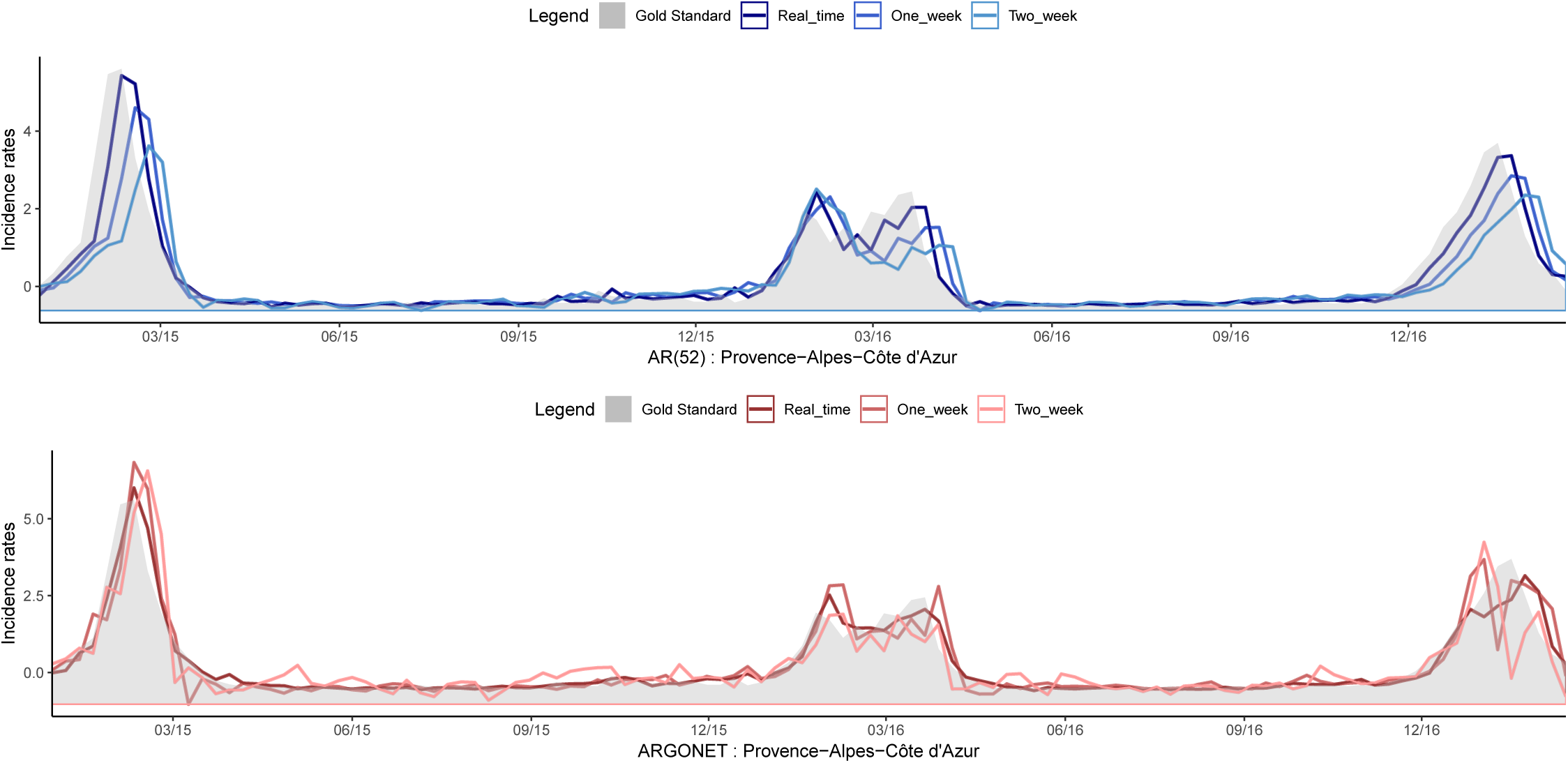
Evolution of Provence-Alpes-Côte d’Azur estimates over time for AR(52) and ARGONet models.

**Table S1.** Real time estimate: MSE and PCC for ARGO models including only historical data (AR(52)) and the 10 most correlated variables from Google data, for the period starting from January 2015 to March 2017

|  | Auv. | Bour. | Bre. | Cen. | Gd Est | Ht Fra. | Ile Fra. | Norm. | Aqui. | Occi. | Loi. | Pro. |
| --- | --- | --- | --- | --- | --- | --- | --- | --- | --- | --- | --- | --- |
| **MSE** |  |  |  |  |  |  |  |  |  |  |  |  |
| AR(52) | 0.083 | 0.203 | 0.167 | 0.161 | 0.096 | 0.169 | 0.129 | 0.241 | 0.161 | 0.108 | 0.368 | 0.141 |
| ARGO | **0.064** | **0.140** | **0.117** | **0.080** | **0.065** | **0.130** | **0.101** | **0.176** | **0.112** | **0.047** | **0.320** | **0.072** |
| **PCC** |  |  |  |  |  |  |  |  |  |  |  |  |
| AR(52) | 0.958 | 0.898 | 0.916 | 0.919 | 0.952 | 0.915 | 0.935 | 0.879 | 0.919 | 0.946 | 0.815 | 0.929 |
| ARGO | **0.967** | **0.929** | **0.941** | **0.959** | **0.967** | **0.935** | **0.949** | **0.911** | **0.944** | **0.976** | **0.839** | **0.964** |

**Table S2.** One-week ahead forecast : MSE and PCC for ARGO models including only historical data (AR(52)) and the 10 most correlated variables from Google data, for the period starting from January 2015 to March 2017

|  | Auv. | Bour. | Bre. | Cen. | Gd Est | Ht Fra. | Ile Fra. | Norm. | Aqui. | Occi. | Loi. | Pro. |
| --- | --- | --- | --- | --- | --- | --- | --- | --- | --- | --- | --- | --- |
| **MSE** |  |  |  |  |  |  |  |  |  |  |  |  |
| AR(52) | 0.262 | 0.611 | 0.433 | 0.438 | 0.276 | 0.433 | 0.372 | 0.580 | 0.480 | 0.278 | 0.704 | 0.389 |
| ARGO | **0.218** | **0.333** | **0.231** | **0.193** | **0.172** | **0.248** | **0.303** | **0.373** | **0.328** | **0.150** | **0.569** | **0.170** |
| **PCC** |  |  |  |  |  |  |  |  |  |  |  |  |
| AR(52) | 0.868 | 0.692 | 0.782 | 0.779 | 0.861 | 0.782 | 0.813 | 0.707 | 0.758 | 0.860 | 0.645 | 0.804 |
| ARGO | **0.890** | **0.832** | **0.884** | **0.903** | **0.913** | **0.875** | **0.847** | **0.812** | **0.835** | **0.924** | **0.713** | **0.914** |

**Table S3.** Two-week ahead forecast : MSE and PCC for ARGO models including only historical data (AR(52)) and the 10 most correlated variables from Google data, for the period starting from January 2015 to March 2017

|  | Auv. | Bour. | Bre. | Cen. | Gd Est | Ht Fra. | Ile Fra. | Norm. | Aqui. | Occi. | Loi. | Pro. |
| --- | --- | --- | --- | --- | --- | --- | --- | --- | --- | --- | --- | --- |
| **MSE** |  |  |  |  |  |  |  |  |  |  |  |  |
| AR(52) | 0.534 | 0.947 | 0.665 | 0.776 | 0.530 | 0.679 | 0.618 | 0.914 | 0.775 | 0.533 | 0.935 | 0.596 |
| ARGO | **0.370** | **0.600** | **0.445** | **0.332** | **0.330** | **0.351** | **0.446** | **0.568** | **0.562** | **0.291** | **0.753** | **0.282** |
| **PCC** |  |  |  |  |  |  |  |  |  |  |  |  |
| AR(52) | 0.731 | 0.522 | 0.665 | 0.609 | 0.733 | 0.658 | 0.688 | 0.539 | 0.610 | 0.731 | 0.528 | 0.699 |
| ARGO | **0.814** | **0.697** | **0.776** | **0.832** | **0.834** | **0.823** | **0.775** | **0.714** | **0.716** | **0.853** | **0.620** | **0.858** |

**Table S4.** Real time estimate: MSE and PCC for ARGO models including only historical data (AR(52)) and the 10 most correlated variables from Hospital data, for the period starting from January 2015 to March 2017

|  | Auv. | Bour. | Bre. | Cen. | Gd Est | Ht Fra. | Ile Fra. | Norm. | Aqui. | Occi. | Loi. | Pro. |
| --- | --- | --- | --- | --- | --- | --- | --- | --- | --- | --- | --- | --- |
| **MSE** |  |  |  |  |  |  |  |  |  |  |  |  |
| AR(52) | 0.083 | 0.203 | 0.167 | 0.161 | 0.096 | 0.169 | 0.129 | 0.241 | 0.161 | 0.108 | 0.368 | 0.141 |
| ARGO | **0.072** | **0.171** | **0.114** | **0.133** | **0.093** | **0.154** | **0.127** | **0.199** | **0.157** | **0.074** | **0.291** | **0.118** |
| **PCC** |  |  |  |  |  |  |  |  |  |  |  |  |
| AR(52) | 0.958 | 0.898 | 0.916 | 0.919 | 0.952 | 0.915 | 0.935 | 0.879 | 0.919 | 0.946 | 0.815 | 0.929 |
| ARGO | **0.964** | **0.914** | **0.943** | **0.933** | **0.953** | **0.922** | **0.936** | **0.900** | **0.921** | **0.963** | **0.853** | **0.941** |

**Table S5.** One-week ahead forecast : MSE and PCC for ARGO models including only historical data (AR(52)) and the 10 most correlated variables from Hospital data, for the period starting from January 2015 to March 2017

|  | Auv. | Bour. | Bre. | Cen. | Gd Est | Ht Fra. | Ile Fra. | Norm. | Aqui. | Occi. | Loi. | Pro. |
| --- | --- | --- | --- | --- | --- | --- | --- | --- | --- | --- | --- | --- |
| **MSE** |  |  |  |  |  |  |  |  |  |  |  |  |
| AR(52) | 0.262 | 0.611 | 0.433 | 0.438 | **0.276** | 0.433 | 0.372 | 0.580 | 0.480 | 0.278 | 0.704 | 0.389 |
| ARGO | **0.234** | **0.440** | **0.315** | **0.375** | 0.284 | **0.342** | **0.365** | **0.441** | **0.432** | **0.193** | **0.626** | **0.326** |
| **PCC** |  |  |  |  |  |  |  |  |  |  |  |  |
| AR(52) | 0.868 | 0.692 | 0.782 | 0.779 | **0.861** | 0.782 | 0.813 | 0.707 | 0.758 | 0.860 | 0.645 | 0.804 |
| ARGO | **0.882** | **0.778** | **0.841** | **0.811** | 0.857 | **0.828** | **0.816** | **0.778** | **0.782** | **0.903** | **0.684** | **0.835** |

**Table S6.** Two-week ahead forecast : MSE and PCC for ARGO models including only historical data (AR(52)) and the 10 most correlated variables from Hospital data, for the period starting from January 2015 to March 2017

|  | Auv. | Bour. | Bre. | Cen. | Gd Est | Ht Fra. | Ile Fra. | Norm. | Aqui. | Occi. | Loi. | Pro. |
| --- | --- | --- | --- | --- | --- | --- | --- | --- | --- | --- | --- | --- |
| **MSE** |  |  |  |  |  |  |  |  |  |  |  |  |
| AR(52) | 0.534 | 0.947 | 0.665 | 0.776 | 0.530 | 0.679 | **0.618** | 0.914 | 0.775 | 0.533 | 0.935 | 0.596 |
| ARGO | **0.488** | **0.812** | **0.532** | **0.632** | **0.620** | **0.514** | 0.638 | **0.658** | **0.730** | **0.452** | **0.758** | **0.482** |
| **PCC** |  |  |  |  |  |  |  |  |  |  |  |  |
| AR(52) | 0.731 | 0.522 | 0.665 | 0.609 | 0.733 | 0.658 | **0.688** | 0.539 | 0.610 | 0.731 | 0.528 | 0.699 |
| ARGO | **0.754** | **0.590** | **0.732** | **0.681** | **0.687** | **0.741** | 0.678 | **0.668** | **0.632** | **0.772** | **0.618** | **0.757** |

**Table S7.** Real time estimate: MSE and PCC for ARGO models including only historical data (AR(52)) and only hospital and Google data (all variables), for the period starting from January 2015 to March 2017

|  | Auv. | Bour. | Bre. | Cen. | Gd Est | Ht Fra. | Ile Fra. | Norm. | Aqui. | Occi. | Loi. | Pro. |
| --- | --- | --- | --- | --- | --- | --- | --- | --- | --- | --- | --- | --- |
| **MSE** |  |  |  |  |  |  |  |  |  |  |  |  |
| AR(52) | **0.083** | 0.203 | 0.167 | 0.161 | 0.096 | **0.169** | **0.129** | 0.241 | 0.161 | 0.108 | 0.368 | 0.141 |
| ARGO | 0.100 | **0.168** | **0.134** | **0.110** | **0.087** | 0.242 | 0.133 | **0.213** | **0.141** | **0.097** | **0.315** | **0.117** |
| **PCC** |  |  |  |  |  |  |  |  |  |  |  |  |
| AR(52) | **0.958** | 0.898 | 0.916 | 0.919 | 0.952 | **0.915** | **0.935** | 0.879 | 0.919 | 0.946 | 0.815 | 0.929 |
| ARGO | 0.950 | **0.916** | **0.933** | **0.945** | **0.957** | 0.878 | 0.933 | **0.893** | **0.929** | **0.951** | **0.842** | **0.941** |

**Table S8.** One-week ahead forecast : MSE and PCC for ARGO models including only historical data (AR(52)) and only hospital and Google data (all variables), for the period starting from January 2015 to March 2017

|  | Auv. | Bour. | Bre. | Cen. | Gd Est | Ht Fra. | Ile Fra. | Norm. | Aqui. | Occi. | Loi. | Pro. |
| --- | --- | --- | --- | --- | --- | --- | --- | --- | --- | --- | --- | --- |
| **MSE** |  |  |  |  |  |  |  |  |  |  |  |  |
| AR(52) | **0.262** | **0.611** | 0.433 | 0.438 | 0.276 | **0.433** | **0.372** | 0.580 | 0.480 | **0.278** | 0.704 | 0.389 |
| ARGO | 0.275 | 0.691 | **0.372** | **0.288** | **0.206** | 0.489 | 0.373 | **0.500** | **0.280** | 0.350 | **0.523** | **0.304** |
| **PCC** |  |  |  |  |  |  |  |  |  |  |  |  |
| AR(52) | **0.868** | **0.692** | 0.782 | 0.779 | 0.861 | **0.782** | 0.813 | 0.707 | 0.758 | **0.860** | 0.645 | 0.804 |
| ARGO | 0.862 | 0.653 | **0.814** | **0.856** | **0.897** | 0.755 | 0.813 | **0.749** | **0.860** | 0.825 | **0.738** | **0.848** |

**Table S9.** Two-week ahead forecast : MSE and PCC for ARGO models including only historical data (AR(52)) and only hospital and Google data (all variables), for the period starting from January 2015 to March 2017

|  | Auv. | Bour. | Bre. | Cen. | Gd Est | Ht Fra. | Ile Fra. | Norm. | Aqui. | Occi. | Loi. | Pro. |
| --- | --- | --- | --- | --- | --- | --- | --- | --- | --- | --- | --- | --- |
| **MSE** |  |  |  |  |  |  |  |  |  |  |  |  |
| AR(52) | 0.534 | 0.947 | 0.665 | 0.776 | 0.530 | 0.679 | 0.618 | 0.914 | 0.775 | 0.533 | 0.935 | 0.596 |
| ARGO | **0.516** | **0.724** | **0.408** | **0.539** | **0.361** | **0.657** | **0.493** | **0.632** | **0.454** | **0.286** | **0.643** | **0.269** |
| **PCC** |  |  |  |  |  |  |  |  |  |  |  |  |
| AR(52) | 0.731 | 0.522 | 0.665 | 0.609 | 0.733 | 0.658 | 0.688 | 0.539 | 0.610 | 0.731 | 0.528 | 0.699 |
| ARGO | **0.741** | **0.637** | **0.795** | **0.730** | **0.819** | **0.670** | **0.753** | **0.683** | **0.772** | **0.857** | **0.678** | **0.865** |

**Table S10.** PCC and MSE for real time estimate for all french regions for the period starting from January 2015 to March 2017 with all the variables from Google and hospital data included in ARGO model

|  | Auv. | Bour. | Bre. | Cen. | Gd Est | Ht Fra. | Ile Fra. | Norm. | Aqui. | Occi. | Loi. | Pro. |
| --- | --- | --- | --- | --- | --- | --- | --- | --- | --- | --- | --- | --- |
| **MSE** |  |  |  |  |  |  |  |  |  |  |  |  |
| AR(52) | 0.083 | 0.203 | 0.167 | 0.161 | 0.096 | 0.169 | 0.129 | 0.241 | 0.161 | 0.108 | 0.368 | 0.141 |
| Argo | 0.069 | 0.129 | **0.090** | **0.088** | **0.065** | 0.152 | 0.122 | 0.219 | 0.125 | **0.060** | **0.309** | 0.105 |
| Net | 0.071 | 0.181 | 0.156 | 0.104 | 0.098 | 0.127 | 0.141 | 0.265 | 0.139 | 0.105 | 0.375 | 0.115 |
| K=1 | 0.075 | **0.120** | 0.102 | 0.098 | 0.072 | 0.127 | 0.116 | 0.247 | **0.118** | 0.068 | 0.357 | 0.133 |
| K=2 | 0.066 | **0.124** | **0.099** | 0.094 | 0.081 | 0.126 | **0.115** | **0.210** | 0.119 | 0.079 | 0.311 | 0.123 |
| K=3 | 0.081 | 0.137 | 0.104 | 0.093 | 0.073 | 0.119 | 0.117 | **0.208** | 0.123 | 0.088 | 0.314 | 0.131 |
| K=4 | 0.068 | 0.128 | 0.106 | 0.101 | 0.072 | 0.129 | 0.116 | 0.242 | 0.124 | 0.069 | **0.292** | 0.136 |
| Mean | **0.057** | 0.141 | 0.109 | **0.090** | 0.072 | **0.115** | **0.116** | 0.224 | **0.118** | 0.070 | 0.311 | **0.090** |
| Lm | **0.059** | 0.139 | 0.105 | 0.097 | **0.063** | **0.118** | 0.131 | 0.264 | 0.129 | **0.066** | 0.336 | **0.100** |
| **PCC** |  |  |  |  |  |  |  |  |  |  |  |  |
| AR(52) | 0.958 | 0.898 | 0.916 | 0.919 | 0.952 | 0.915 | 0.935 | 0.879 | 0.919 | 0.946 | 0.815 | 0.929 |
| Argo | 0.965 | 0.935 | **0.954** | **0.955** | **0.967** | 0.923 | 0.938 | 0.890 | 0.937 | **0.970** | **0.844** | 0.947 |
| Net | 0.964 | 0.909 | 0.921 | 0.948 | 0.951 | 0.936 | 0.929 | 0.866 | 0.930 | 0.947 | 0.811 | 0.942 |
| K=1 | 0.962 | **0.956** | 0.948 | 0.951 | 0.963 | 0.936 | 0.941 | 0.875 | **0.941** | 0.966 | 0.820 | 0.933 |
| K=2 | 0.967 | **0.939** | **0.950** | 0.952 | 0.959 | 0.936 | **0.942** | **0.894** | 0.940 | 0.960 | 0.843 | 0.938 |
| K=3 | 0.959 | 0.937 | 0.948 | 0.953 | 0.963 | 0.940 | 0.942 | **0.895** | 0.938 | 0.956 | 0.842 | 0.934 |
| K=4 | 0.966 | 0.931 | 0.947 | 0.949 | 0.964 | 0.935 | 0.941 | 0.877 | 0.938 | 0.965 | **0.852** | 0.931 |
| Mean | **0.971** | 0.929 | 0.945 | **0.955** | 0.964 | **0.942** | **0.942** | 0.887 | **0.941** | 0.965 | 0.843 | **0.954** |
| Lm | **0.970** | 0.930 | 0.947 | 0.951 | **0.968** | **0.940** | 0.942 | 0.867 | 0.935 | **0.967** | 0.830 | **0.949** |

**Table S11.** PCC and MSE for one-week forecast for all french regions for the period starting from January 2015 to March 2017 with all the variables from Google and hospital data included in ARGO model

|  | Auv. | Bour. | Bre. | Cen. | Gd Est | Ht Fra. | Ile Fra. | Norm. | Aqui. | Occi. | Loi. | Pro. |
| --- | --- | --- | --- | --- | --- | --- | --- | --- | --- | --- | --- | --- |
| **MSE** |  |  |  |  |  |  |  |  |  |  |  |  |
| AR(52) | 0.262 | 0.611 | 0.433 | 0.438 | 0.276 | 0.433 | 0.372 | 0.580 | 0.480 | 0.278 | 0.704 | 0.389 |
| Argo | 0.205 | 0.803 | 0.334 | 0.259 | 0.394 | 0.289 | 0.279 | 0.487 | 0.300 | 0.168 | 0.479 | 0.254 |
| Net | 0.343 | **0.269** | 0.398 | 0.282 | 0.271 | 0.324 | 0.439 | 0.595 | 0.518 | 0.235 | 0.770 | 0.173 |
| K=1 | 0.203 | 0.783 | **0.213** | 0.237 | 0.328 | 0.385 | 0.238 | 0.347 | **0.196** | 0.112 | **0.460** | 0.159 |
| K=2 | 0.253 | 0.766 | 0.227 | 0.167 | 0.320 | 0.333 | 0.249 | **0.338** | 0.212 | **0.101** | 0.486 | 0.173 |
| K=3 | 0.235 | **0.205** | 0.244 | 0.173 | 0.319 | 0.366 | 0.245 | 0.343 | **0.209** | 0.213 | **0.465** | 0.156 |
| K=4 | 0.187 | 0.765 | 0.339 | 0.189 | 0.314 | 0.374 | 0.254 | **0.331** | 0.227 | 0.155 | 0.479 | 0.180 |
| Mean | **0.101** | 0.435 | 0.217 | **0.156** | **0.172** | **0.188** | **0.170** | 0.444 | 0.252 | **0.098** | 0.523 | **0.084** |
| Lm | **0.118** | 0.327 | **0.216** | **0.165** | **0.182** | **0.226** | **0.219** | 0.484 | 0.284 | 0.103 | 0.536 | **0.083** |
| **PCC** |  |  |  |  |  |  |  |  |  |  |  |  |
| AR(52) | 0.868 | 0.692 | 0.782 | 0.779 | 0.861 | 0.782 | 0.813 | 0.707 | 0.758 | 0.860 | 0.645 | 0.804 |
| Argo | 0.896 | 0.595 | 0.832 | 0.869 | 0.801 | 0.854 | 0.859 | 0.754 | 0.849 | 0.915 | 0.758 | 0.876 |
| Net | 0.827 | **0.864** | 0.799 | 0.858 | 0.863 | 0.837 | 0.778 | 0.700 | 0.739 | 0.881 | 0.612 | 0.912 |
| K=1 | 0.897 | 0.605 | **0.893** | 0.881 | 0.835 | 0.806 | 0.880 | 0.825 | **0.901** | 0.944 | **0.768** | 0.920 |
| K=2 | 0.872 | 0.614 | 0.885 | 0.916 | 0.839 | 0.832 | 0.875 | **0.860** | 0.893 | **0.949** | 0.755 | 0.913 |
| K=3 | 0.881 | **0.897** | 0.877 | 0.913 | 0.839 | 0.815 | 0.877 | 0.827 | **0.895** | 0.893 | **0.765** | 0.921 |
| K=4 | 0.906 | 0.614 | 0.829 | 0.905 | 0.842 | 0.811 | 0.872 | **0.833** | 0.885 | 0.922 | 0.759 | 0.909 |
| Mean | **0.949** | 0.781 | 0.890 | **0.921** | **0.913** | **0.905** | **0.914** | 0.776 | 0.873 | **0.951** | 0.736 | **0.957** |
| Lm | **0.940** | 0.835 | **0.891** | **0.917** | **0.908** | **0.886** | **0.890** | 0.756 | 0.857 | 0.948 | 0.729 | **0.958** |

**Table S12.**
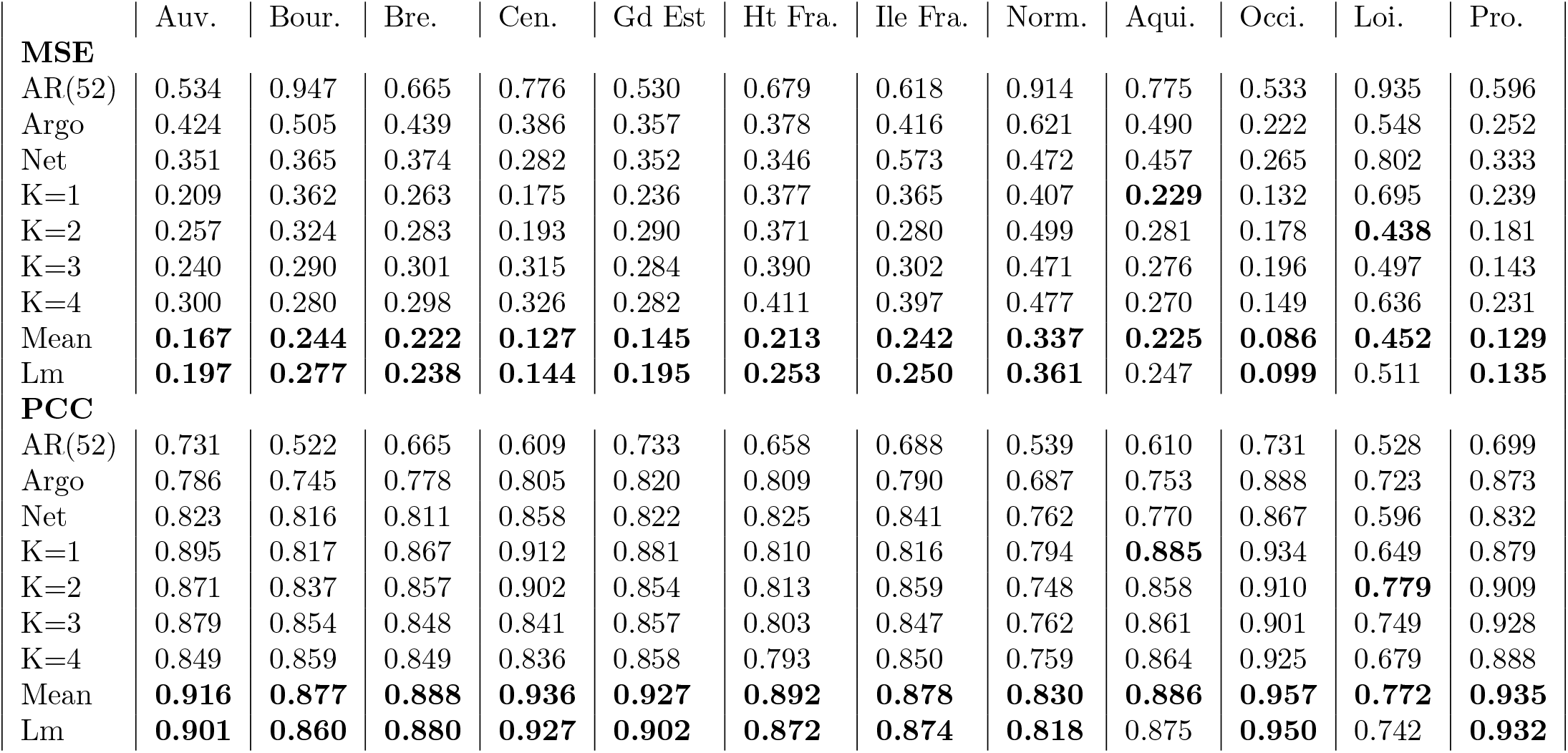
PCC and MSE for two-week forecast for all french regions for the period starting from January 2015 to March 2017 with all the variables from Google and hospital data included in ARGO model

**Fig S14.**
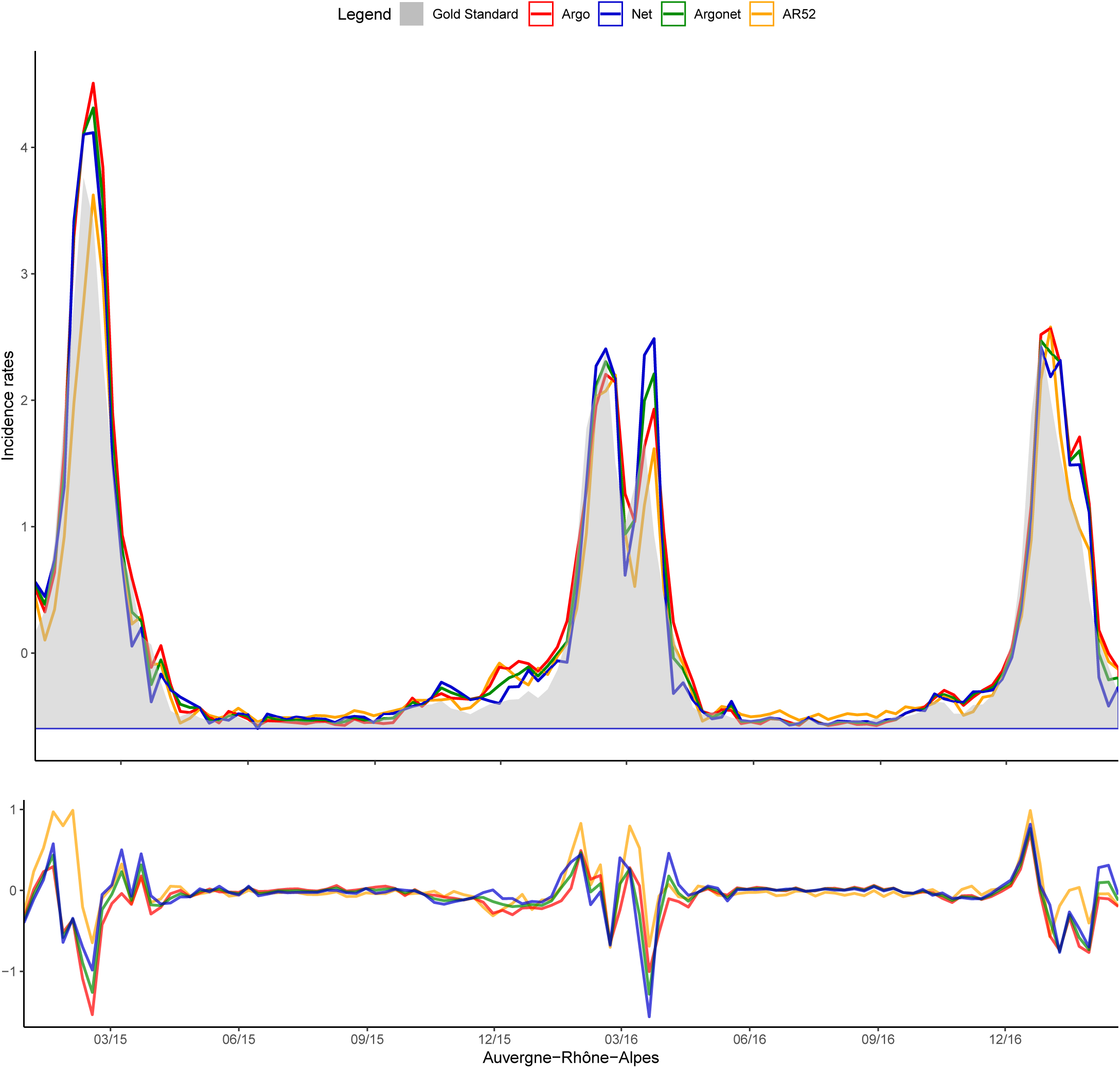
Auvergne Real-time estimate.

**Fig S15.**
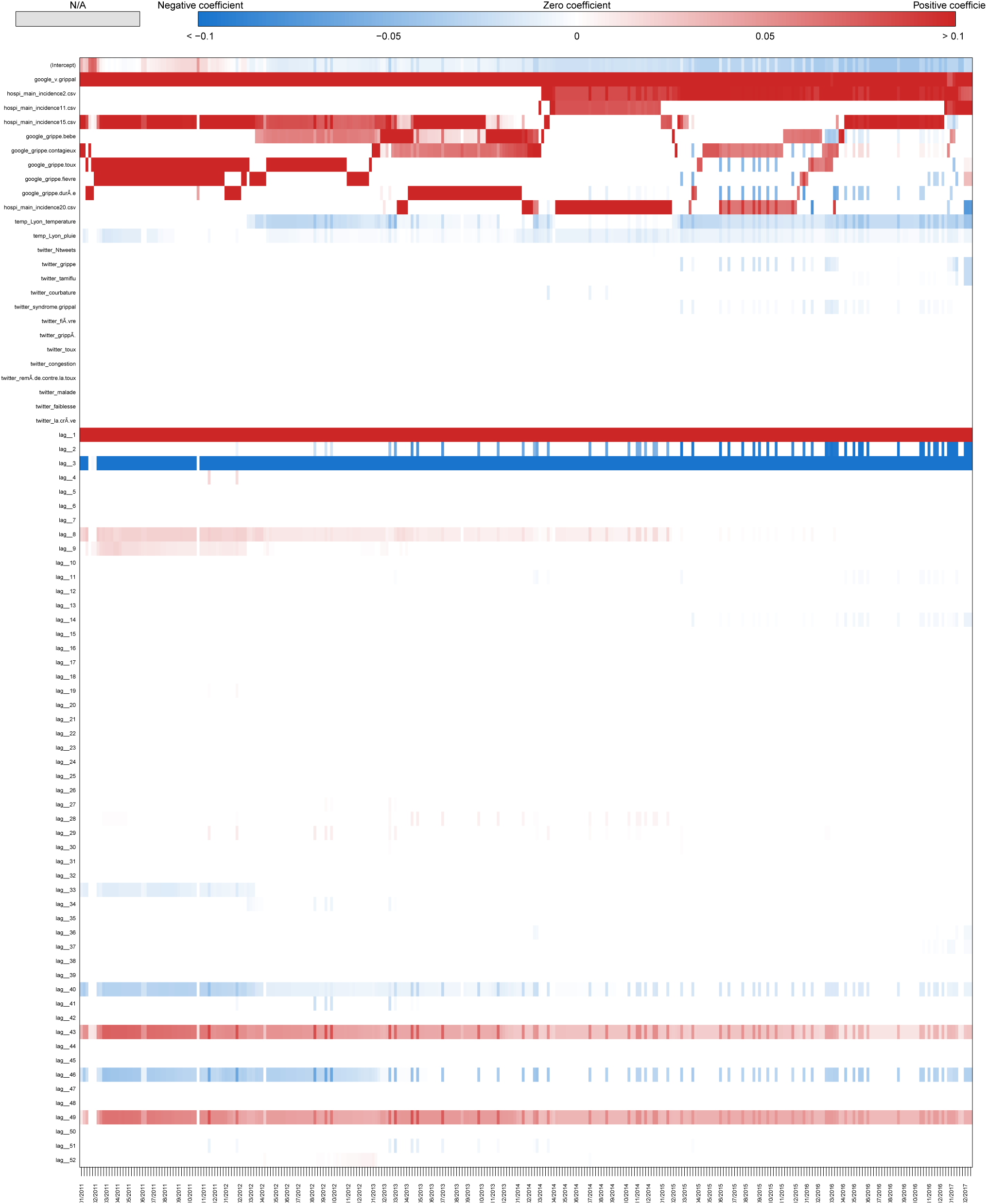
Coefficients Auvergne Real-time estimate.

**Fig S16.**
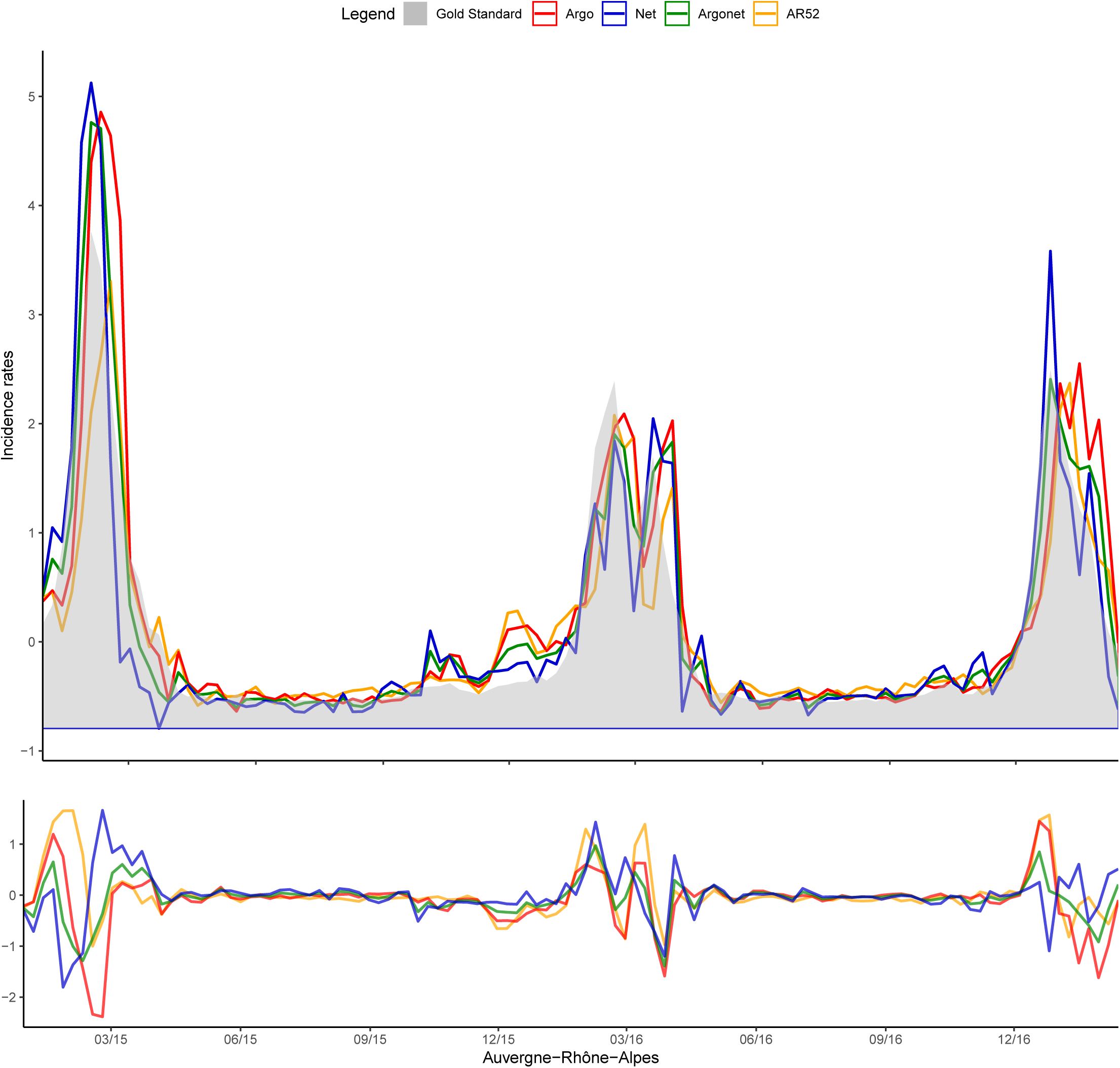
Auvergne One-week estimate.

**Fig S17.**
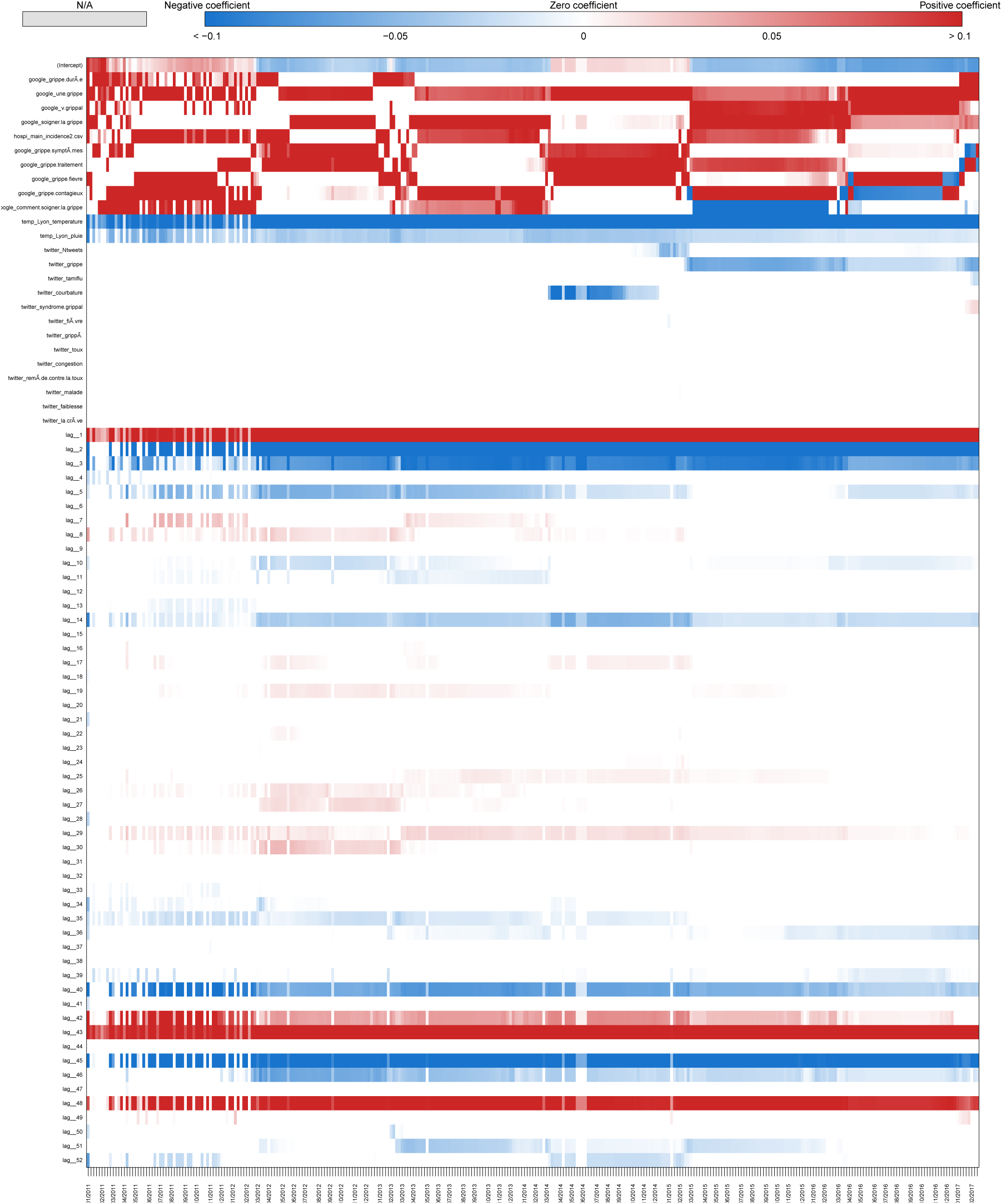
Coefficients Auvergne One-week estimate.

**Fig S18.**
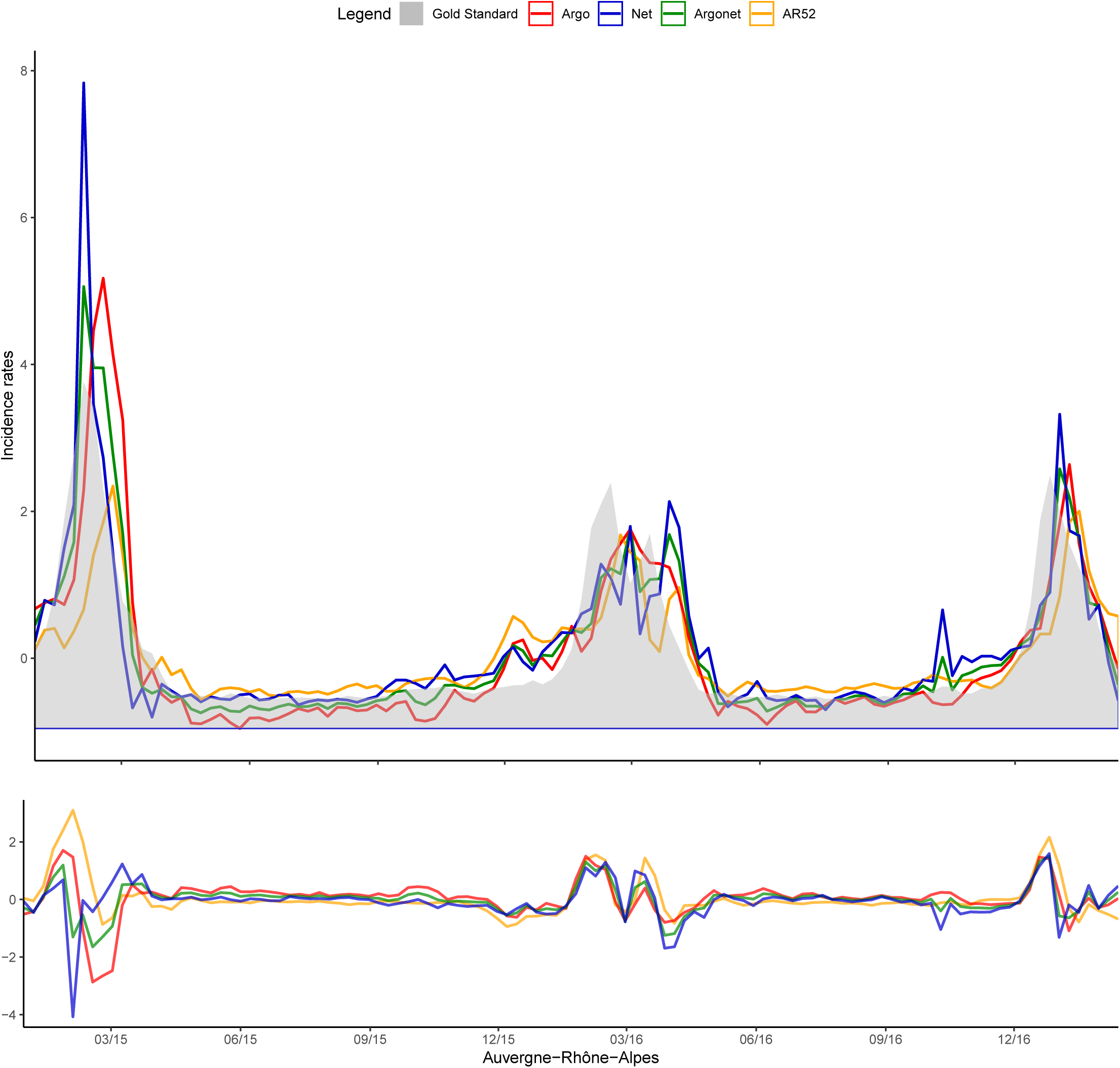
Auvergne Two-week estimate.

**Fig S19.**
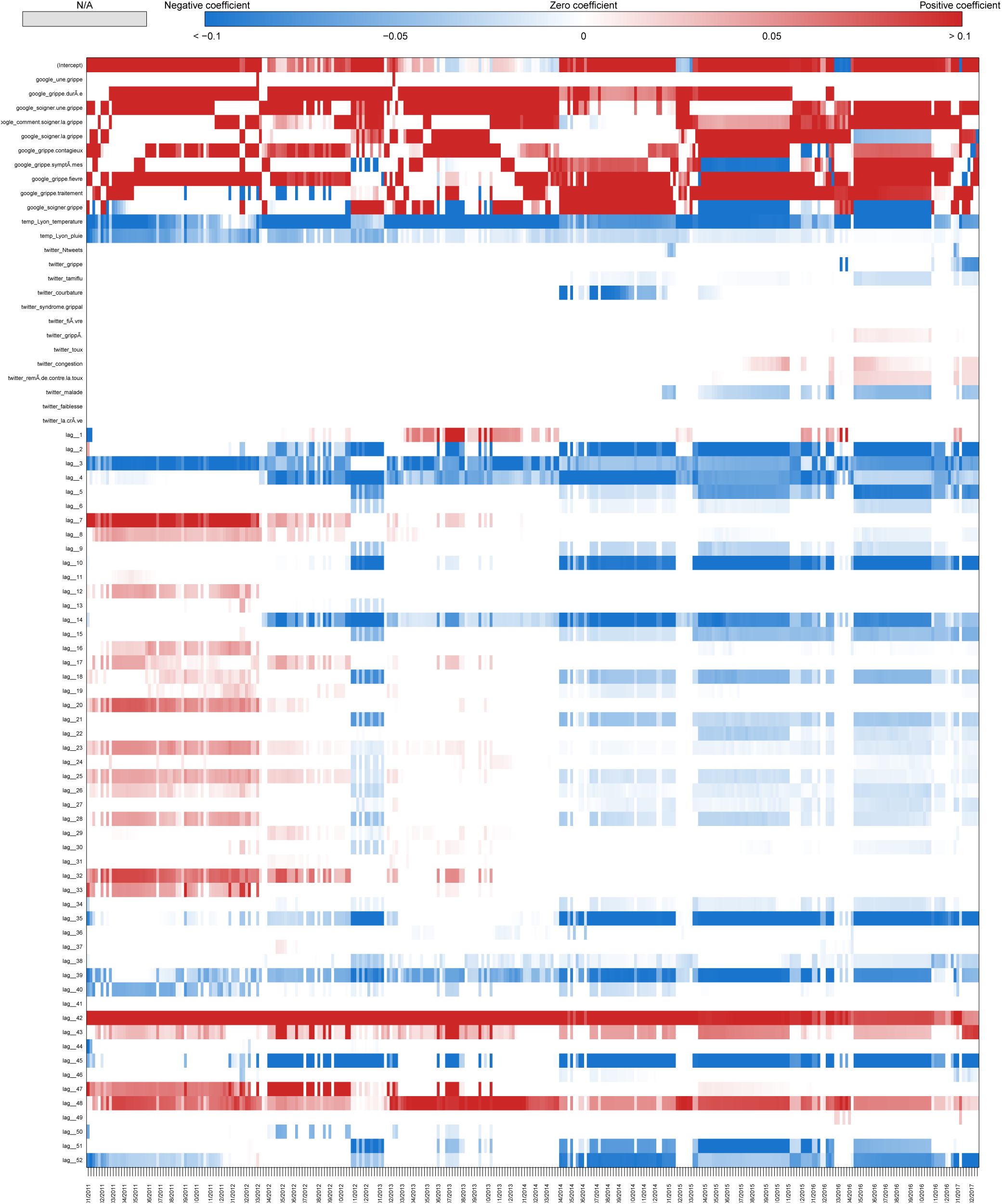
Coefficients Auvergne Two-week estimate.

**Fig S20.**
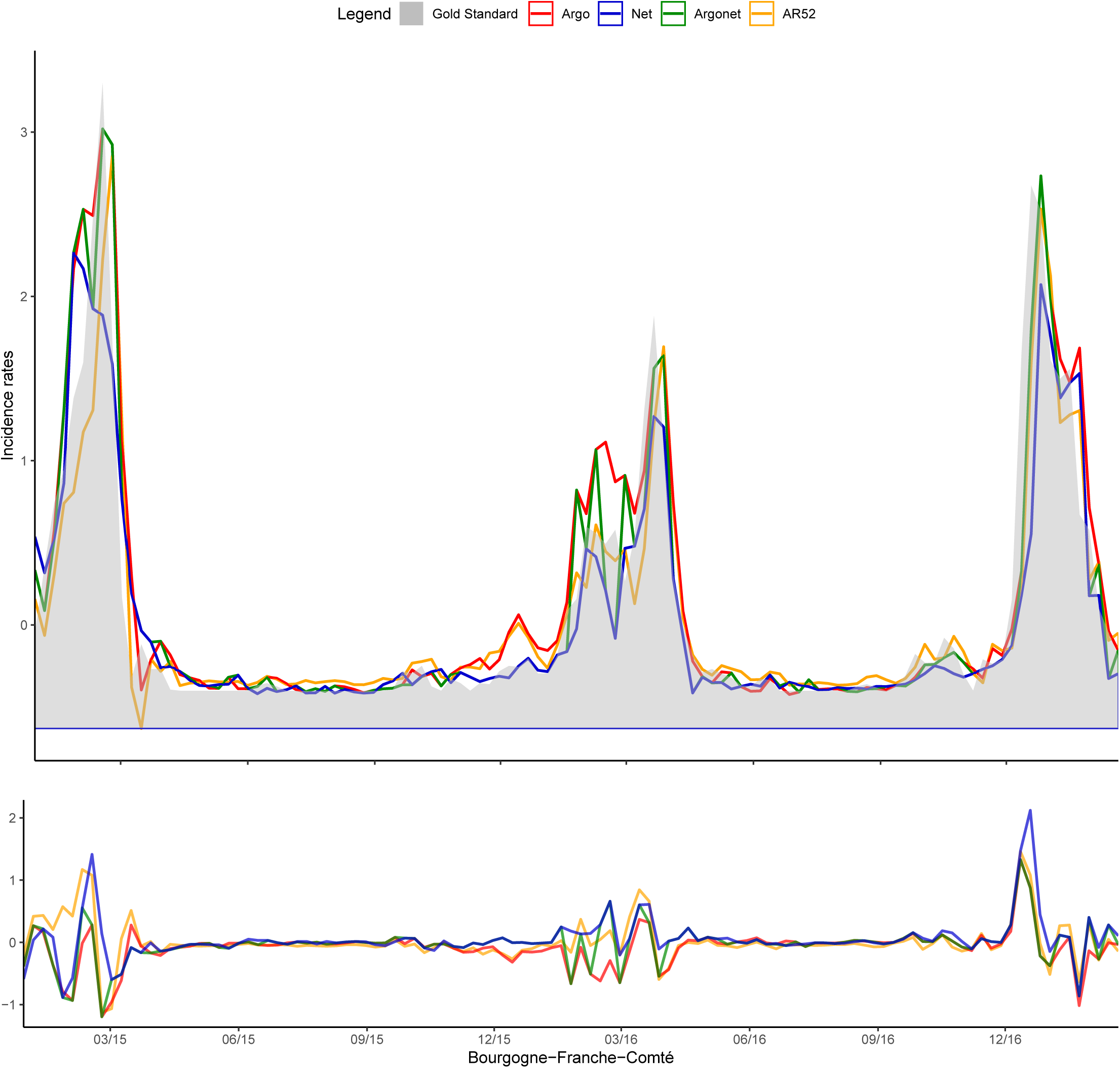
Bourgogne Franche Comté Real-time estimate.

**Fig S21.**
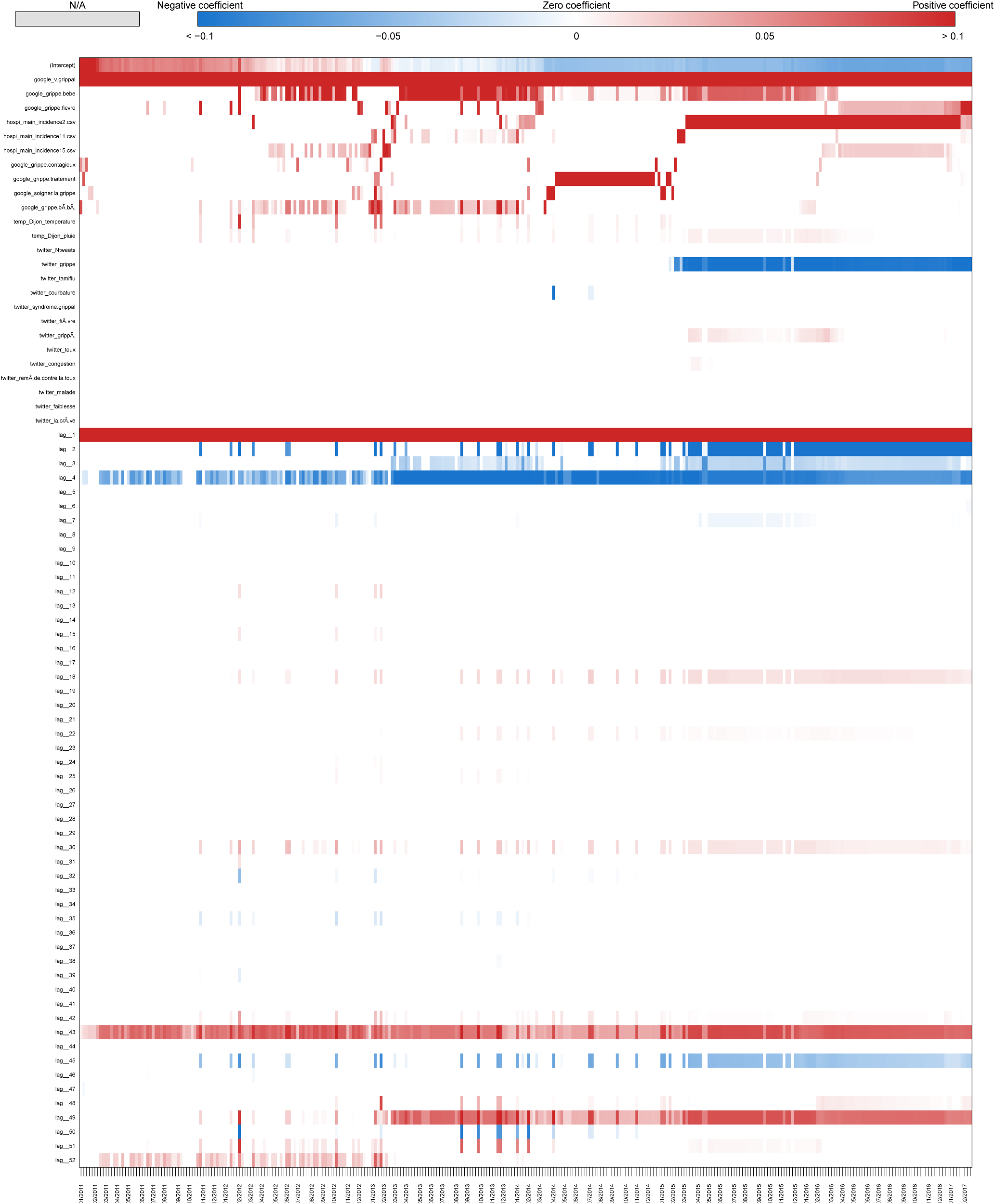
Coefficients Bourgogne Franche Comté Real-time estimate.

**Fig S22.**
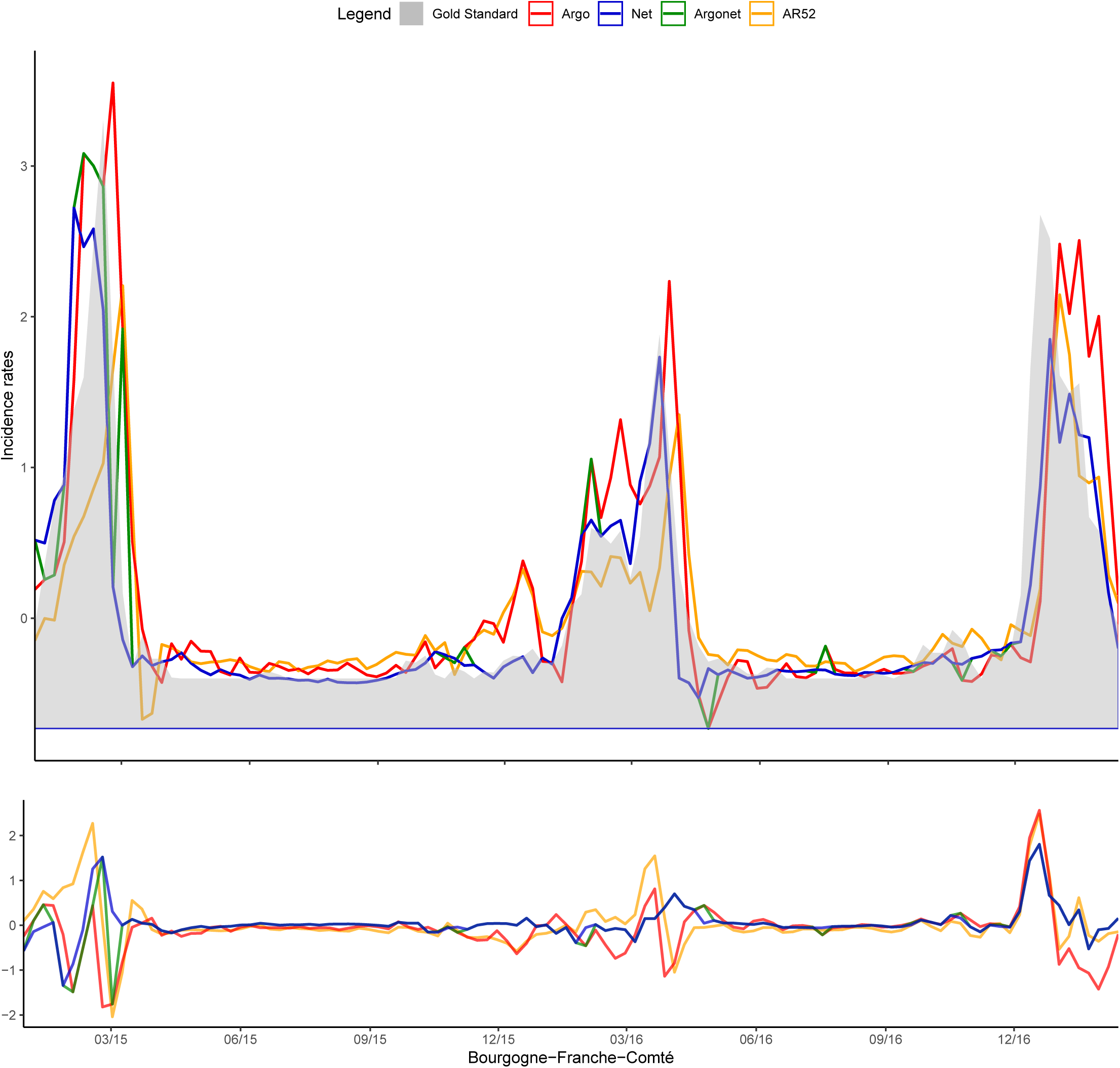
Bourgogne One-week estimate.

**Fig S23.**
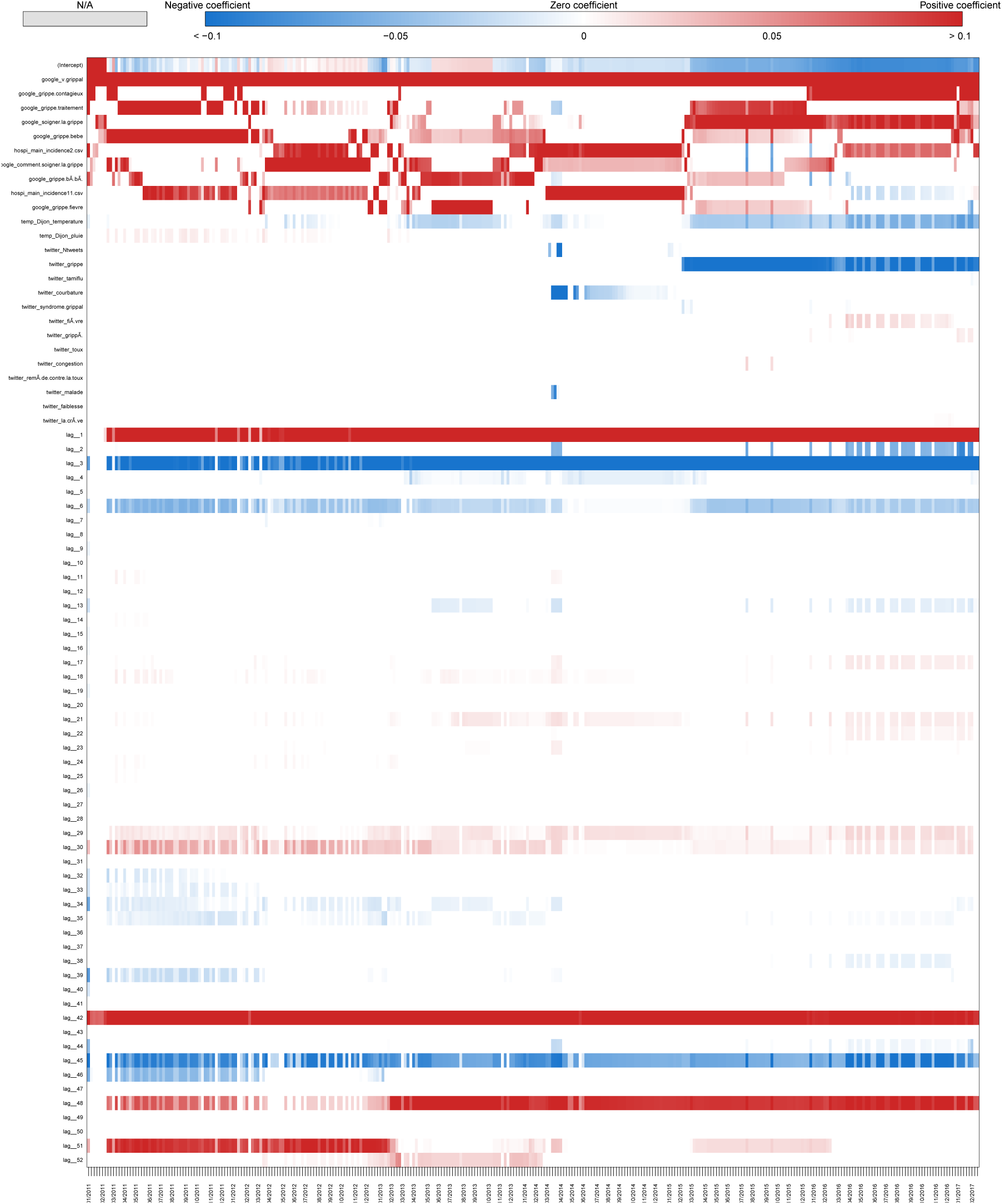
Coefficients Bourgogne Franche Comté One-week estimate.

**Fig S24.**
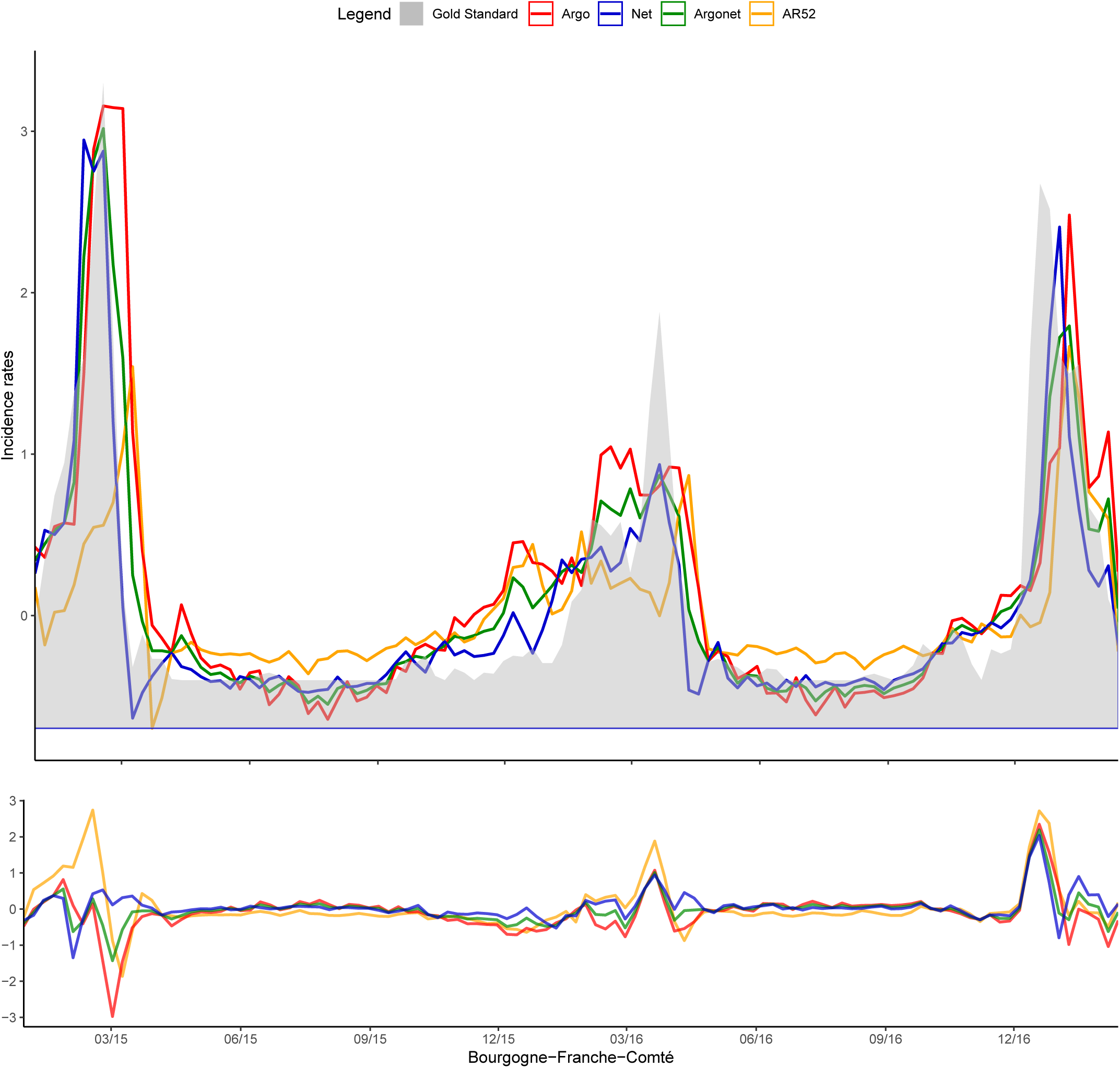
Bourgogne Two-week estimate.

**Fig S25.**
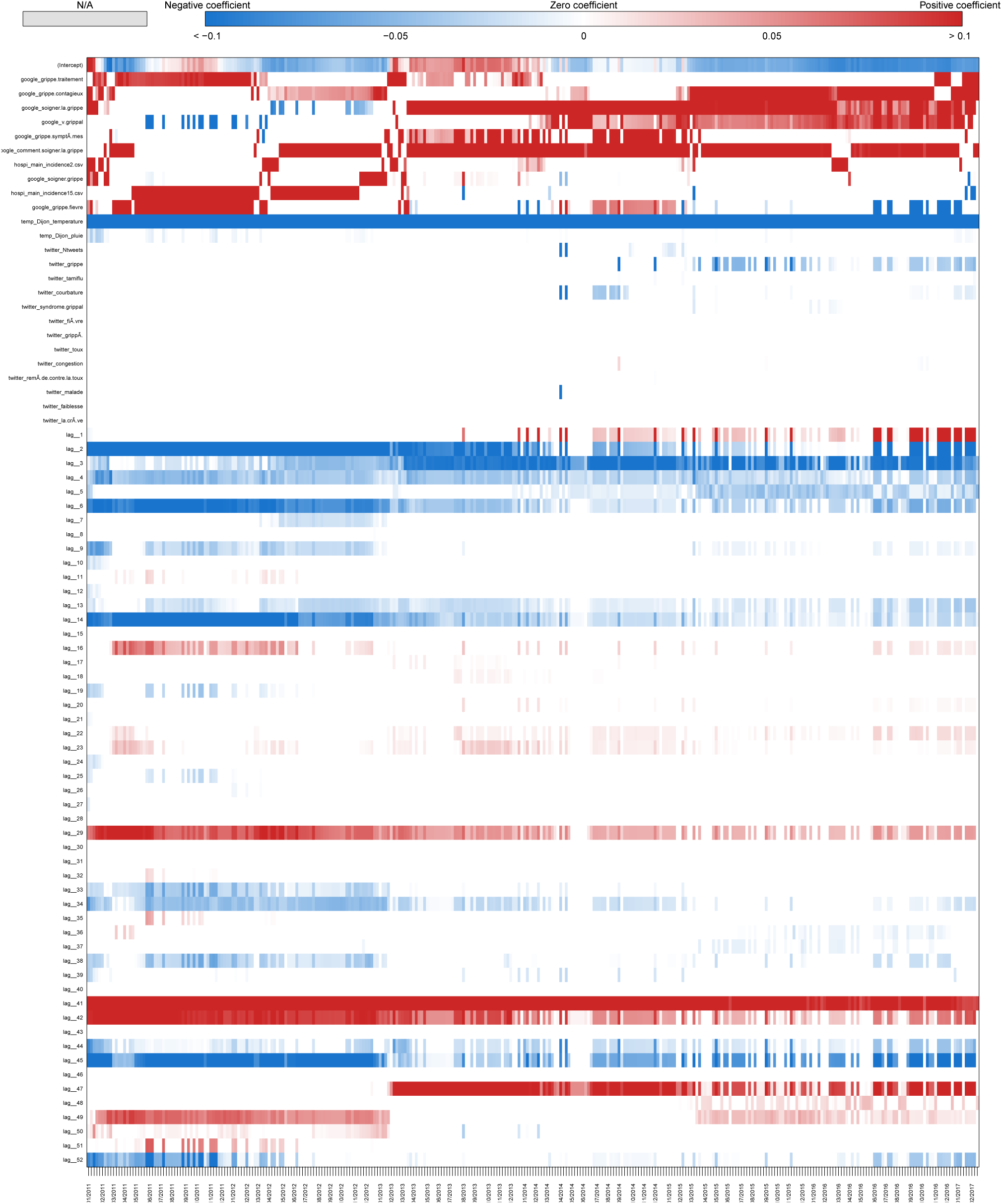
Coefficients Bourgogne Franche Comté Two-week estimate.

**Fig S26.**
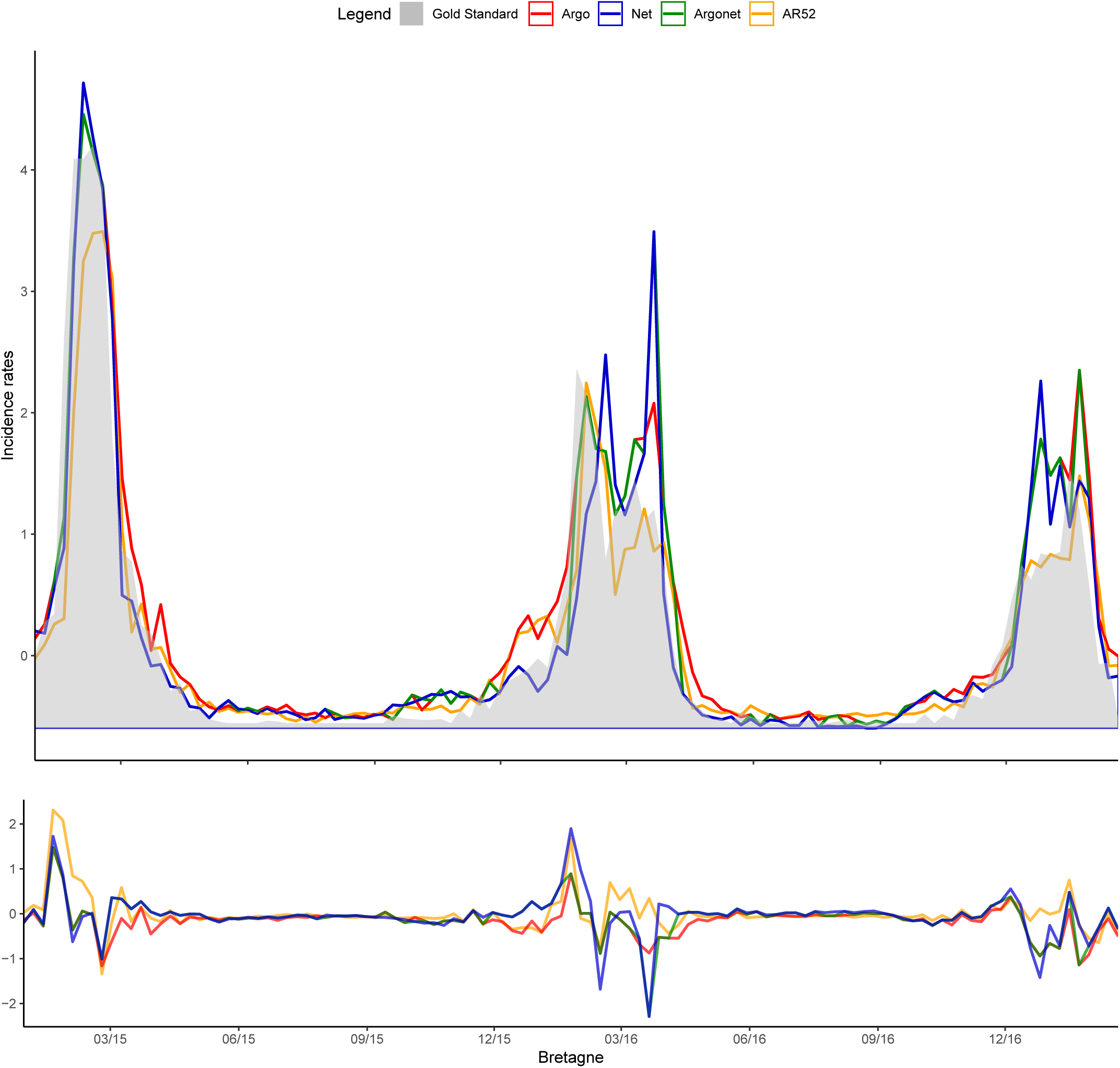
Bretagne Real-time estimate.

**Fig S27.**
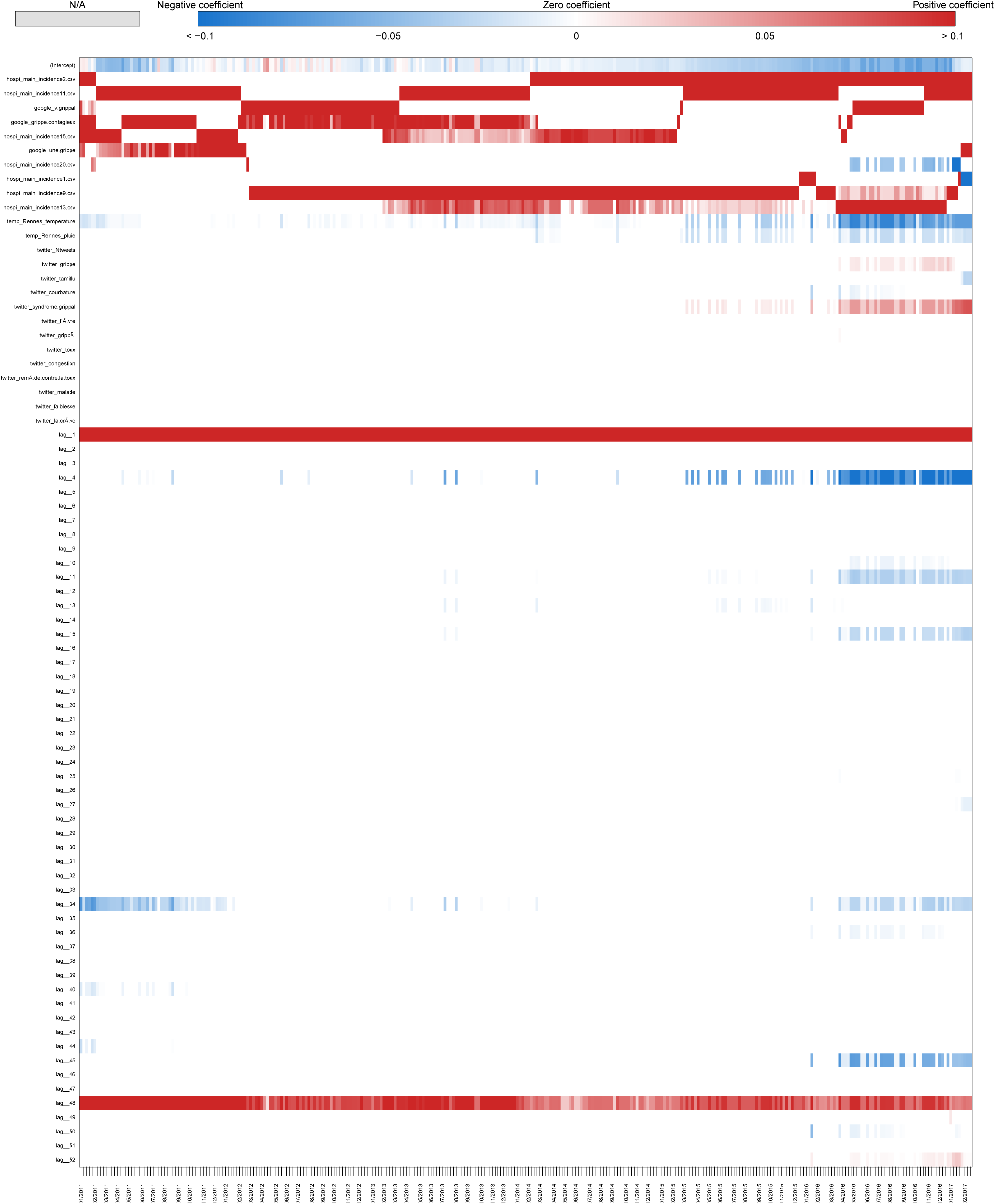
Coefficients Bretagne Real-time estimate.

**Fig S28.**
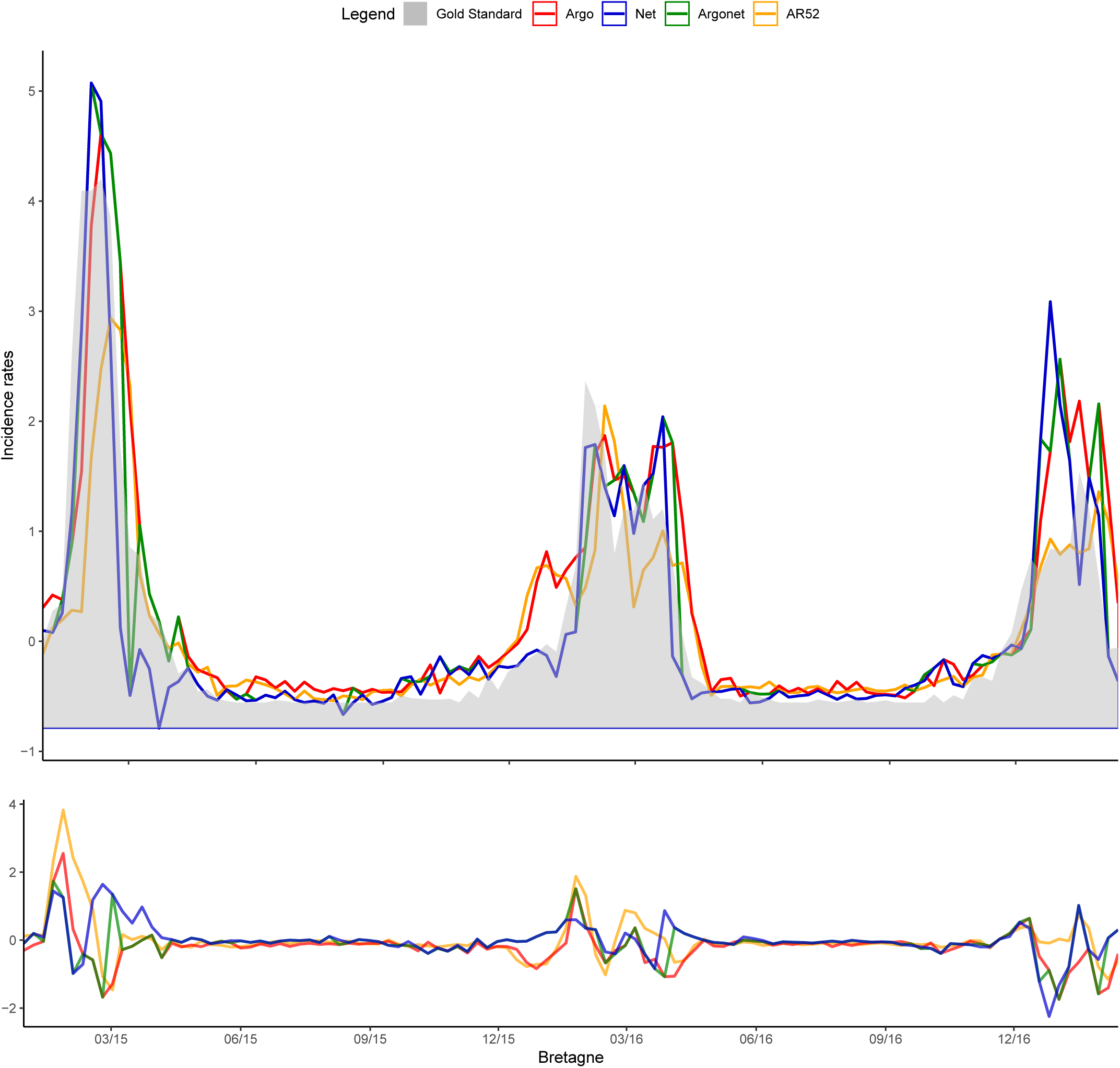
Bretagne One-week estimate.

**Fig S29.**
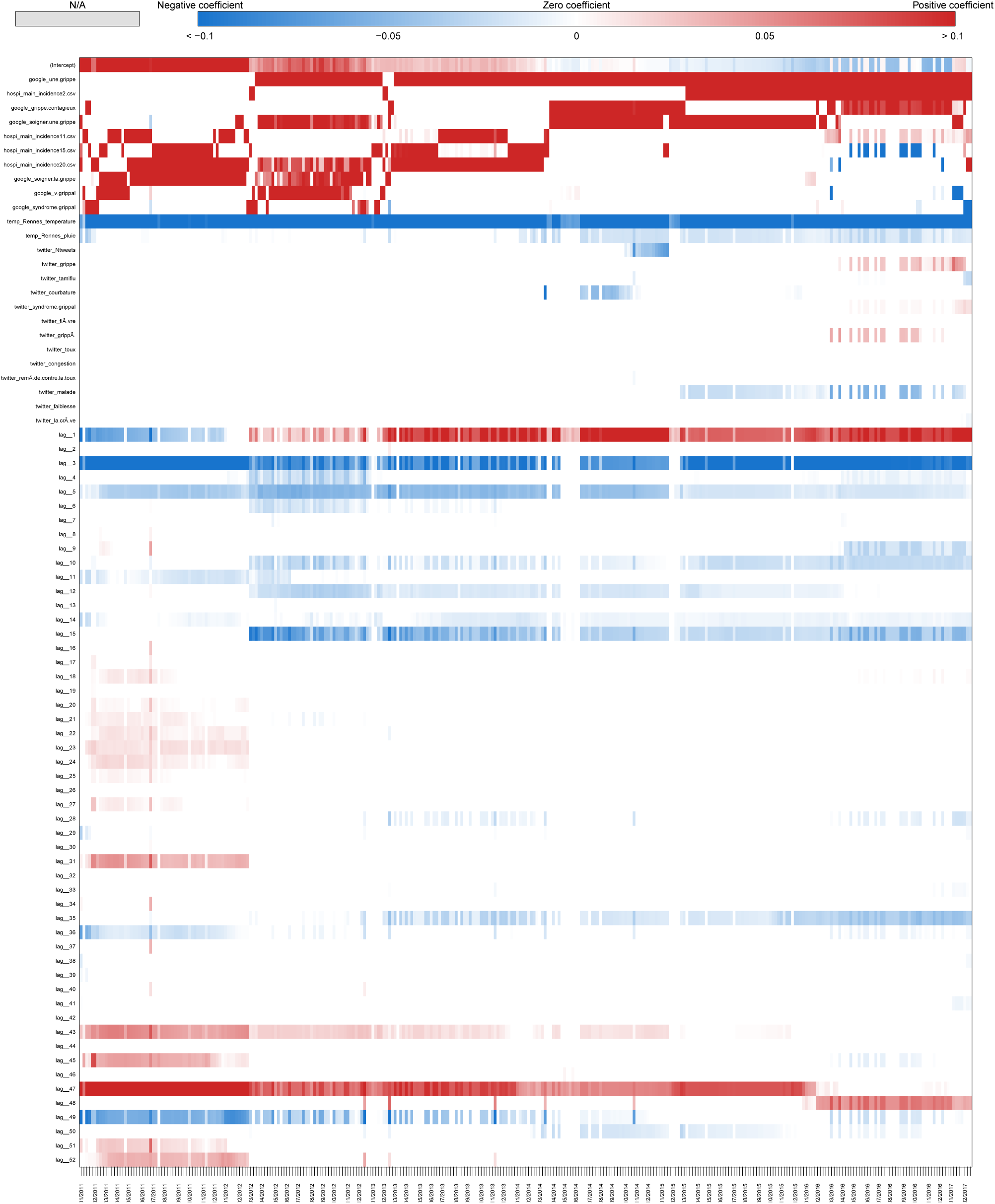
Coefficients Bretagne One-week estimate.

**Fig S30.**
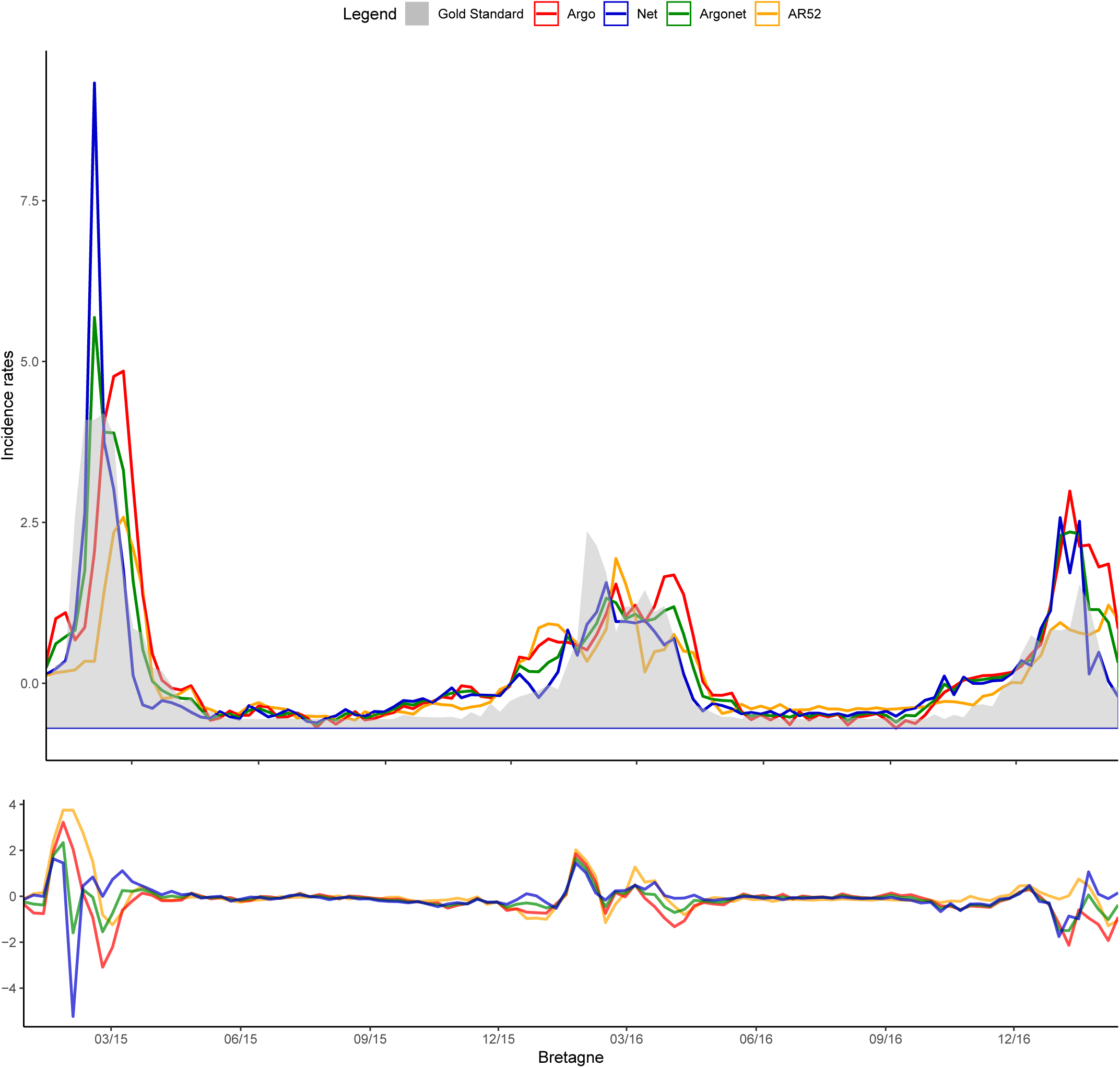
Bretagne Two-week estimate.

**Fig S31.**
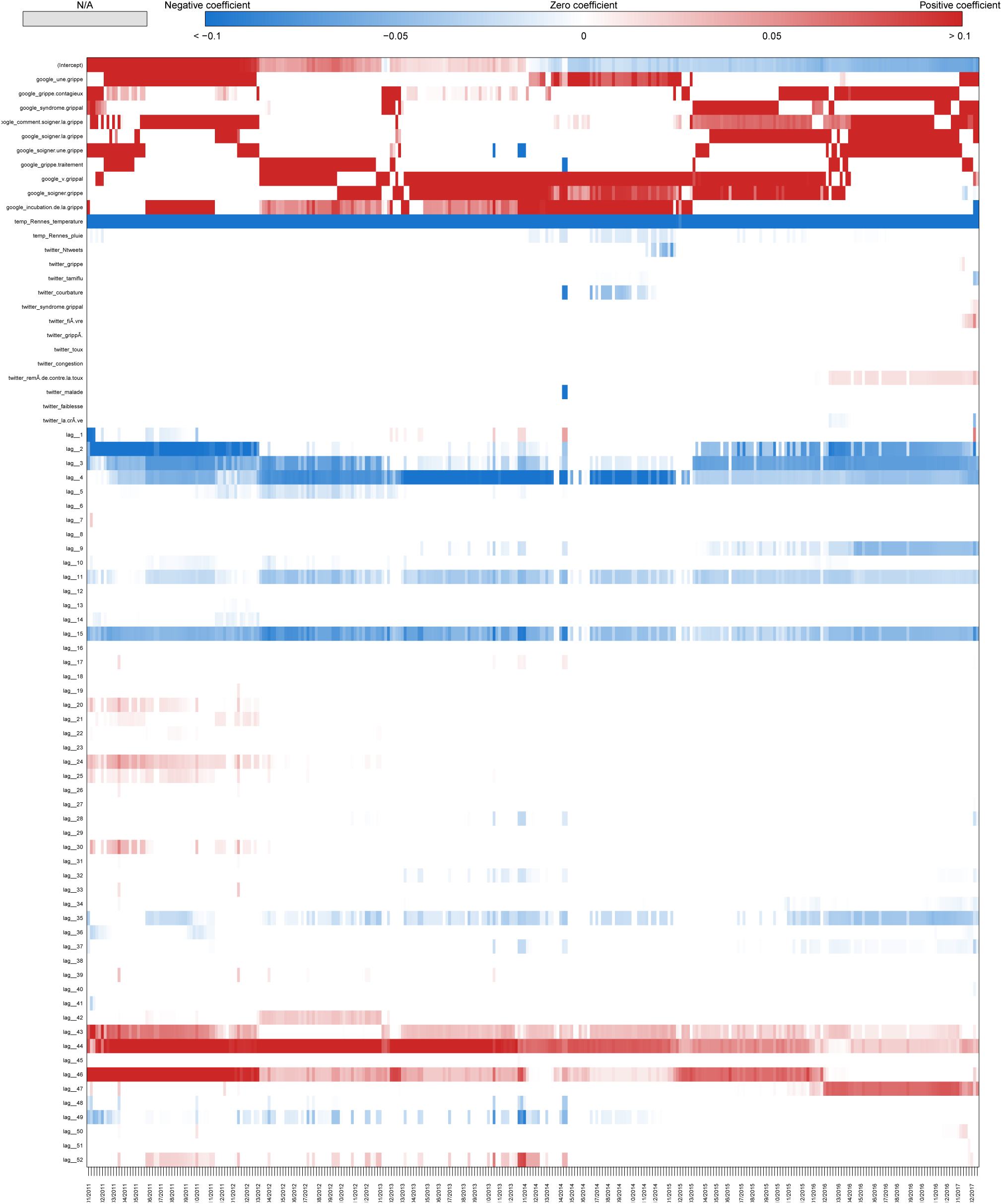
Coefficients Bretagne Two-week estimate.

**Fig S32.**
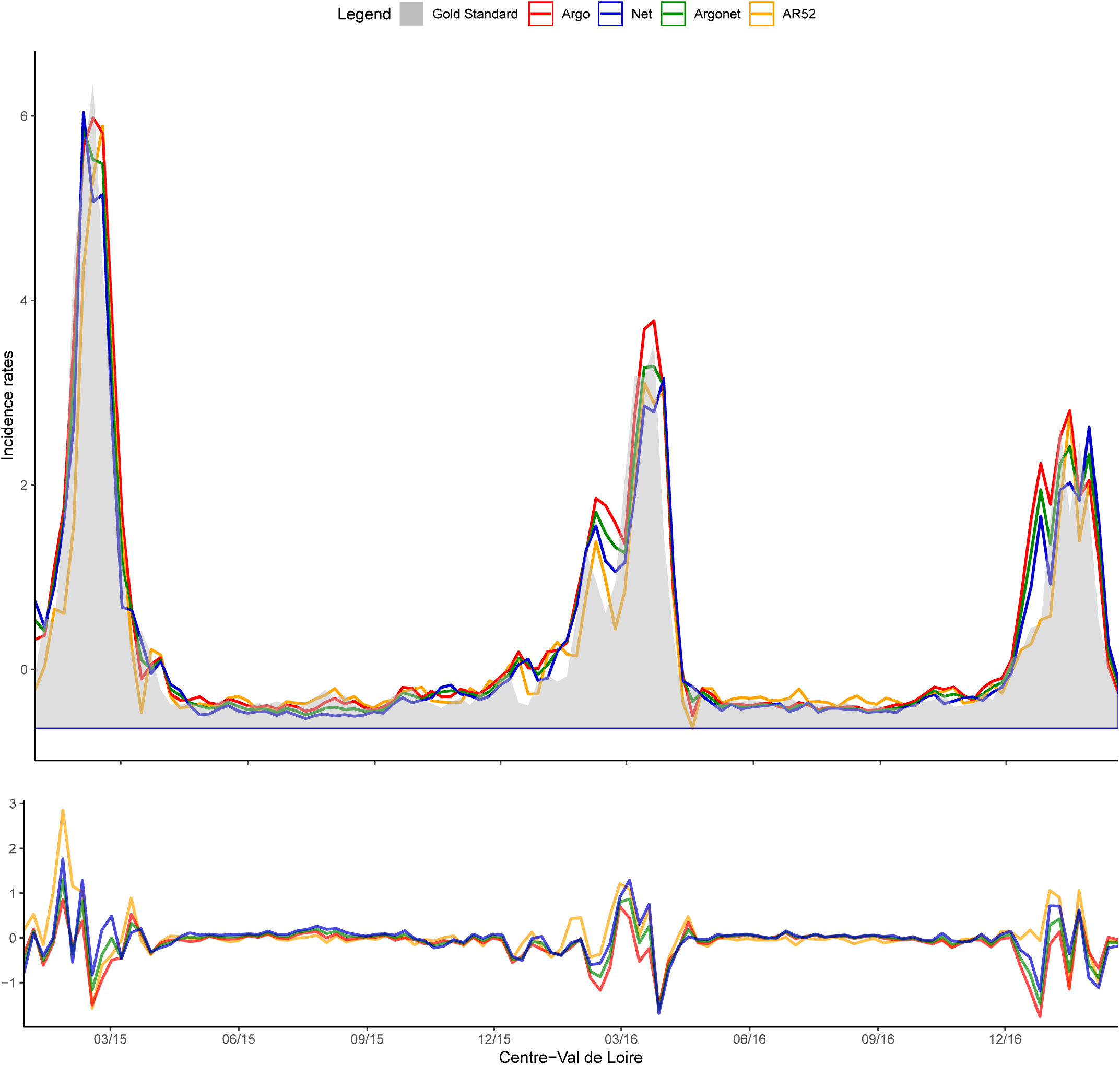
Centre Val-de-Loire Real-time estimate.

**Fig S33.**
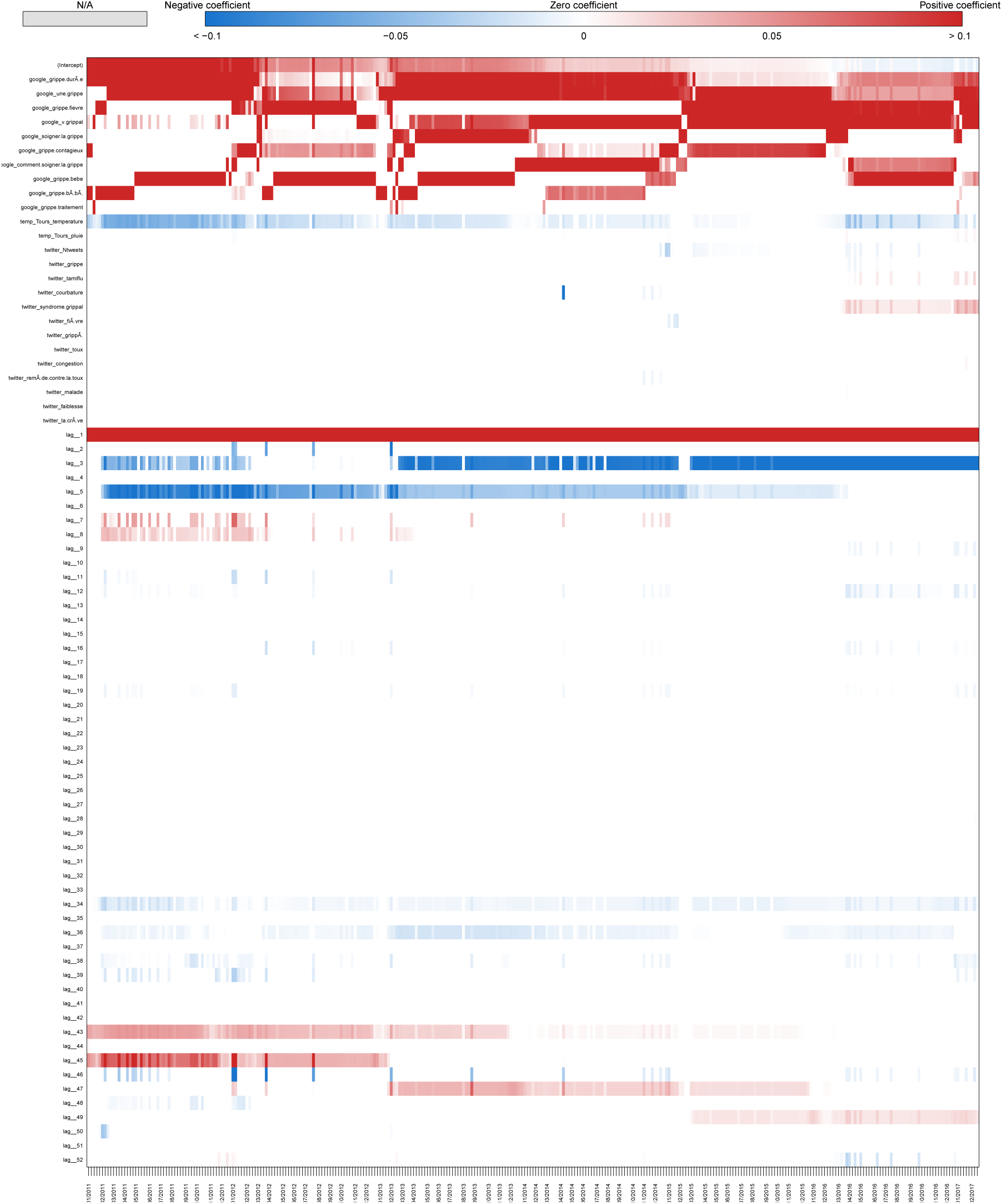
Coefficients Centre Val-de-Loire Real-time estimate.

**Fig S34.**
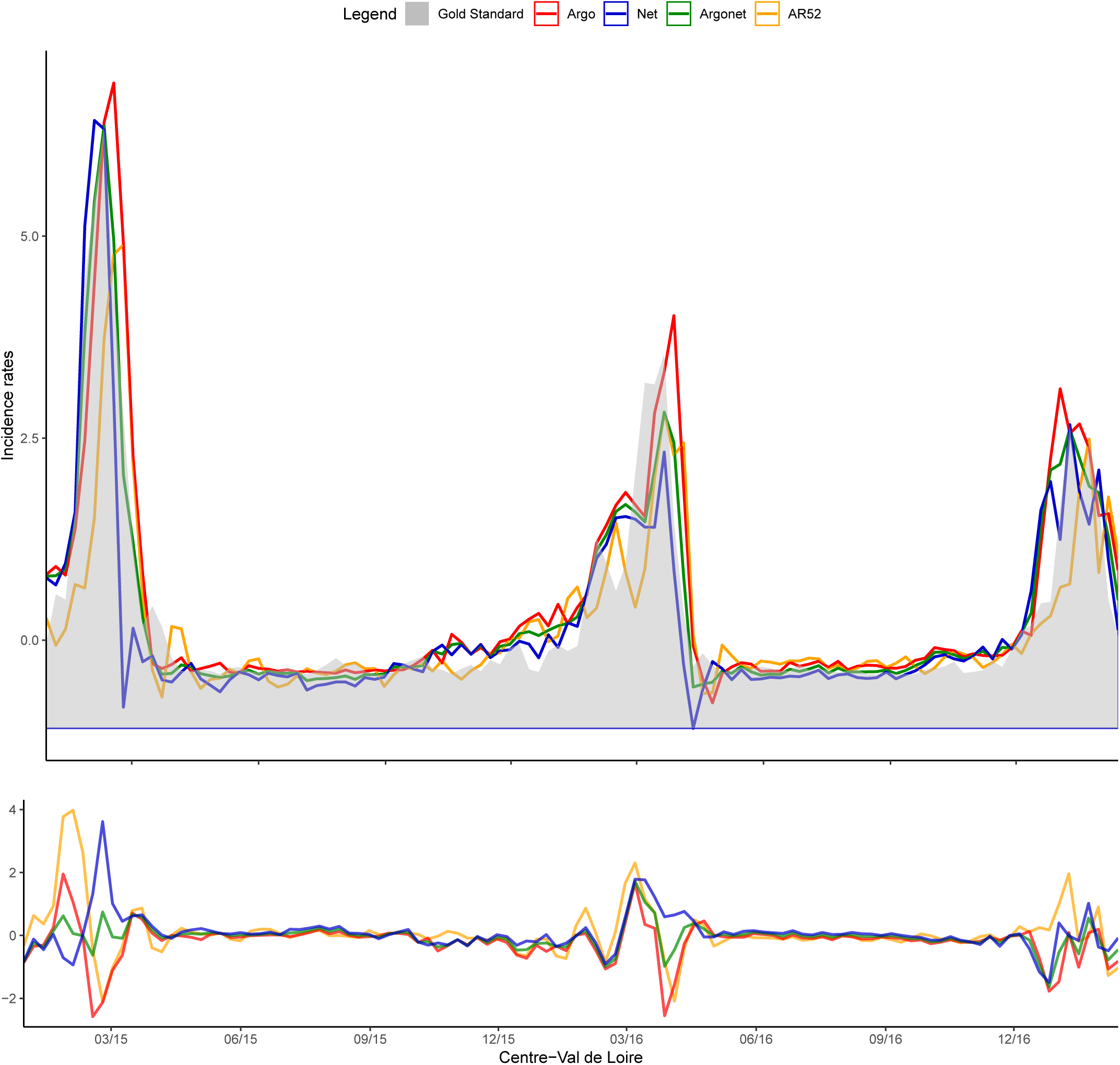
Centre Val-de-Loire One-week estimate.

**Fig S35.**
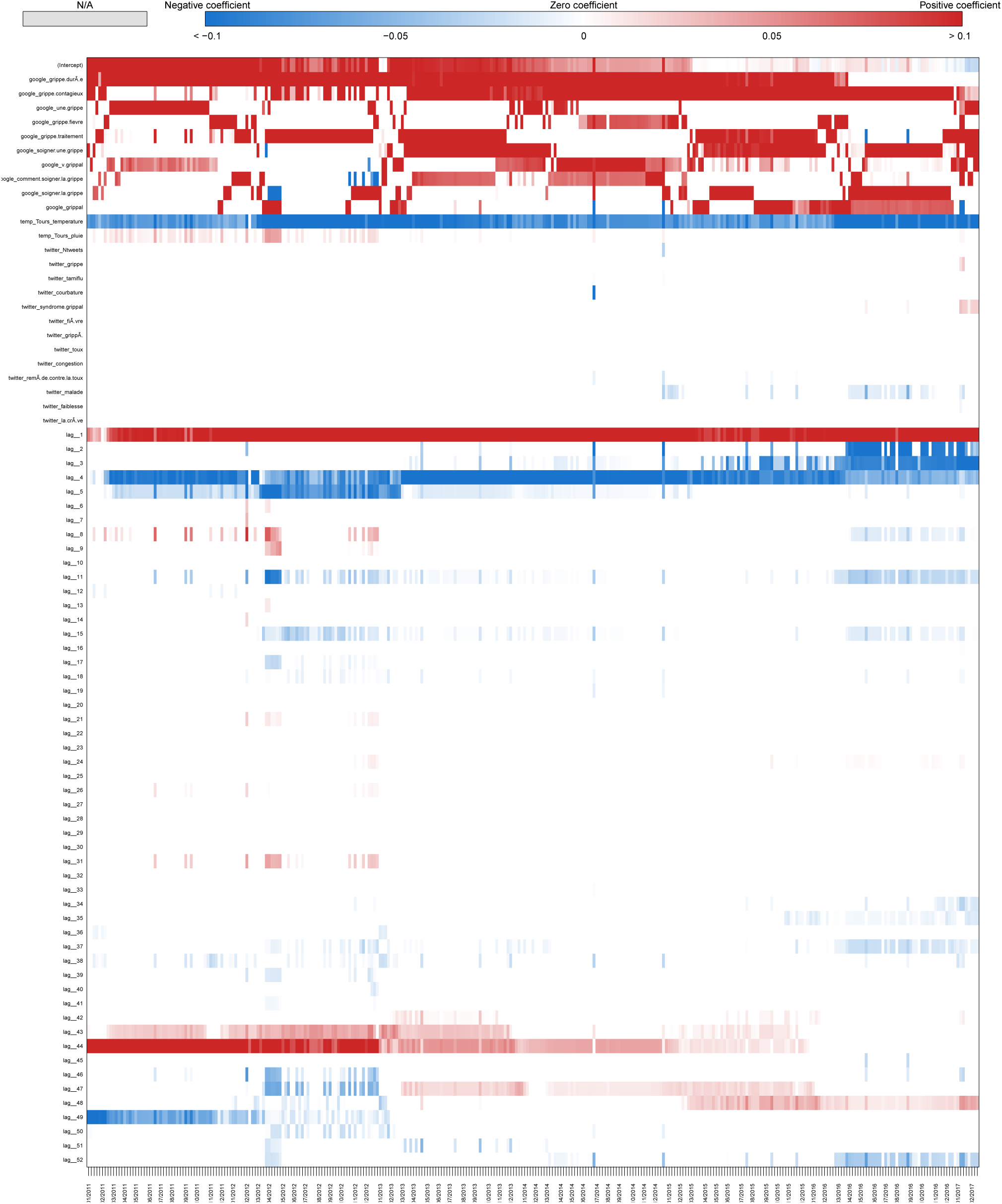
Coefficients Centre Val-de-Loire One-week estimate.

**Fig S36.**
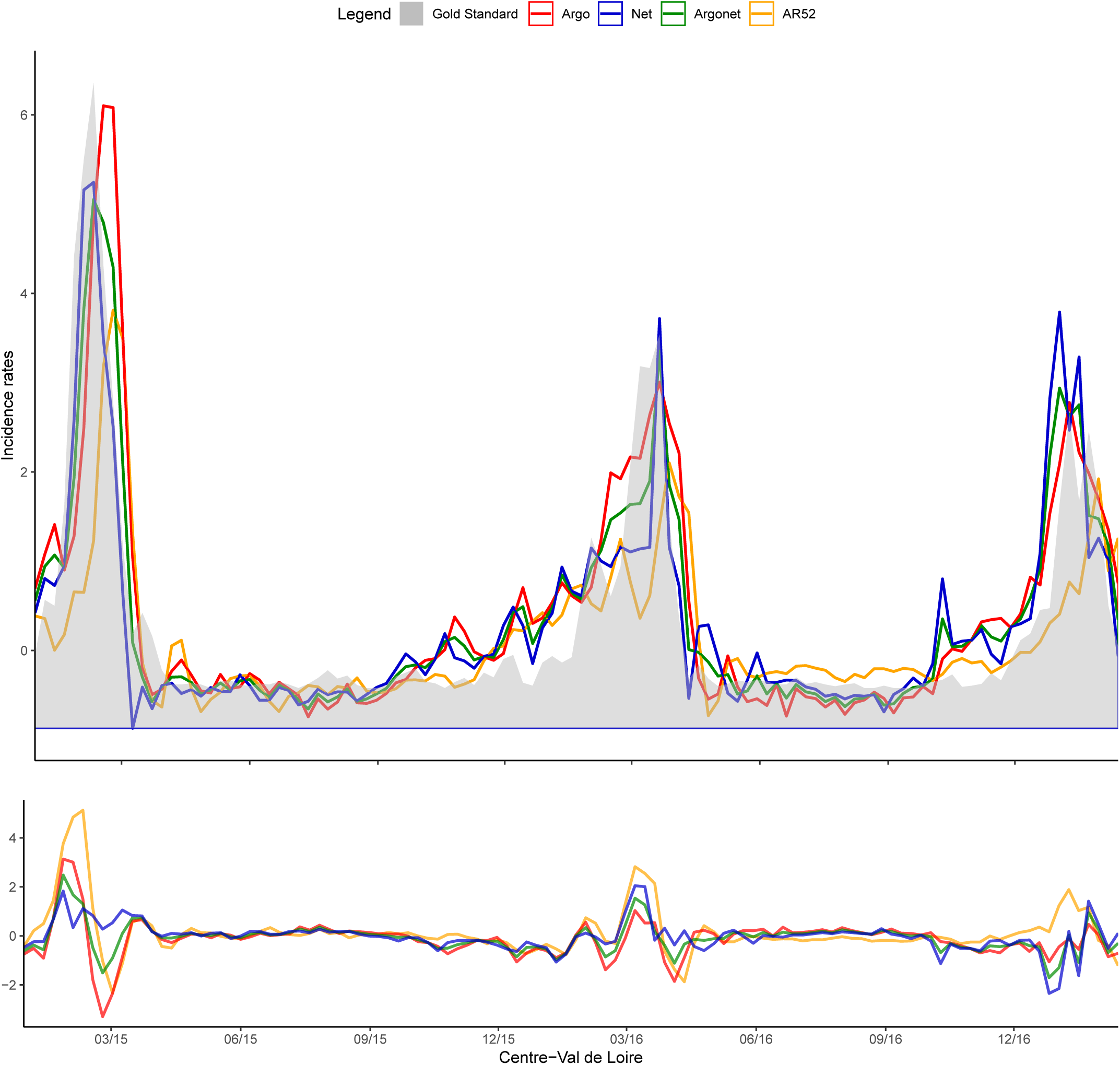
Centre Val-de-Loire Two-week estimate.

**Fig S37.**
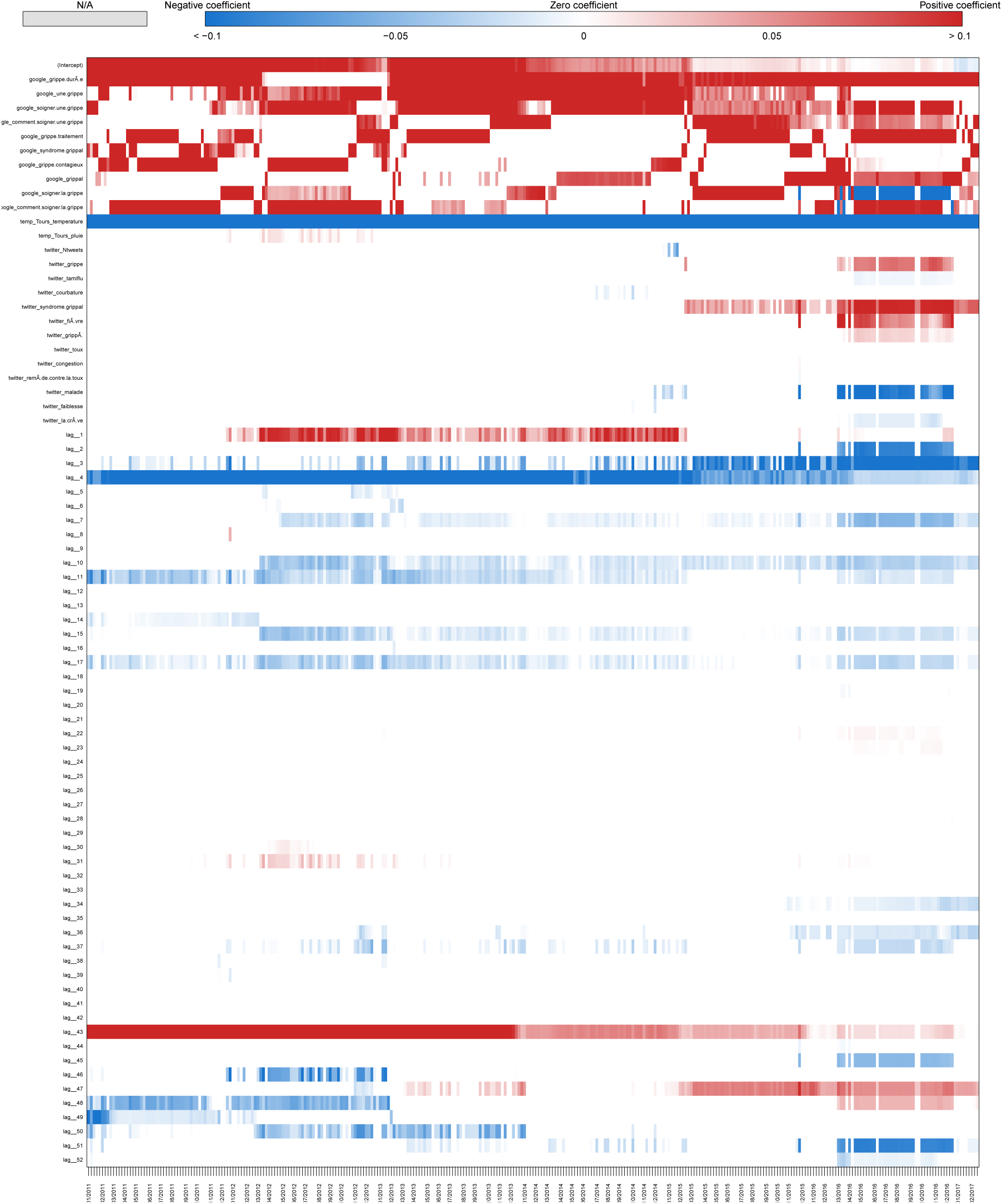
Coefficients Centre Val-de-Loire Two-week estimate.

**Fig S38.**
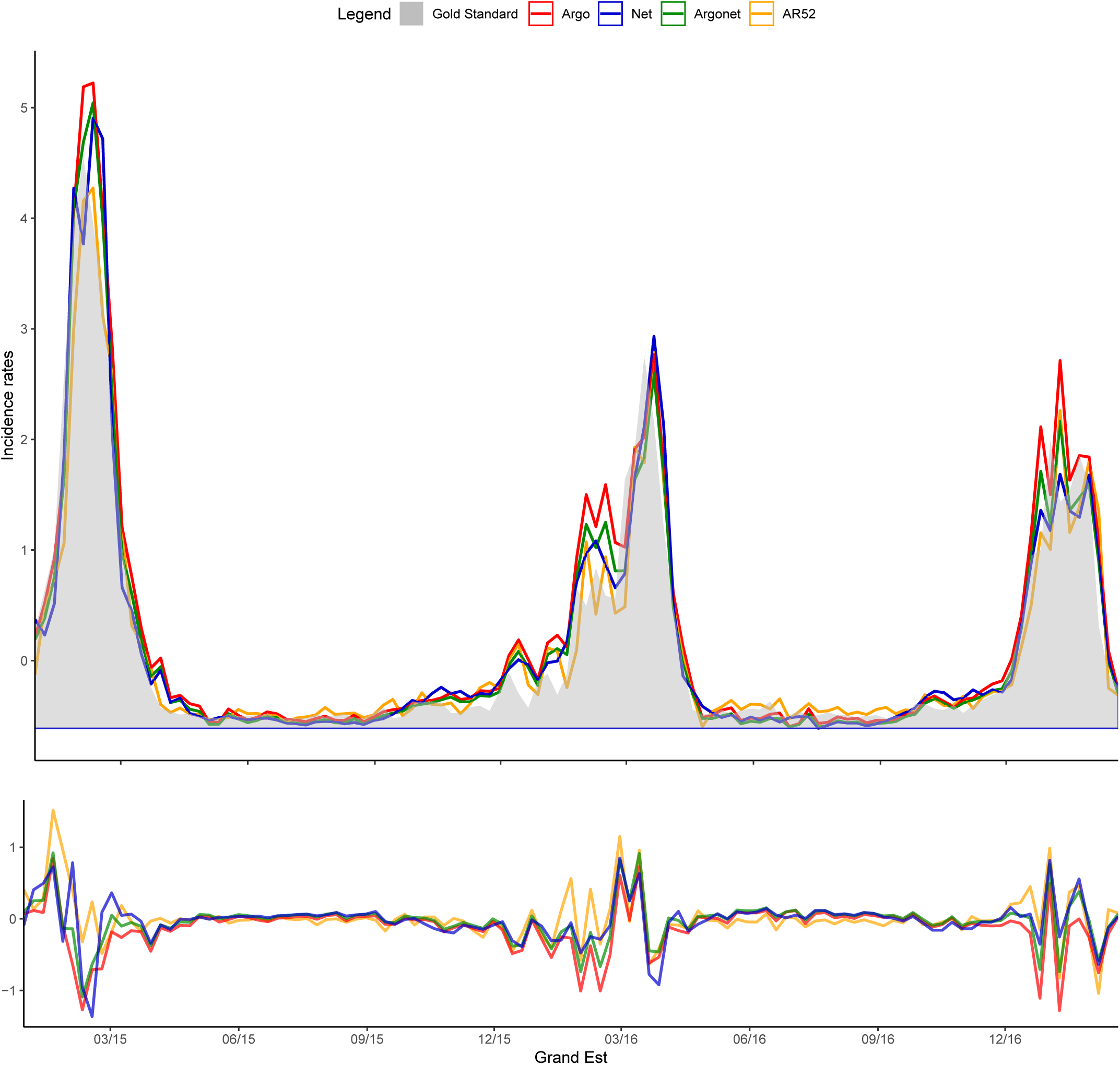
Grand Est Real-time estimate.

**Fig S39.**
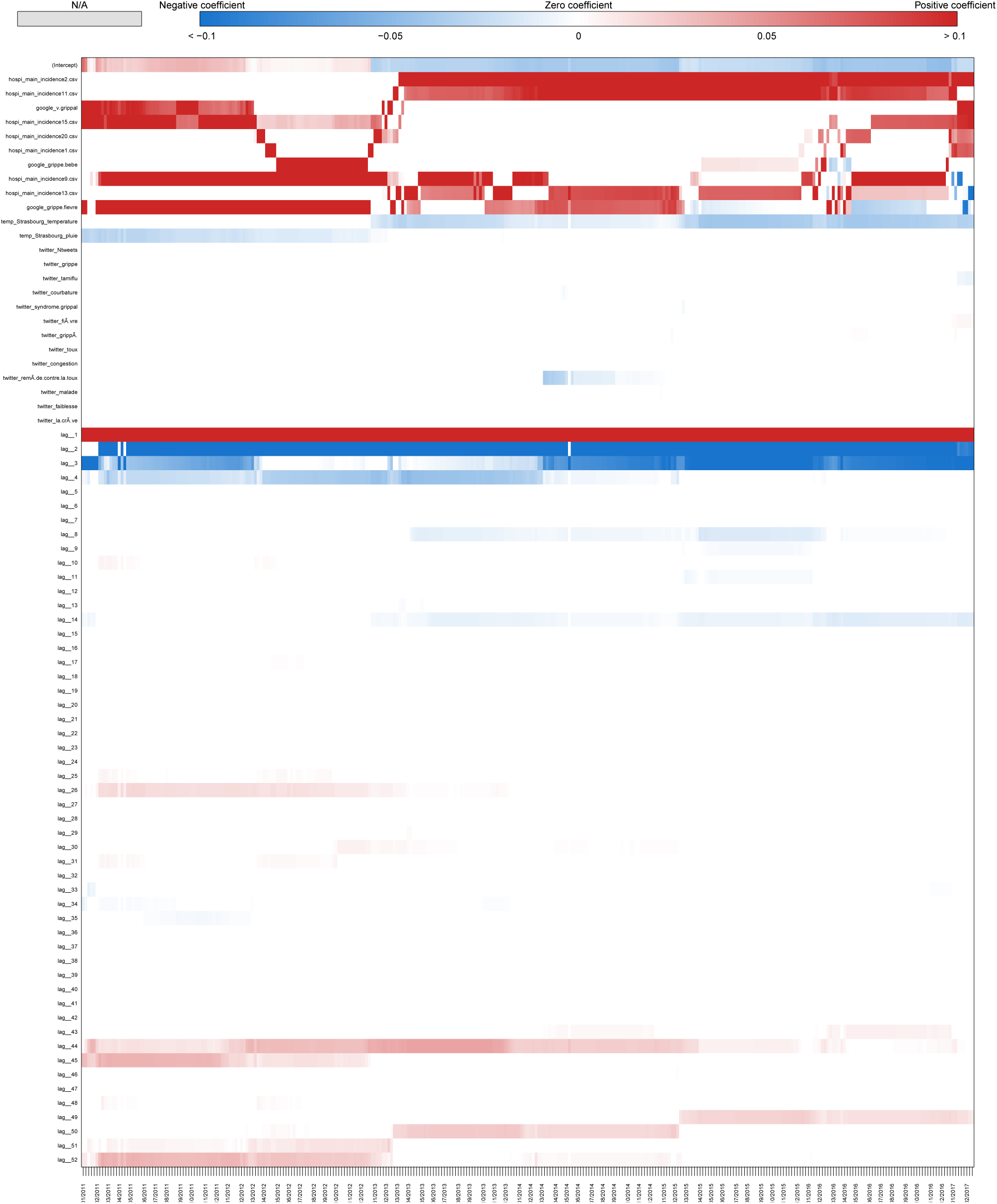
Coefficients Grand Est Real-time estimate.

**Fig S40.**
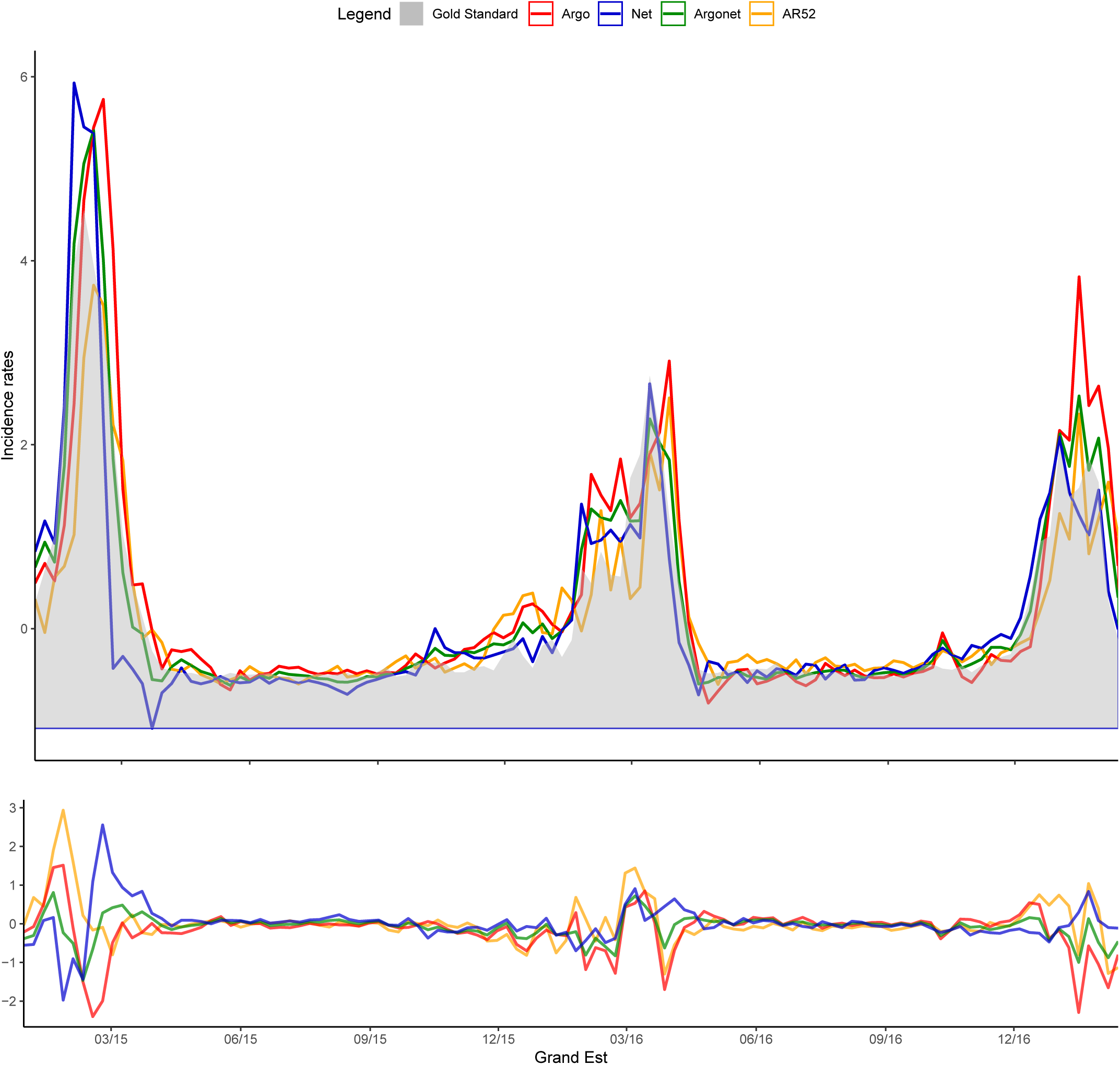
Grand Est One-week estimate.

**Fig S41.**
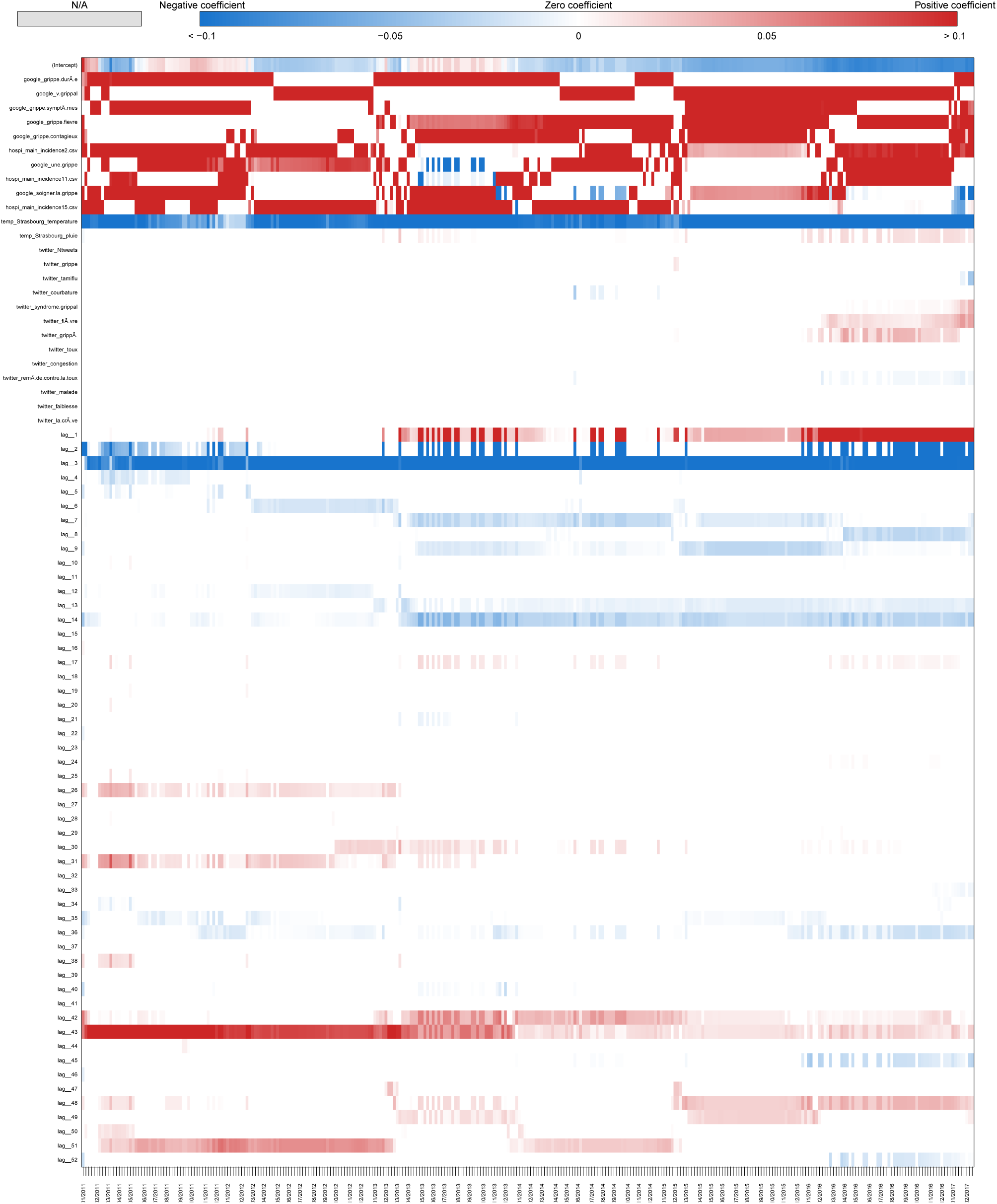
Coefficients Grand Est One-week estimate.

**Fig S42.**
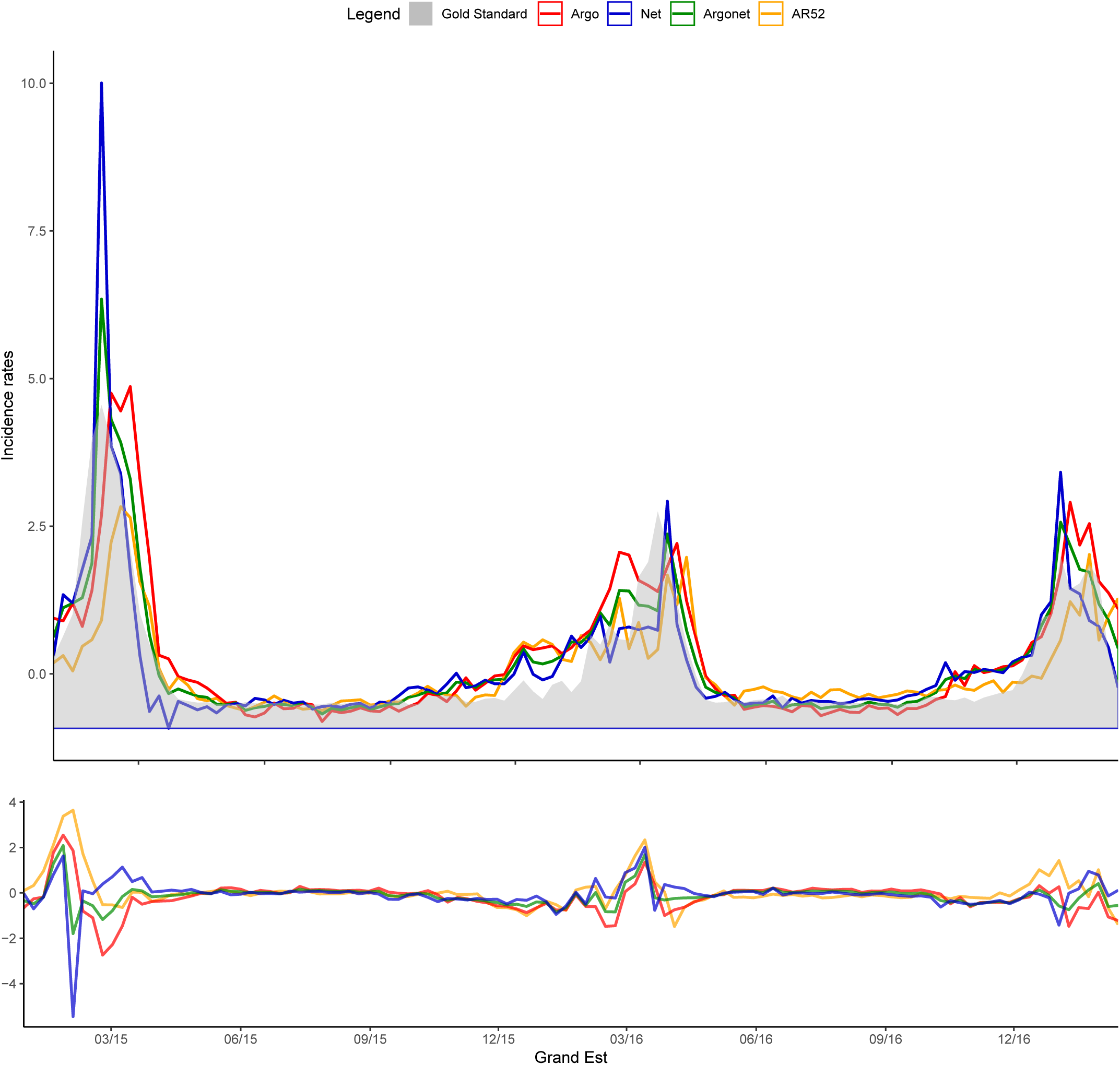
Grand Est Two-week estimate.

**Fig S43.**
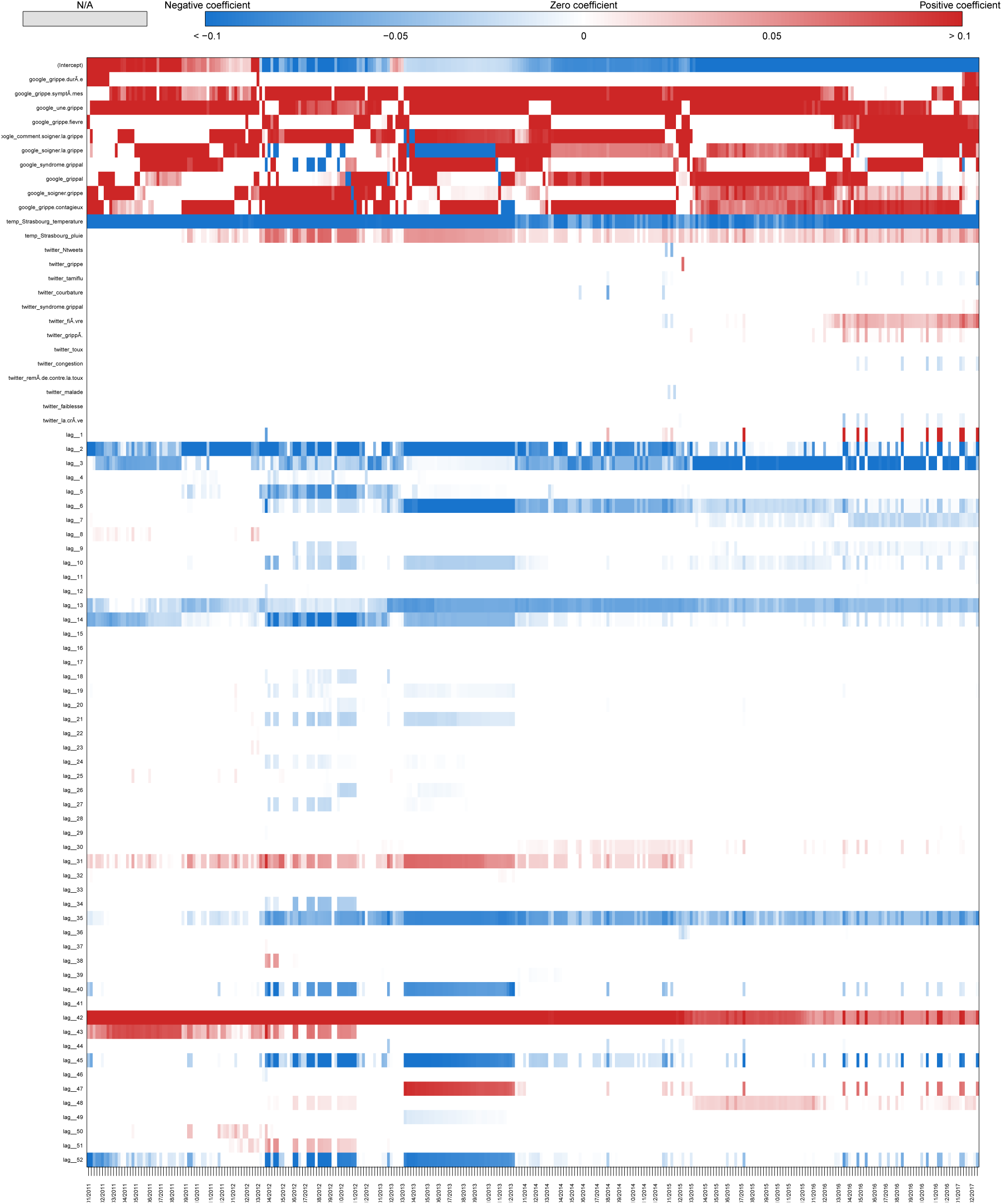
Coefficients Grand Est Two-week estimate.

**Fig S44.**
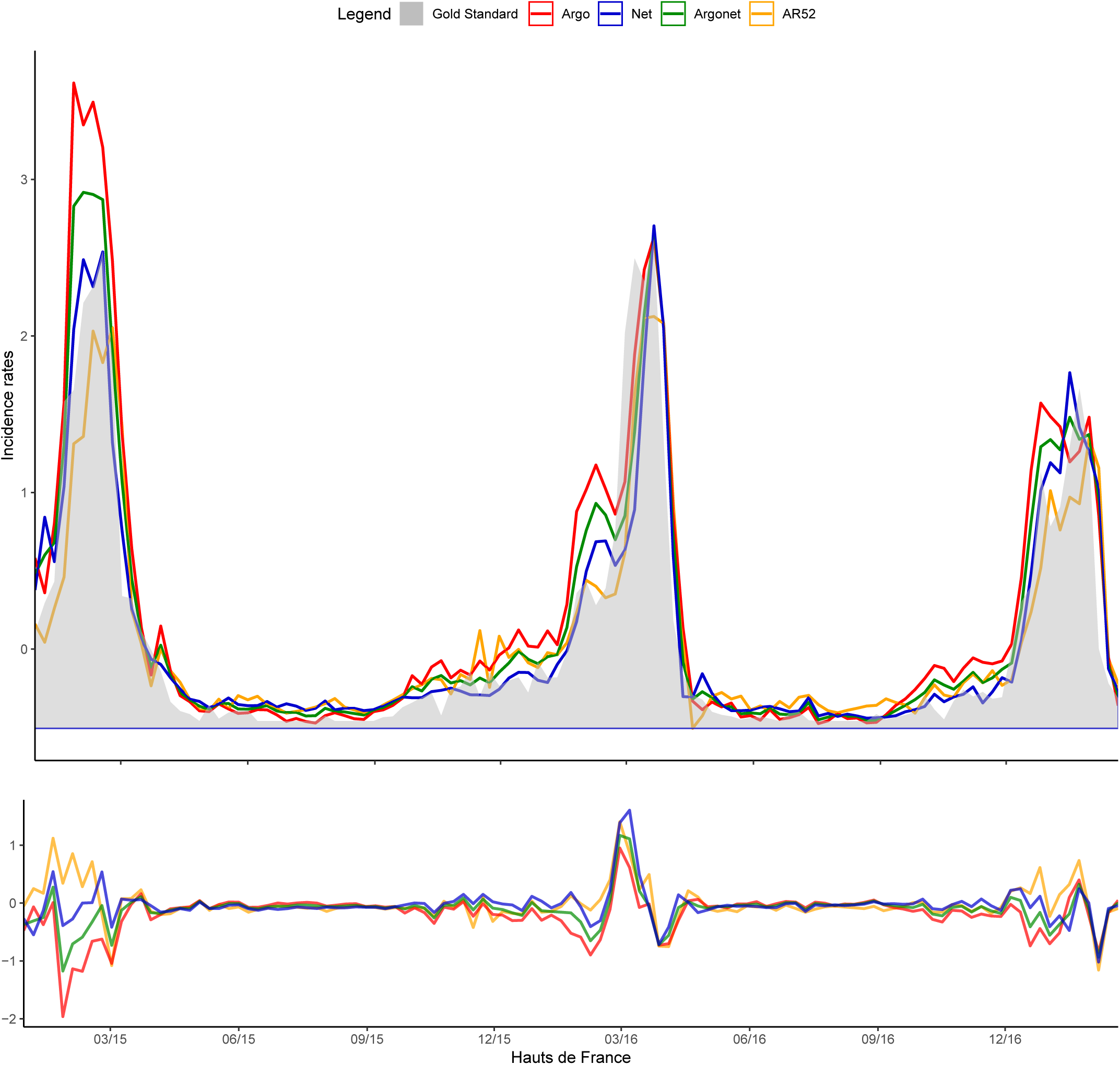
Hauts de France Real-time estimate.

**Fig S45.**
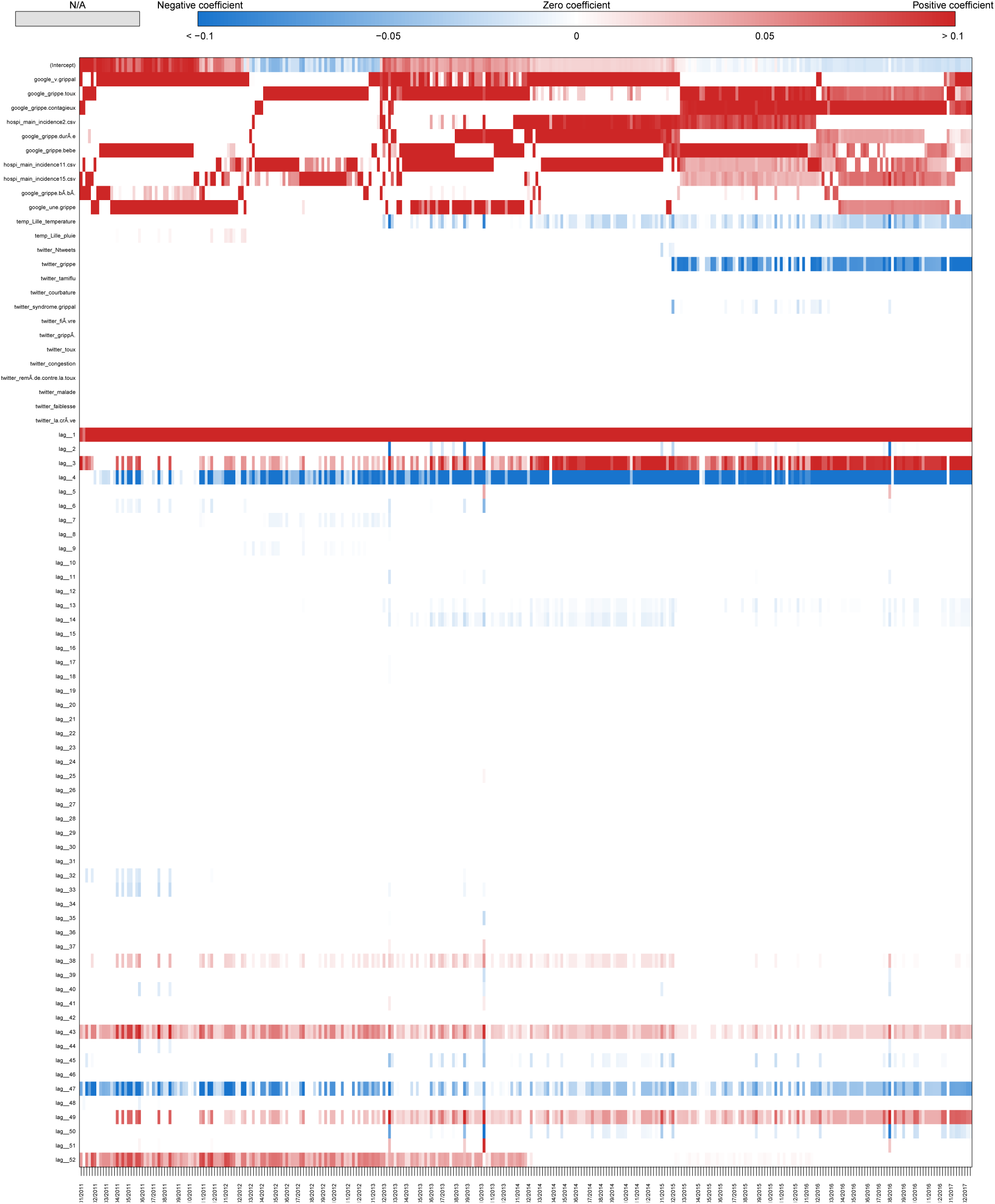
Coefficients Hauts de France Real-time estimate.

**Fig S46.**
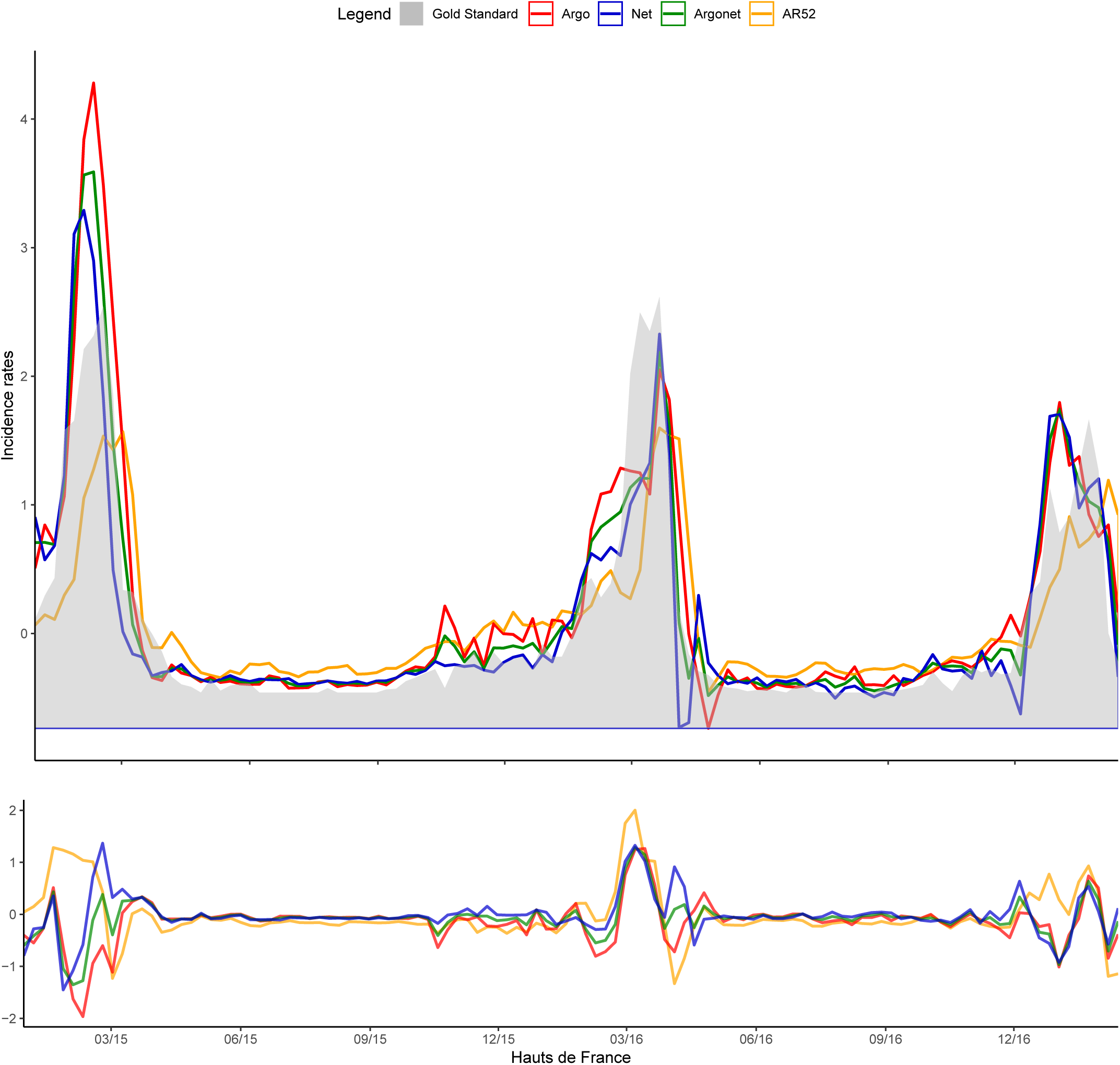
Hauts de France One-week estimate.

**Fig S47.**
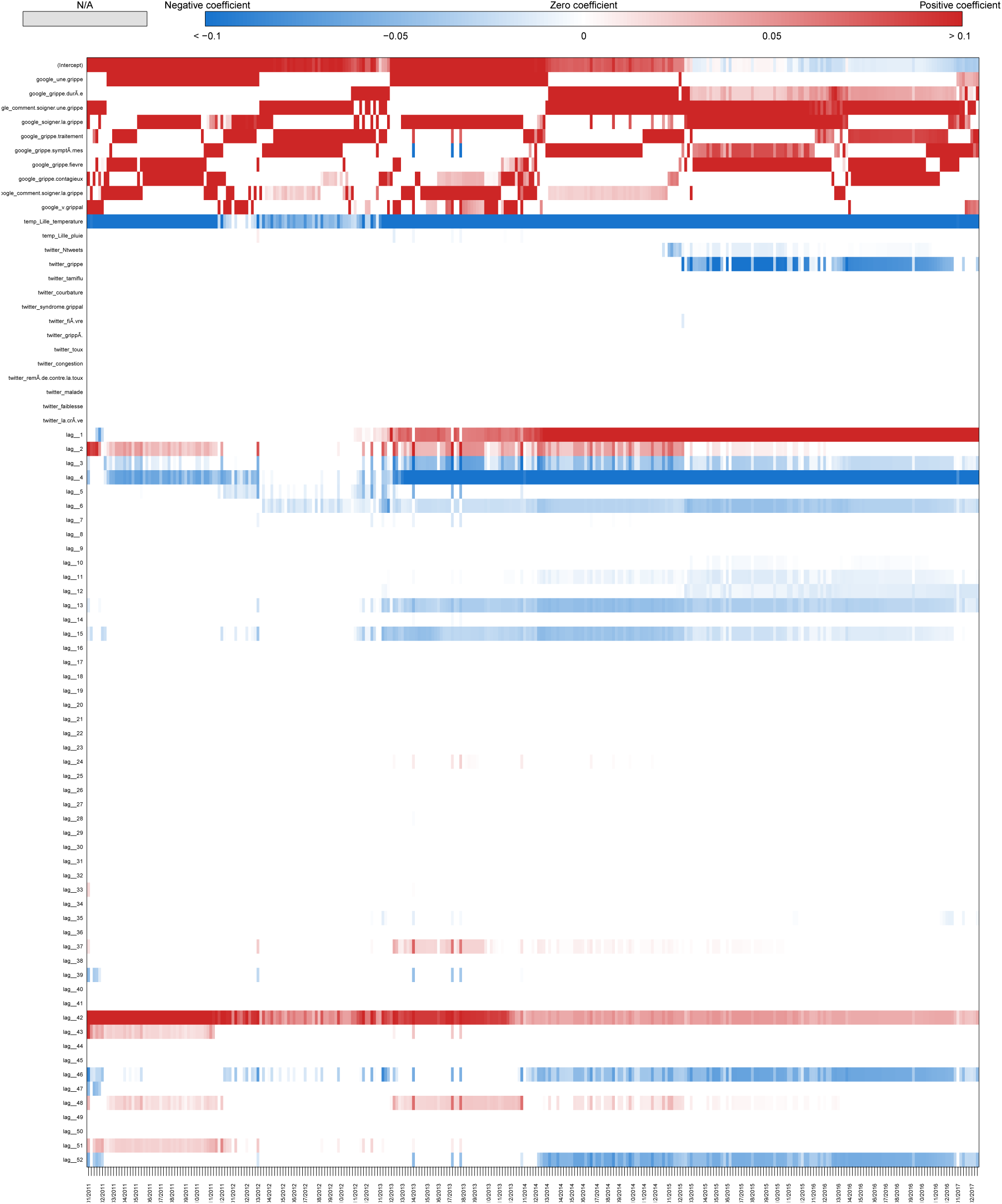
Coefficients Hauts de France One-week estimate.

**Fig S48.**
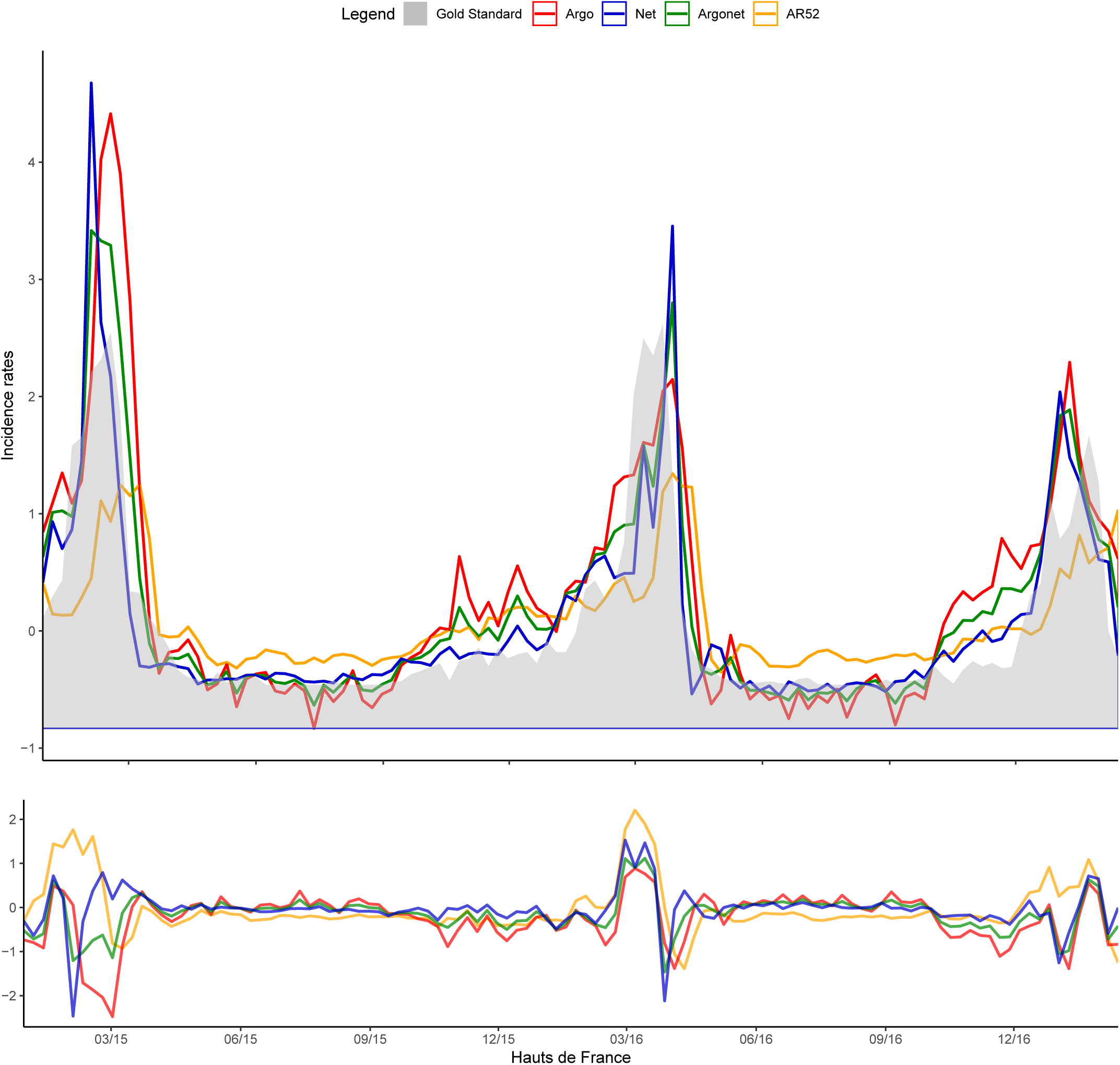
Hauts de France Two-week estimate.

**Fig S49.**
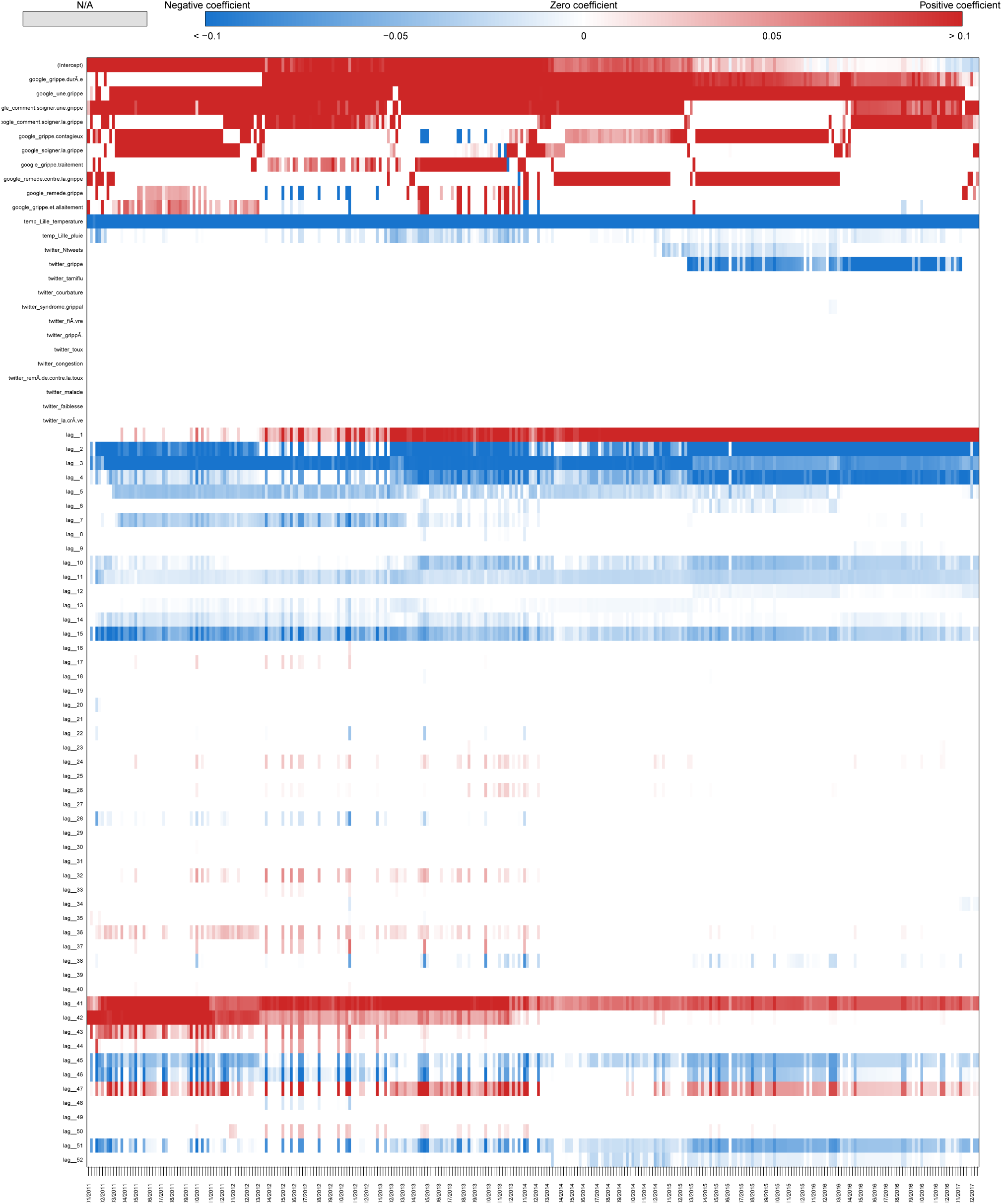
Coefficients Hauts de France Two-week estimate.

**Fig S50.**
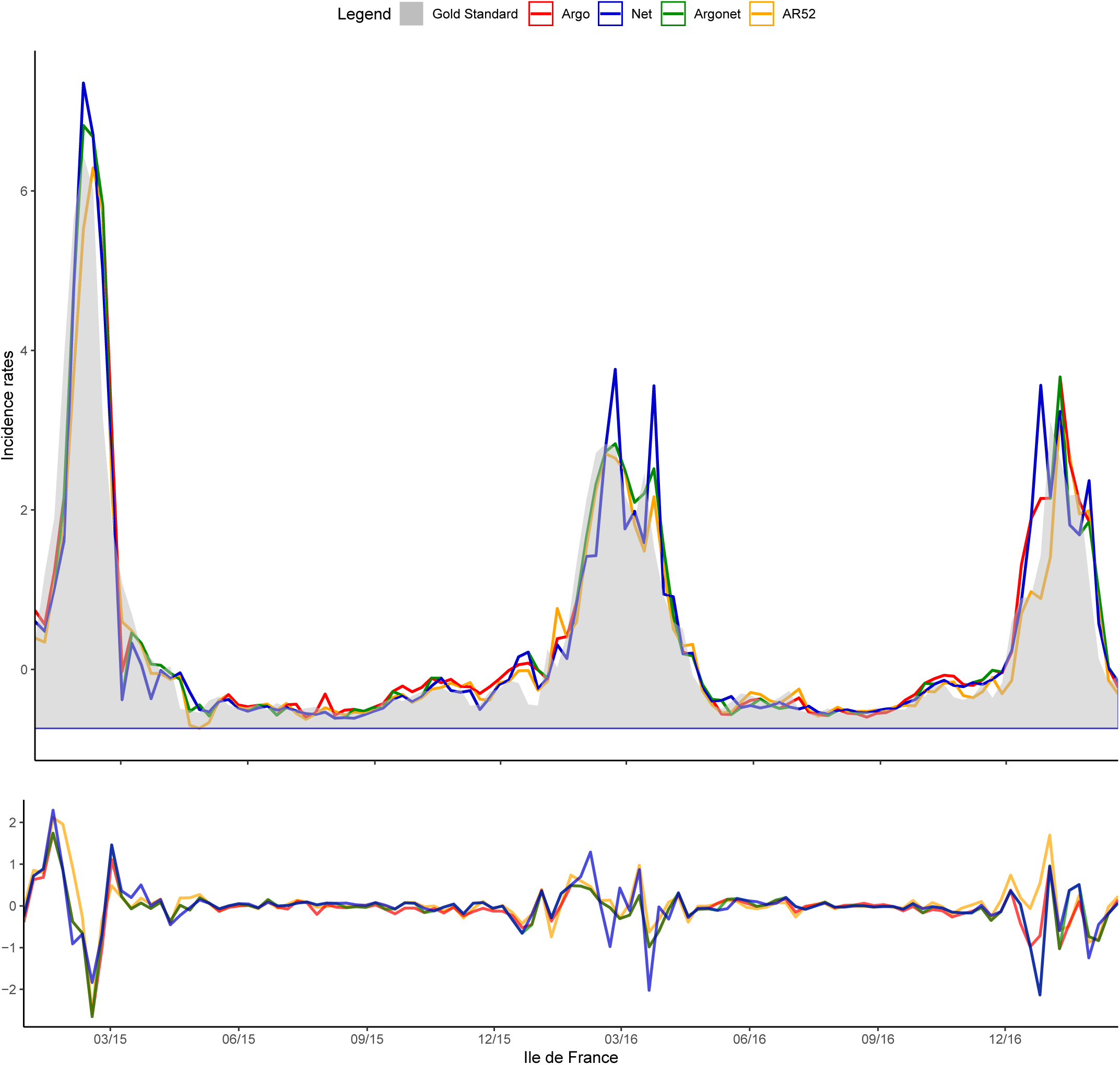
Ile de France Real-time estimate.

**Fig S51.**
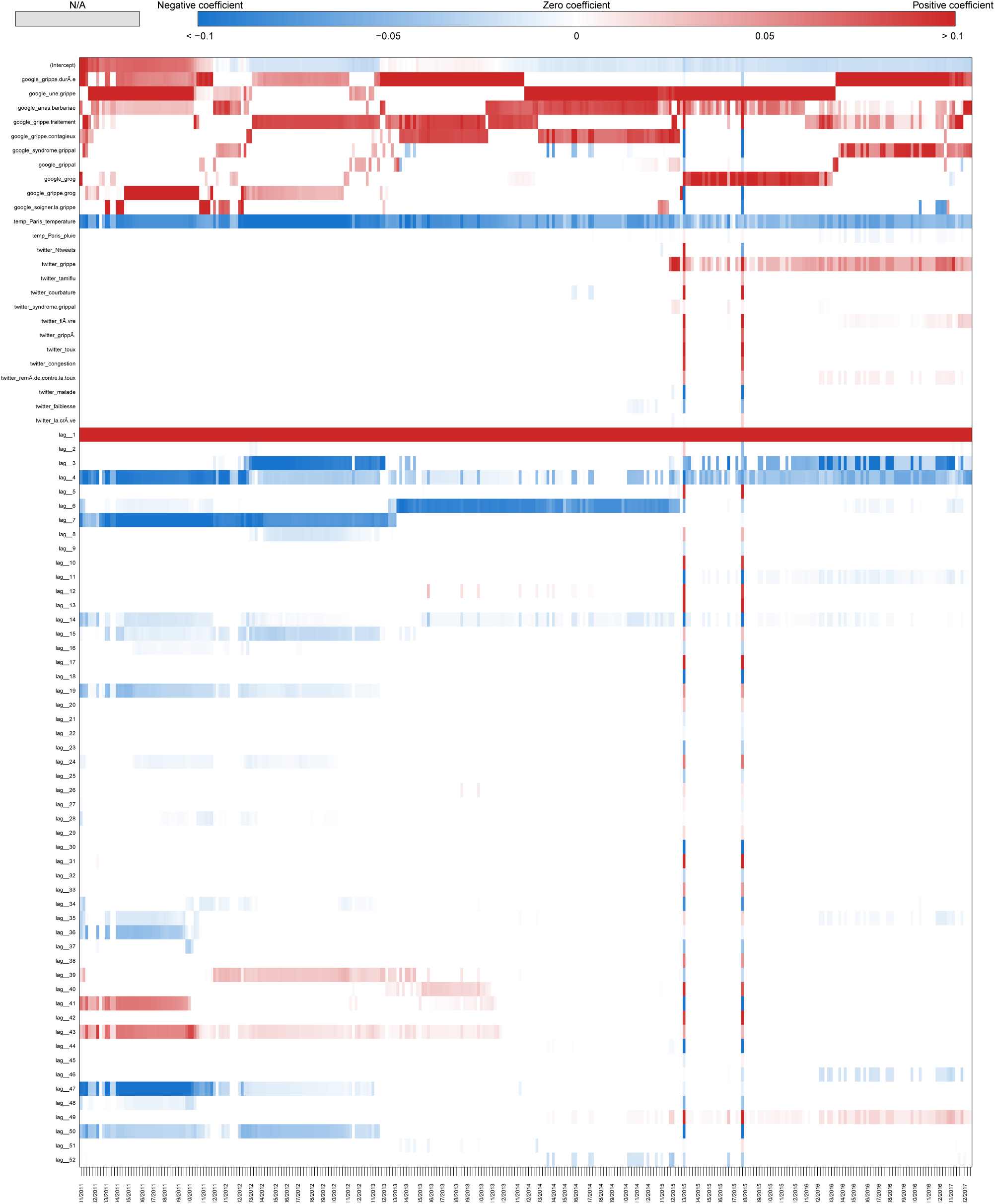
Coefficients Ile de France Real-time estimate.

**Fig S52.**
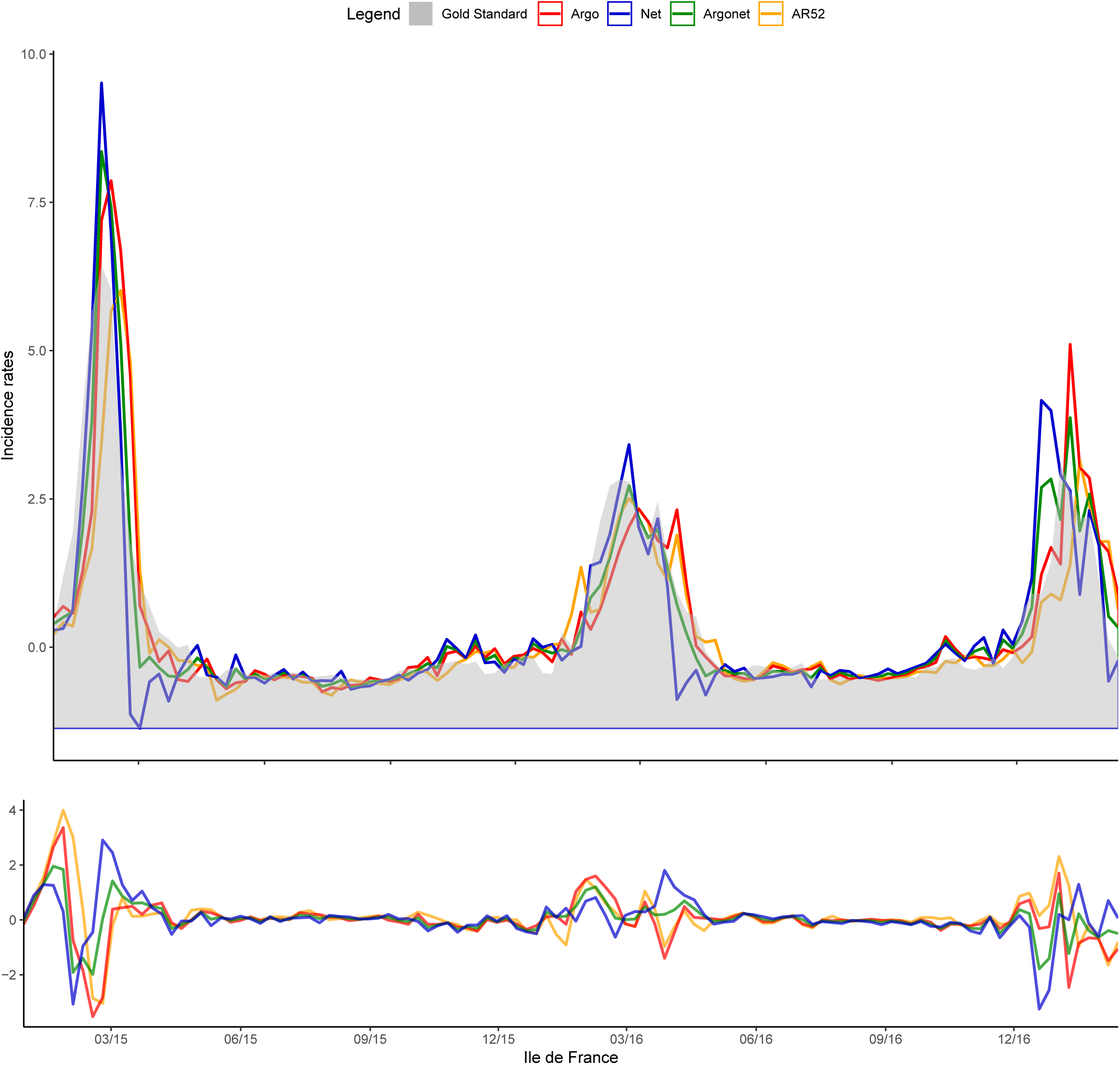
Ile de France One-week estimate.

**Fig S53.**
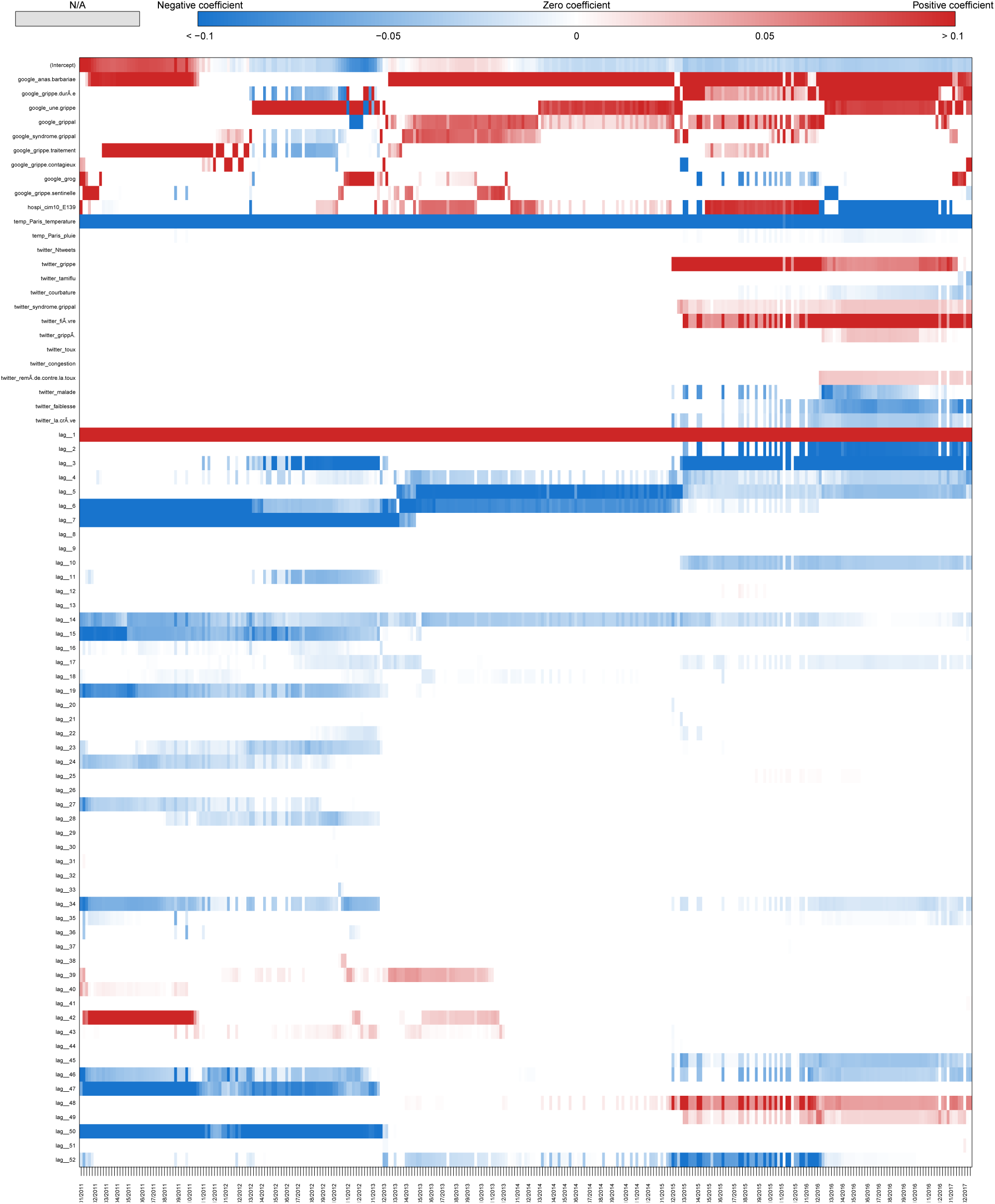
Coefficients Ile de France One-week estimate.

**Fig S54.**
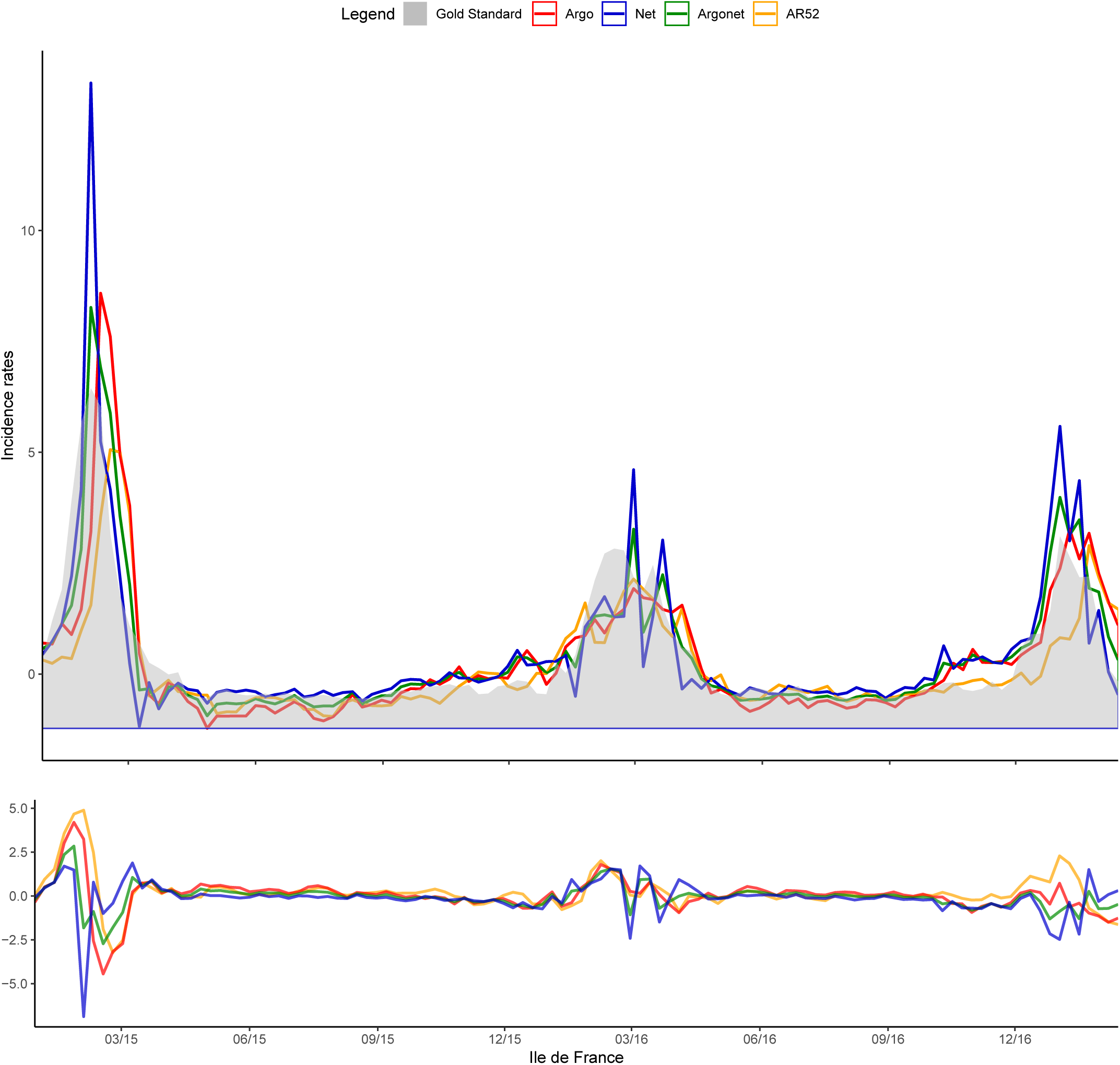
Ile de France Two-week estimate.

**Fig S55.**
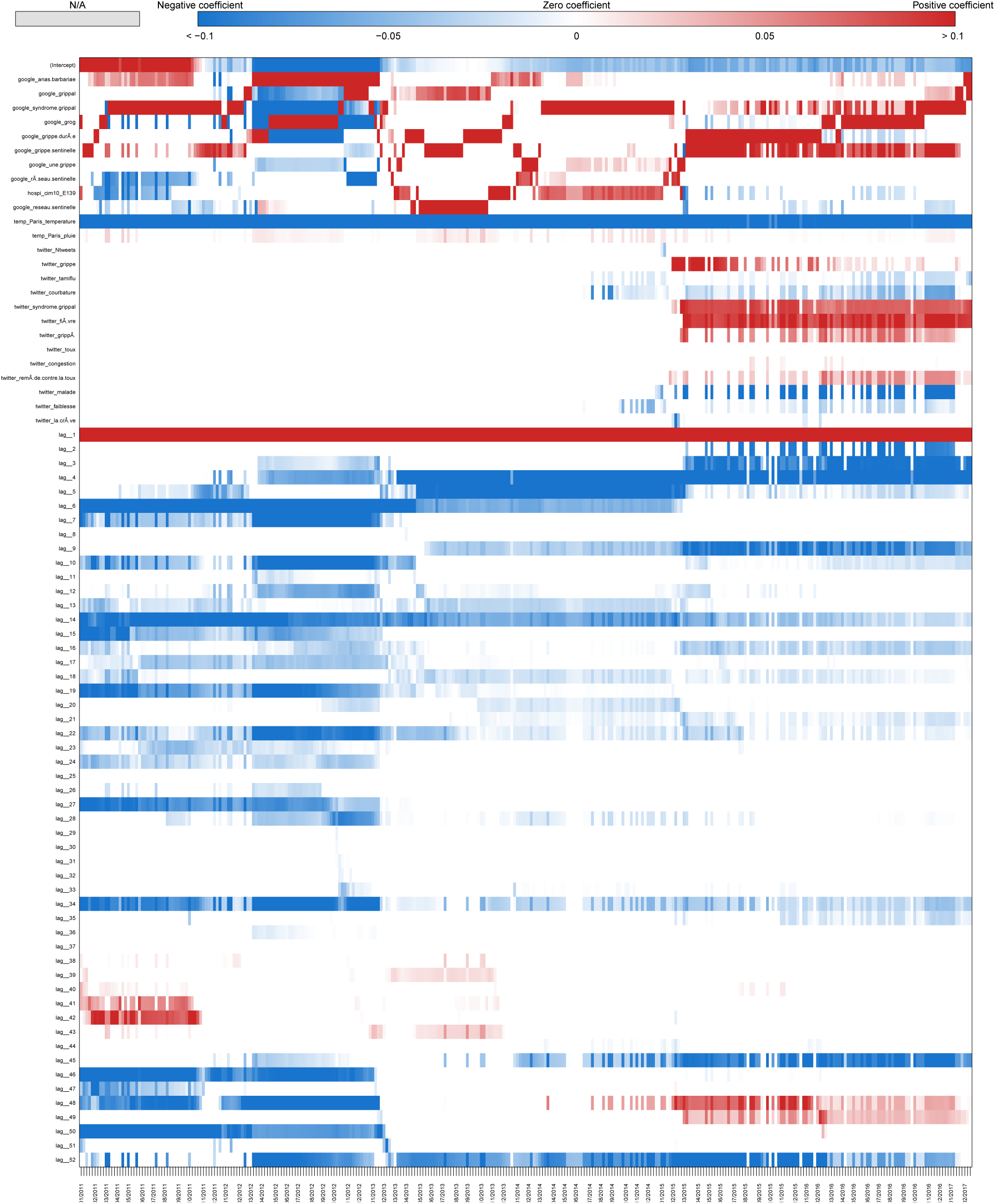
Coefficients Ile de France Two-week estimate.

**Fig S56.**
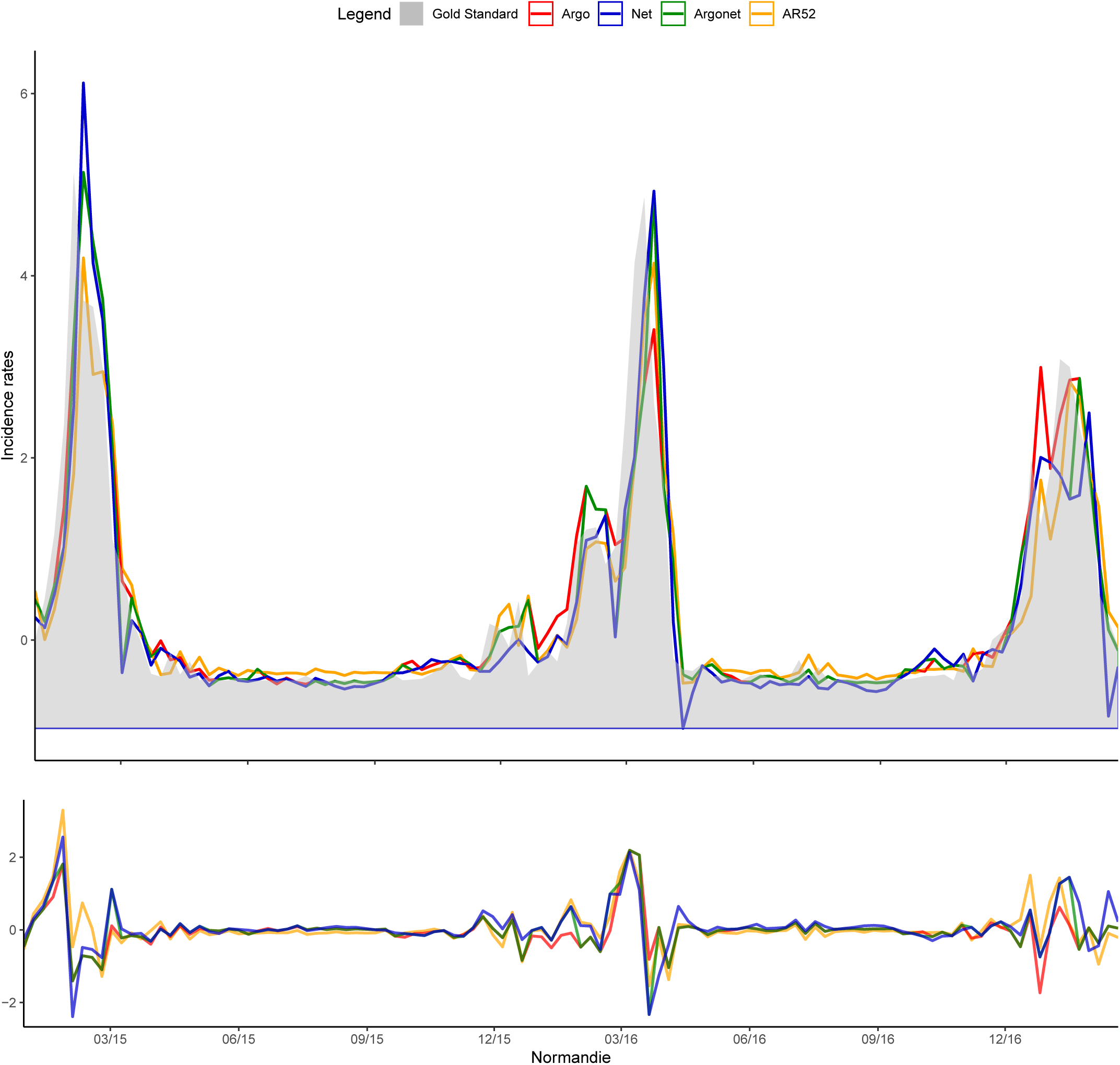
Normandie Real-time estimate.

**Fig S57.**
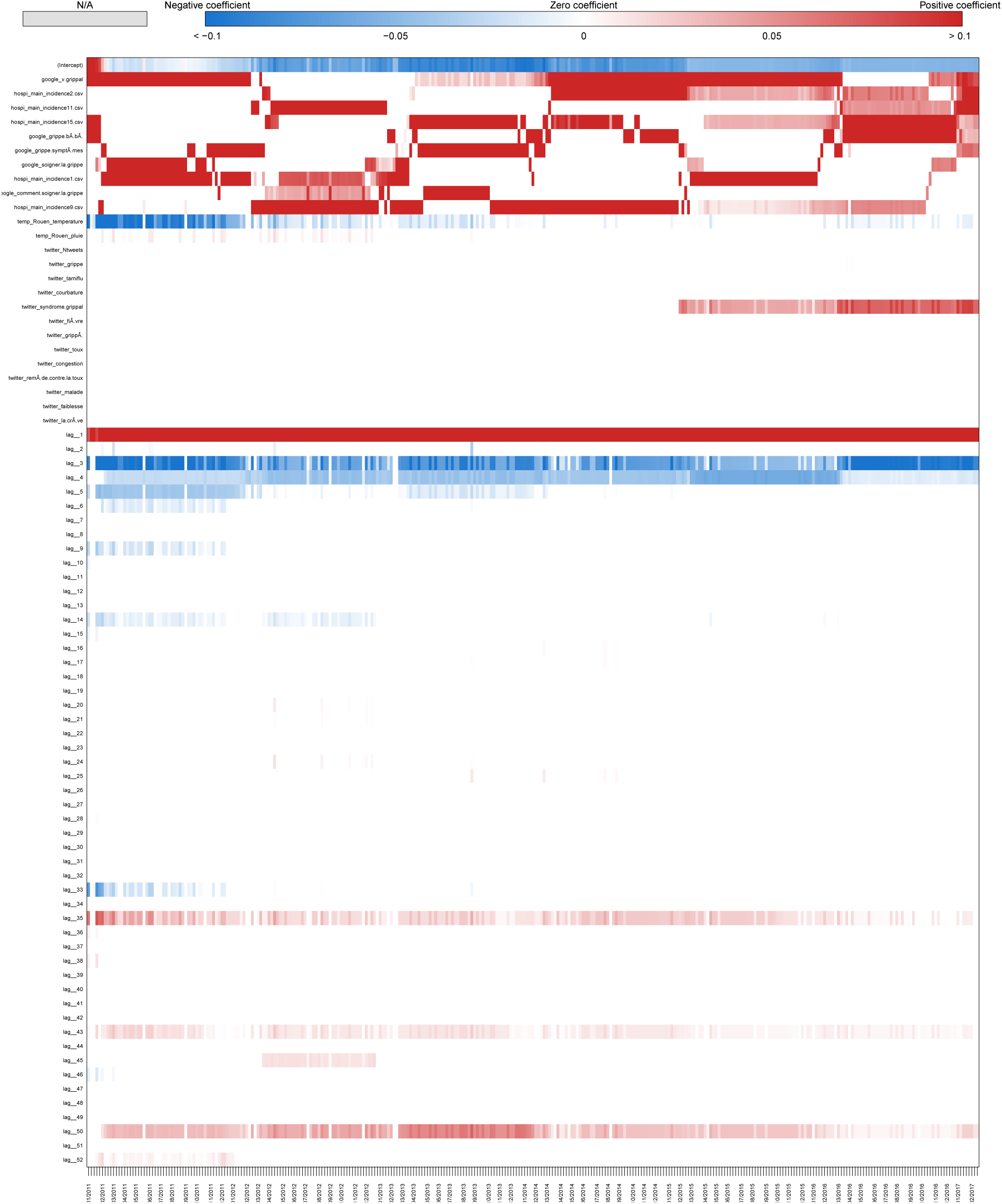
Coefficients Normandie Real-time estimate.

**Fig S58.**
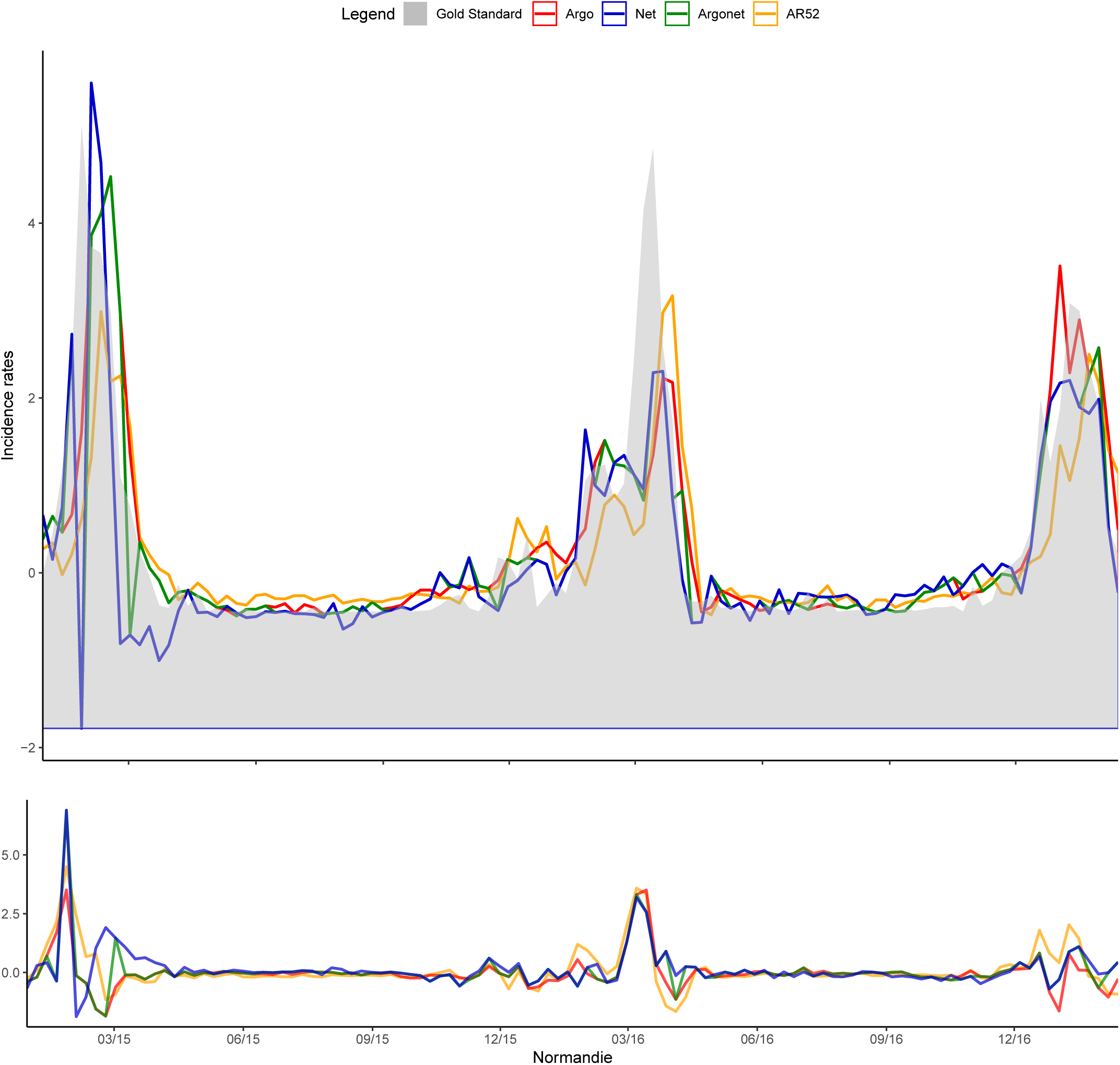
Normandie One-week estimate.

**Fig S59.**
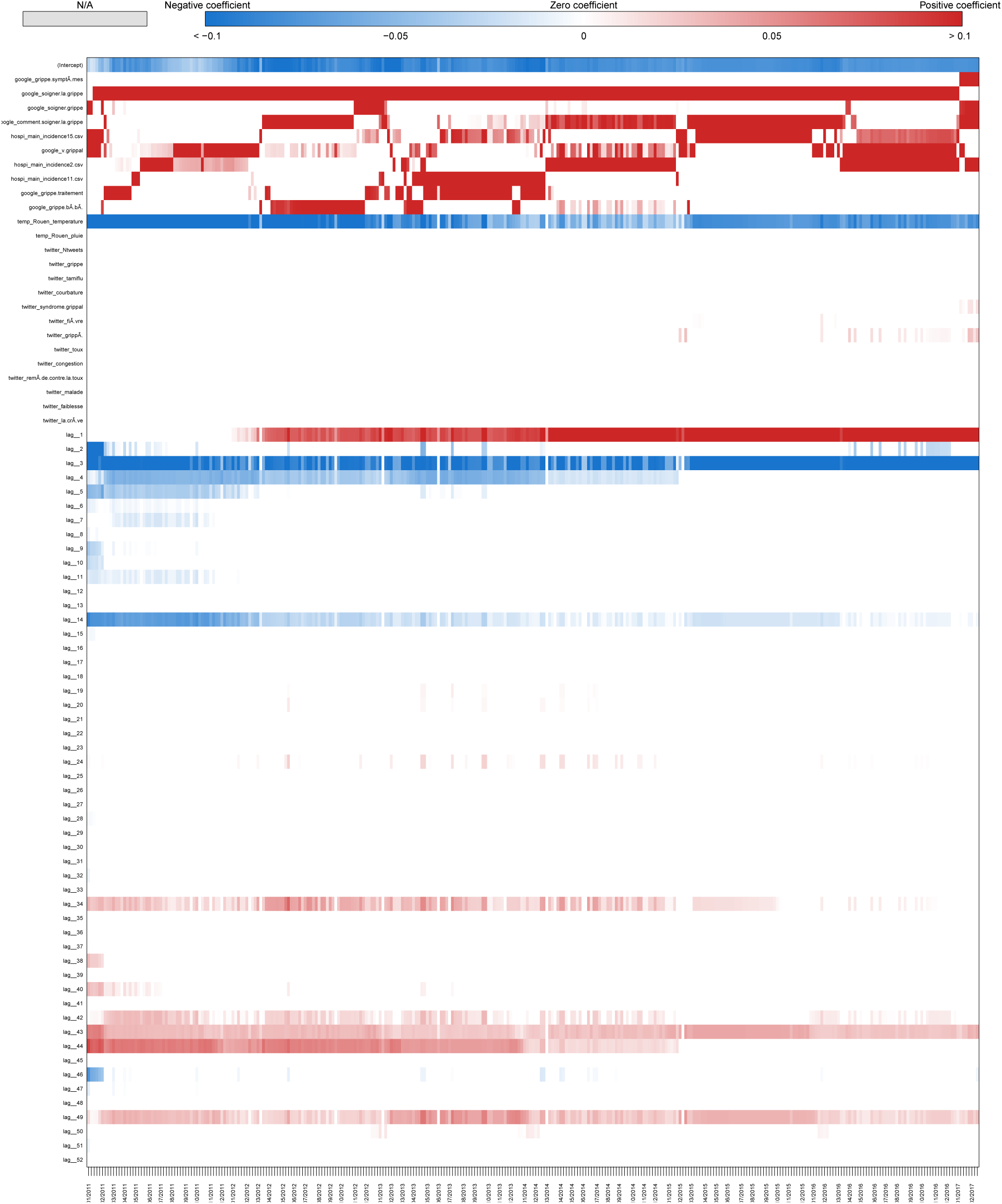
Coefficients Normandie One-week estimate.

**Fig S60.**
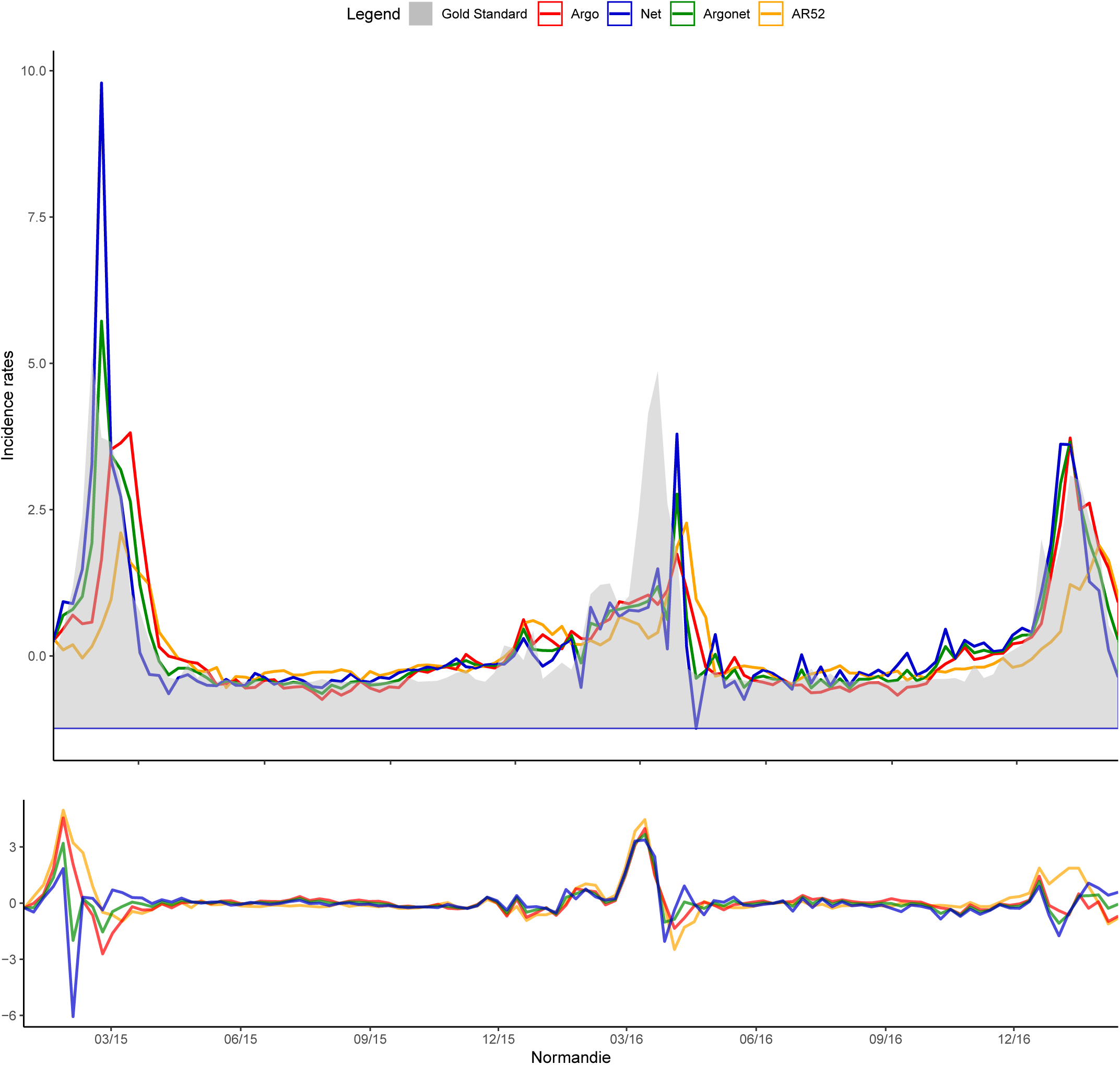
Normandie Two-week estimate.

**Fig S61.**
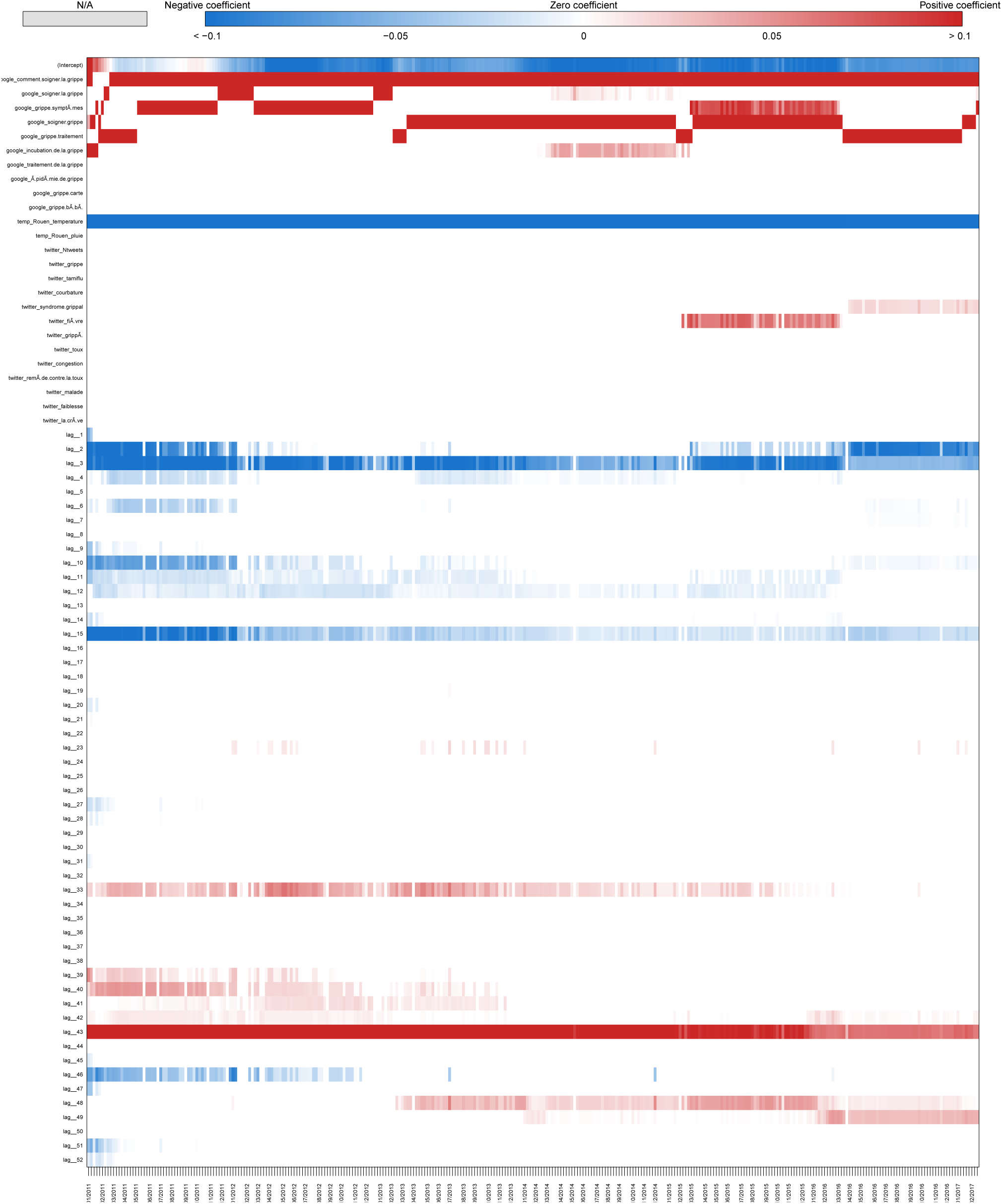
Coefficients Normandie Two-week estimate.

**Fig S62.**
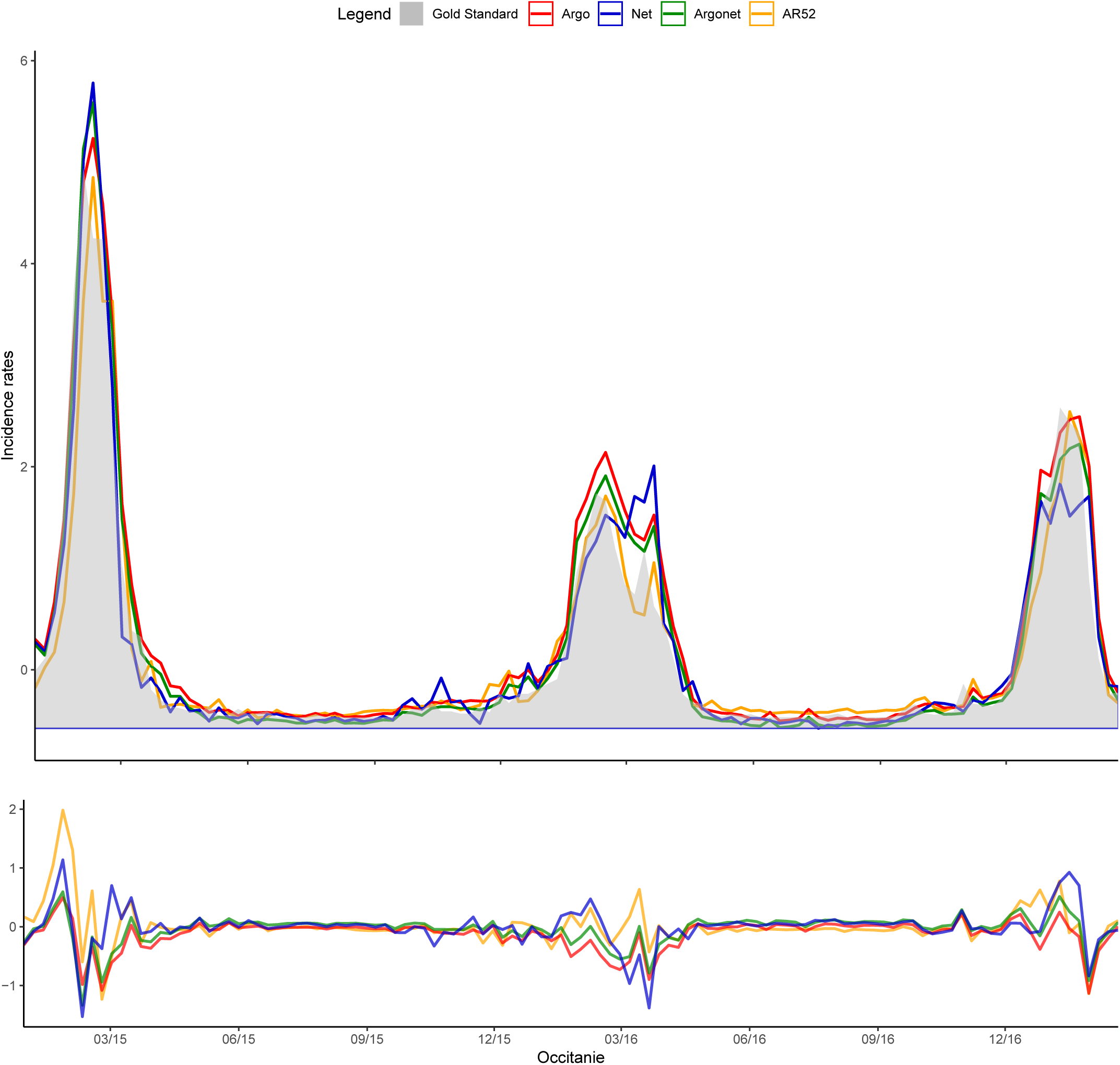
Occitanie Real-time estimate.

**Fig S63.**
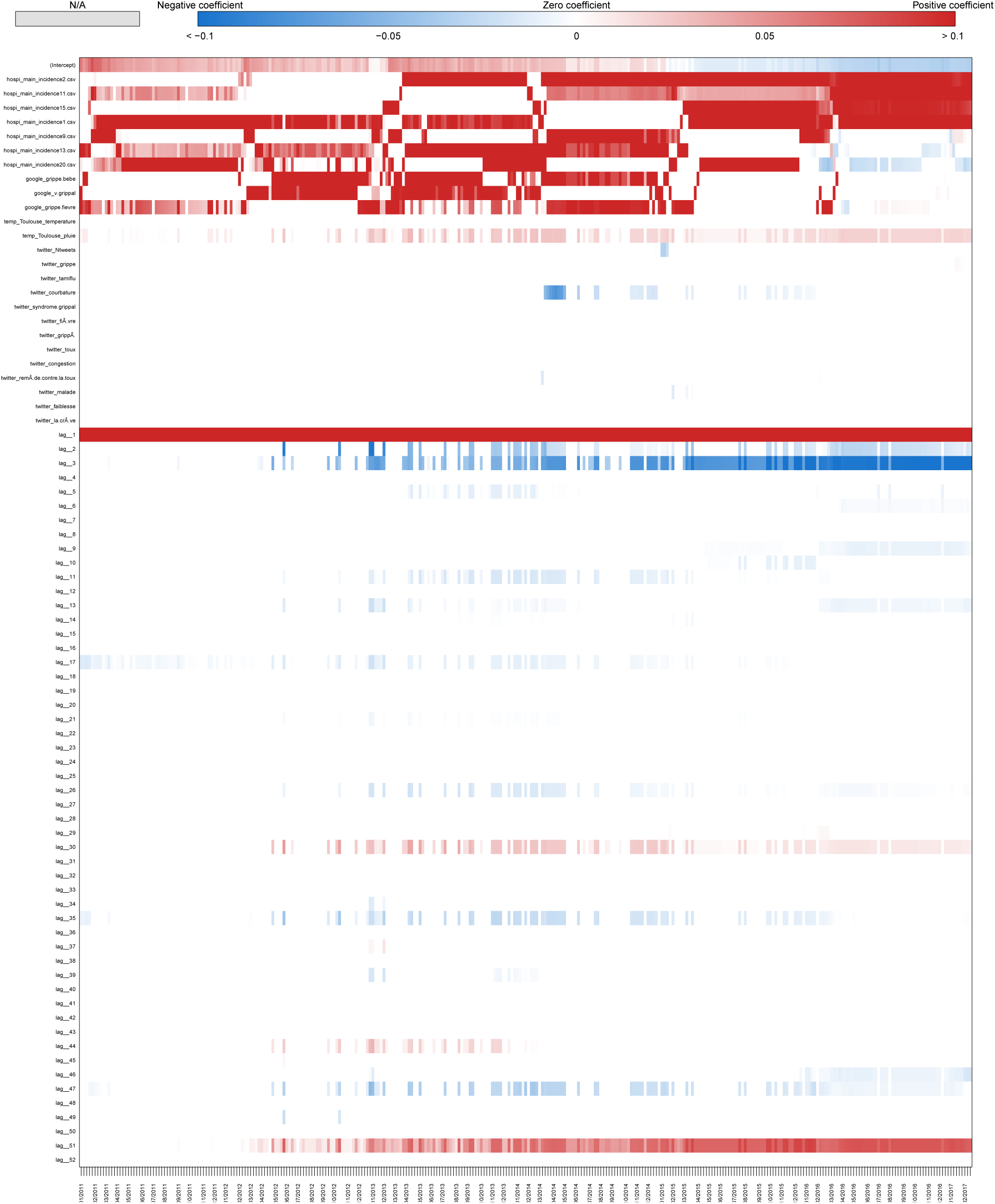
Coefficients Occitanie Real-time estimate.

**Fig S64.**
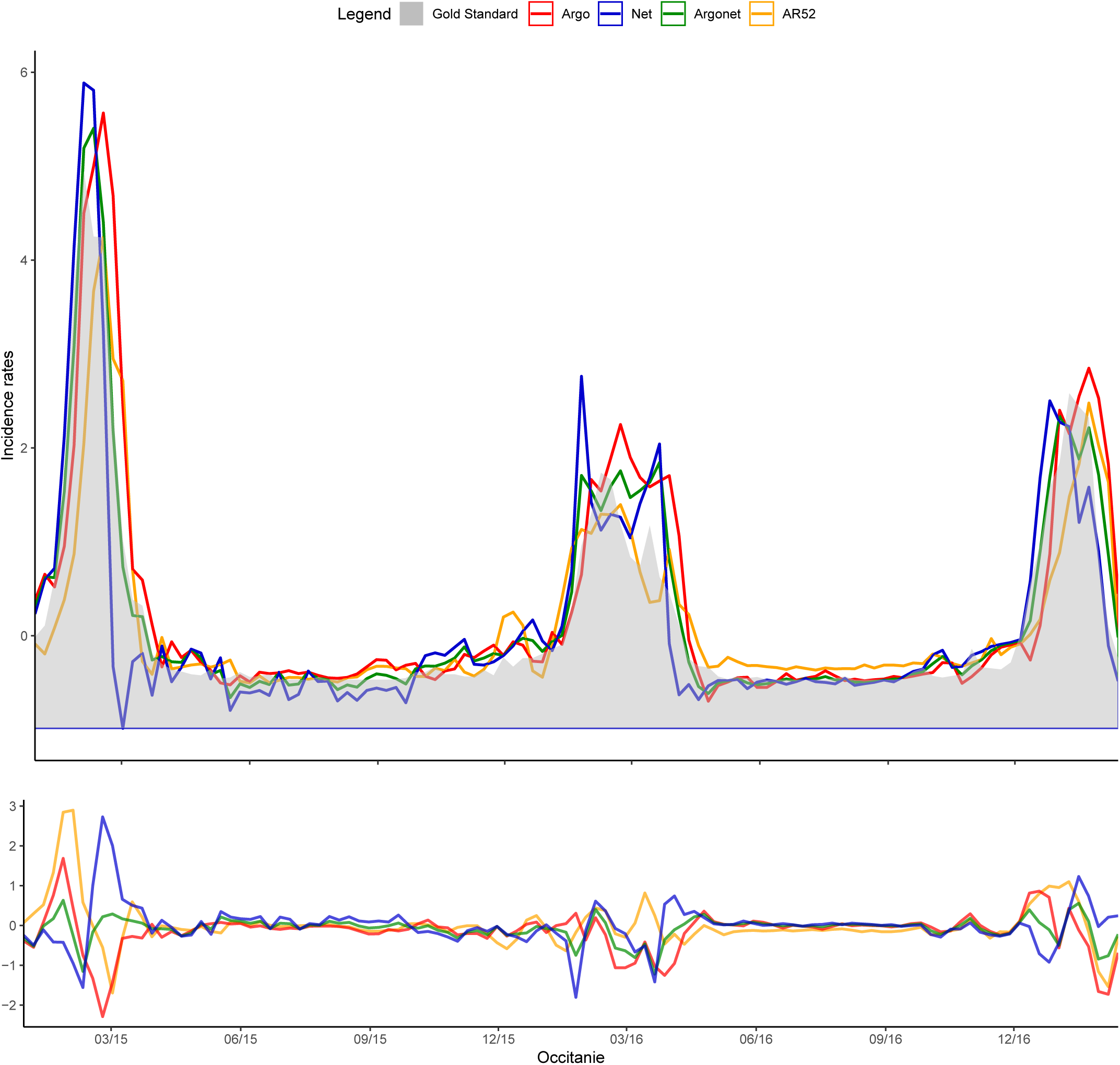
Occitanie One-week estimate.

**Fig S65.**
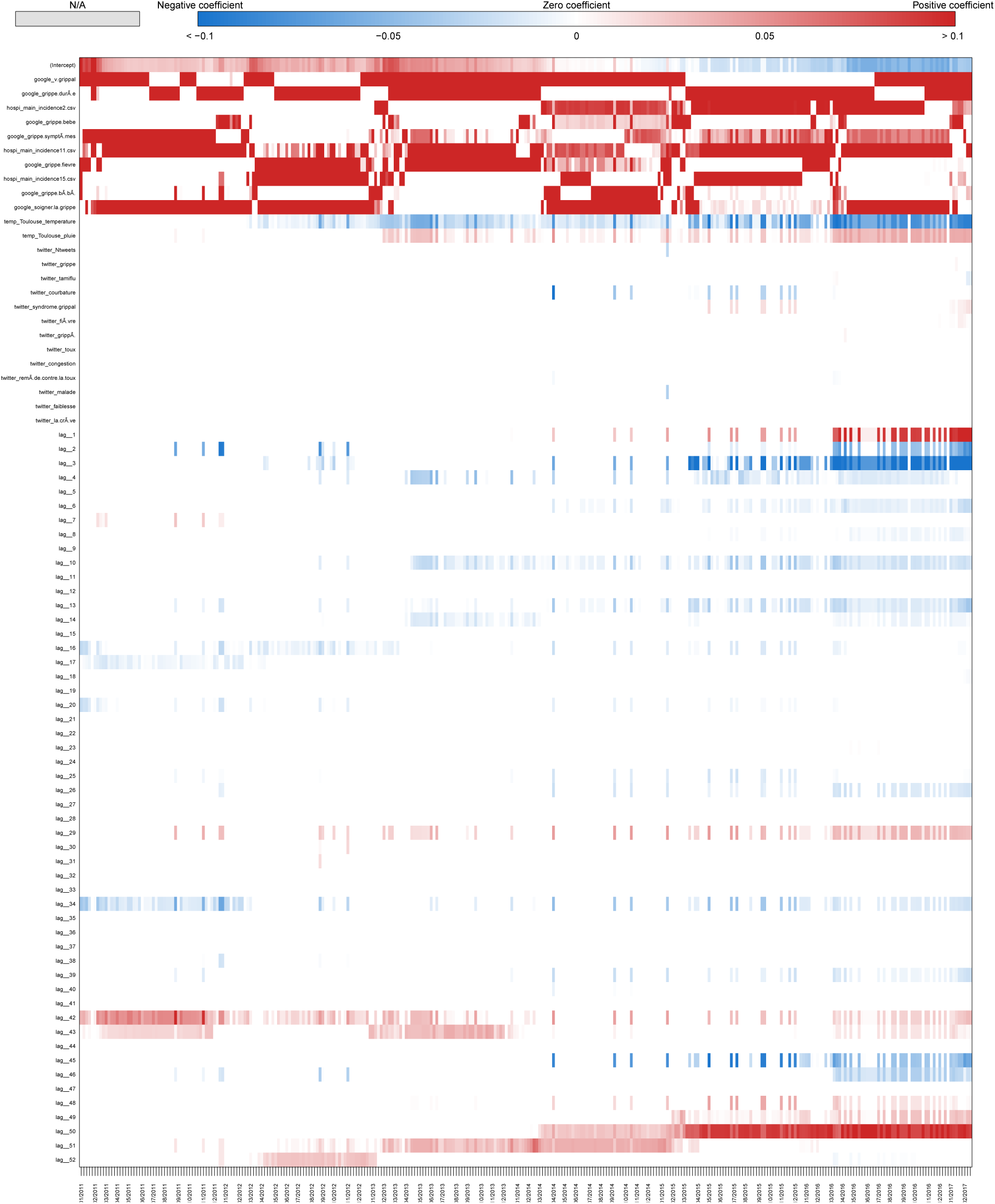
Coefficients Occitanie One-week estimate.

**Fig S66.**
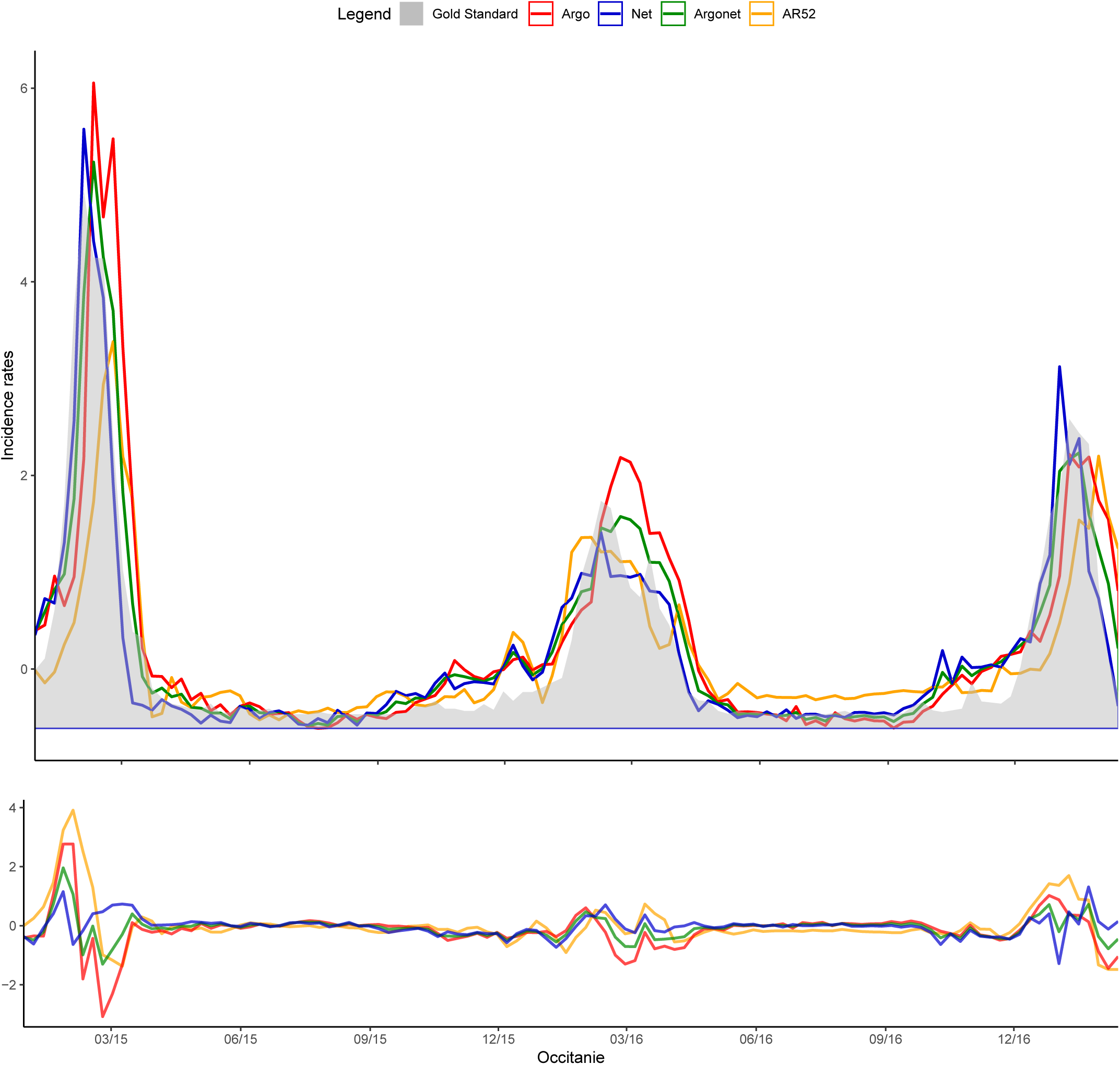
Occitanie Two-week estimate.

**Fig S67.**
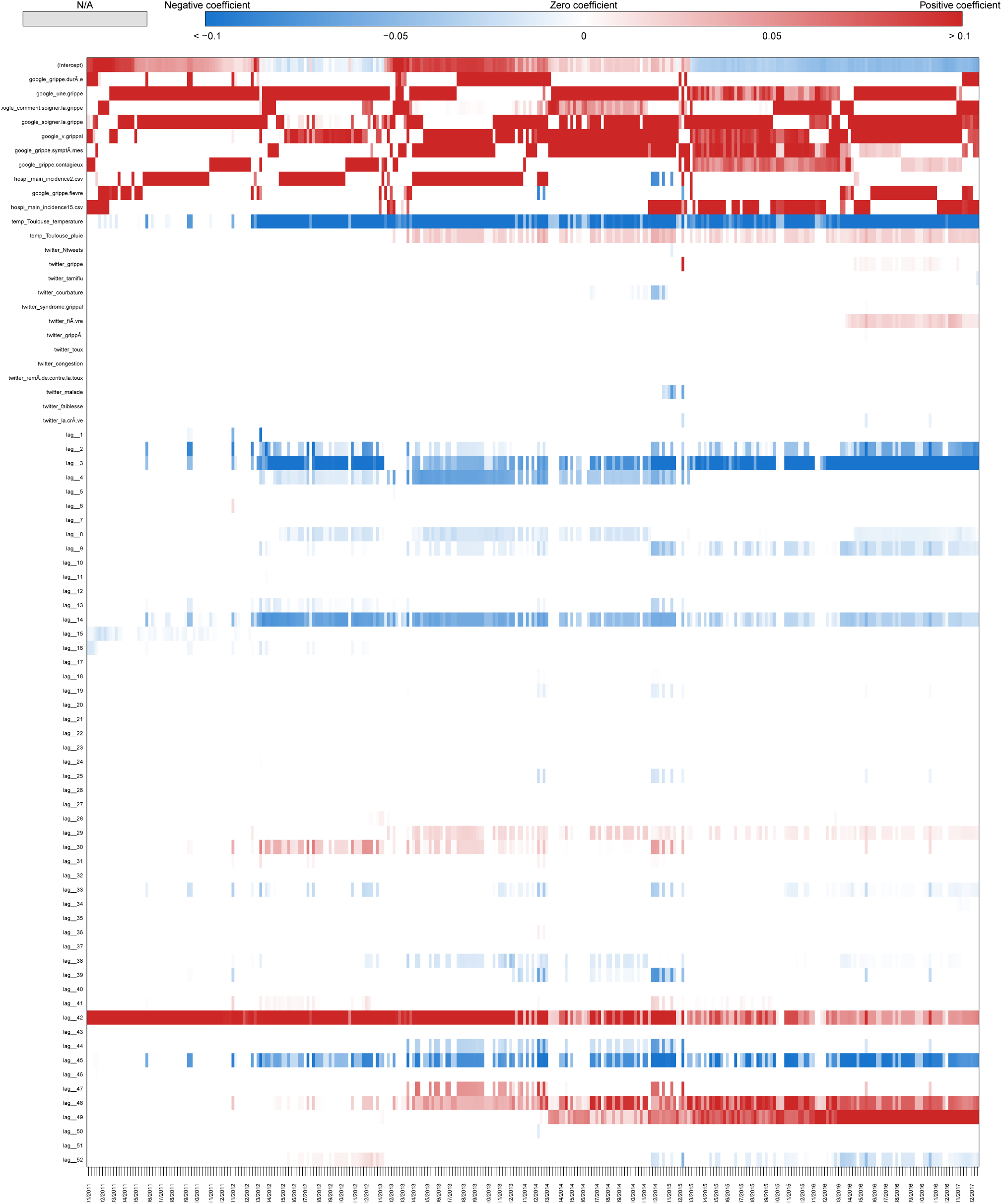
Coefficients Occitanie Two-week estimate.

**Fig S68.**
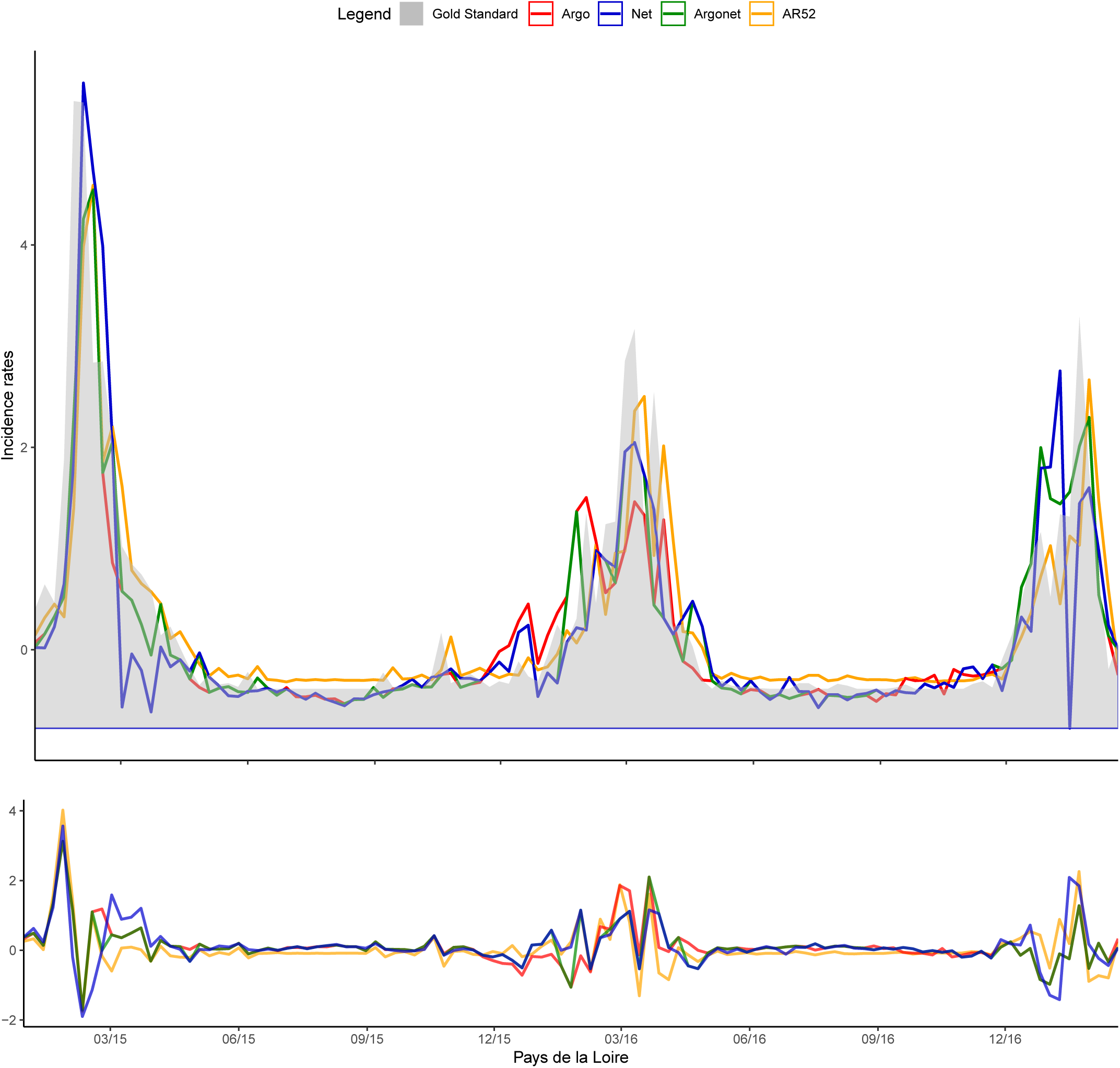
Pays de la Loire Real-time estimate.

**Fig S69.**
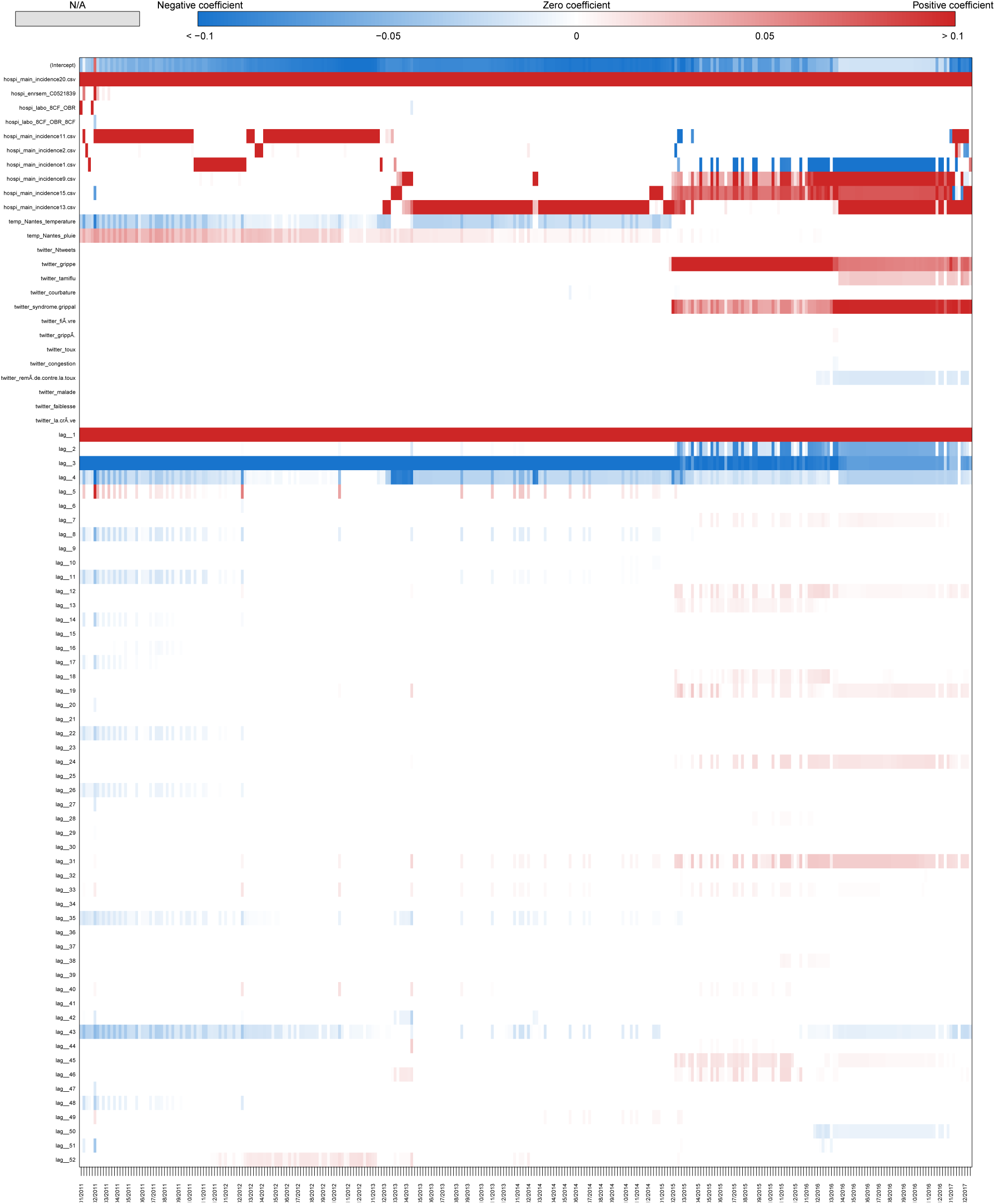
Coefficients Pays de la Loire Real-time estimate.

**Fig S70.**
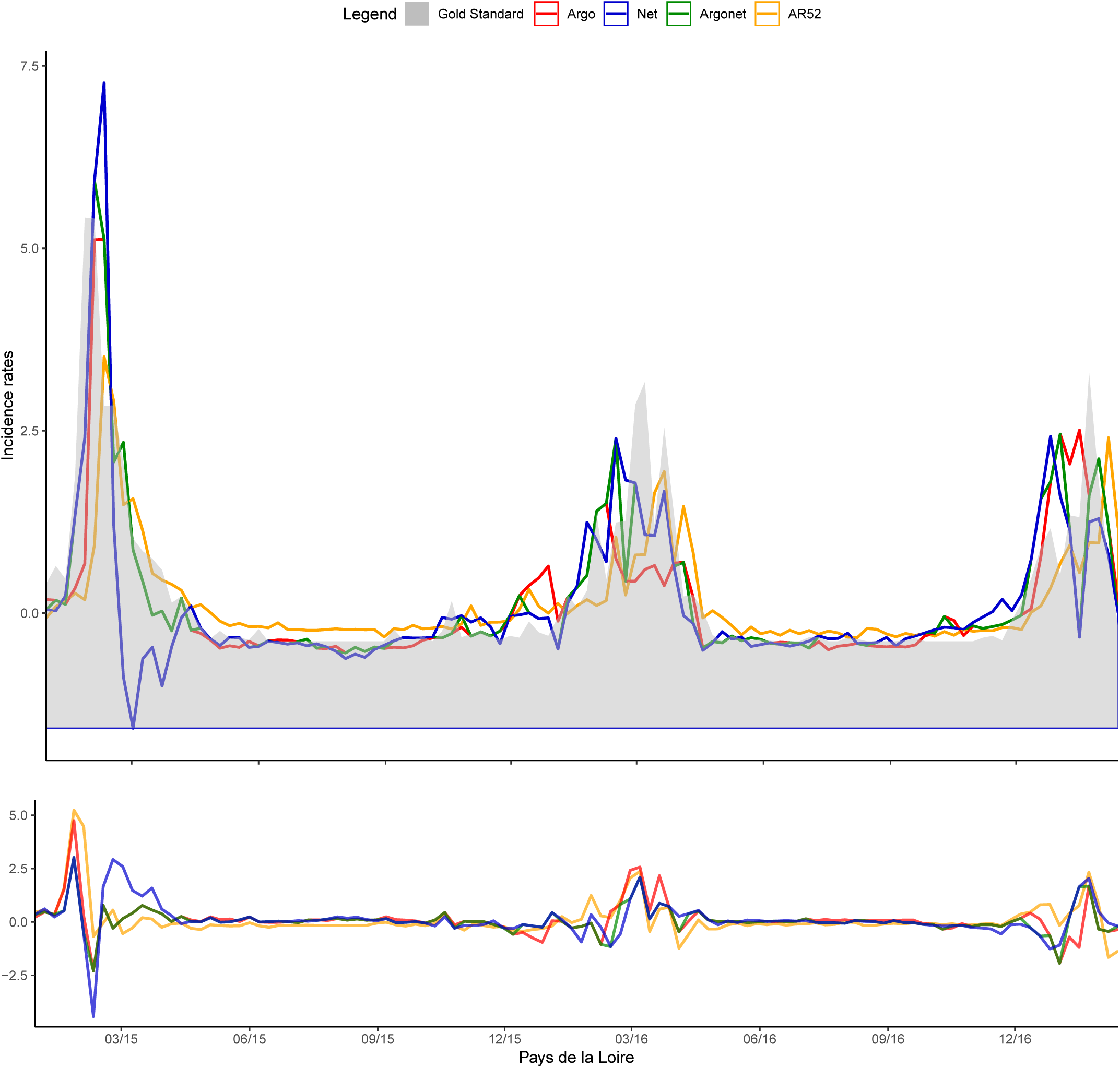
Pays de la Loire One-week estimate.

**Fig S71.**
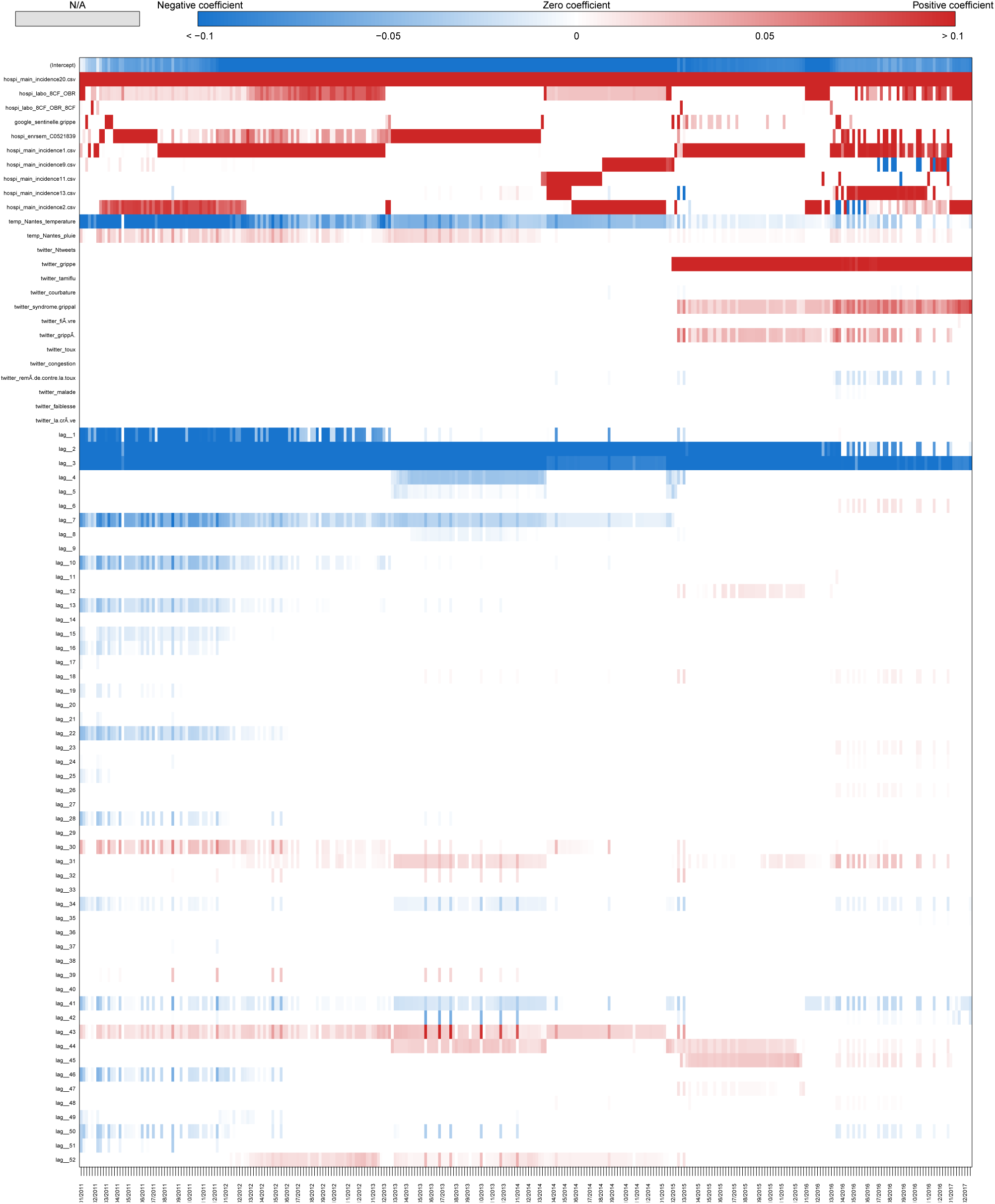
Coefficients Pays de la Loire One-week estimate.

**Fig S72.**
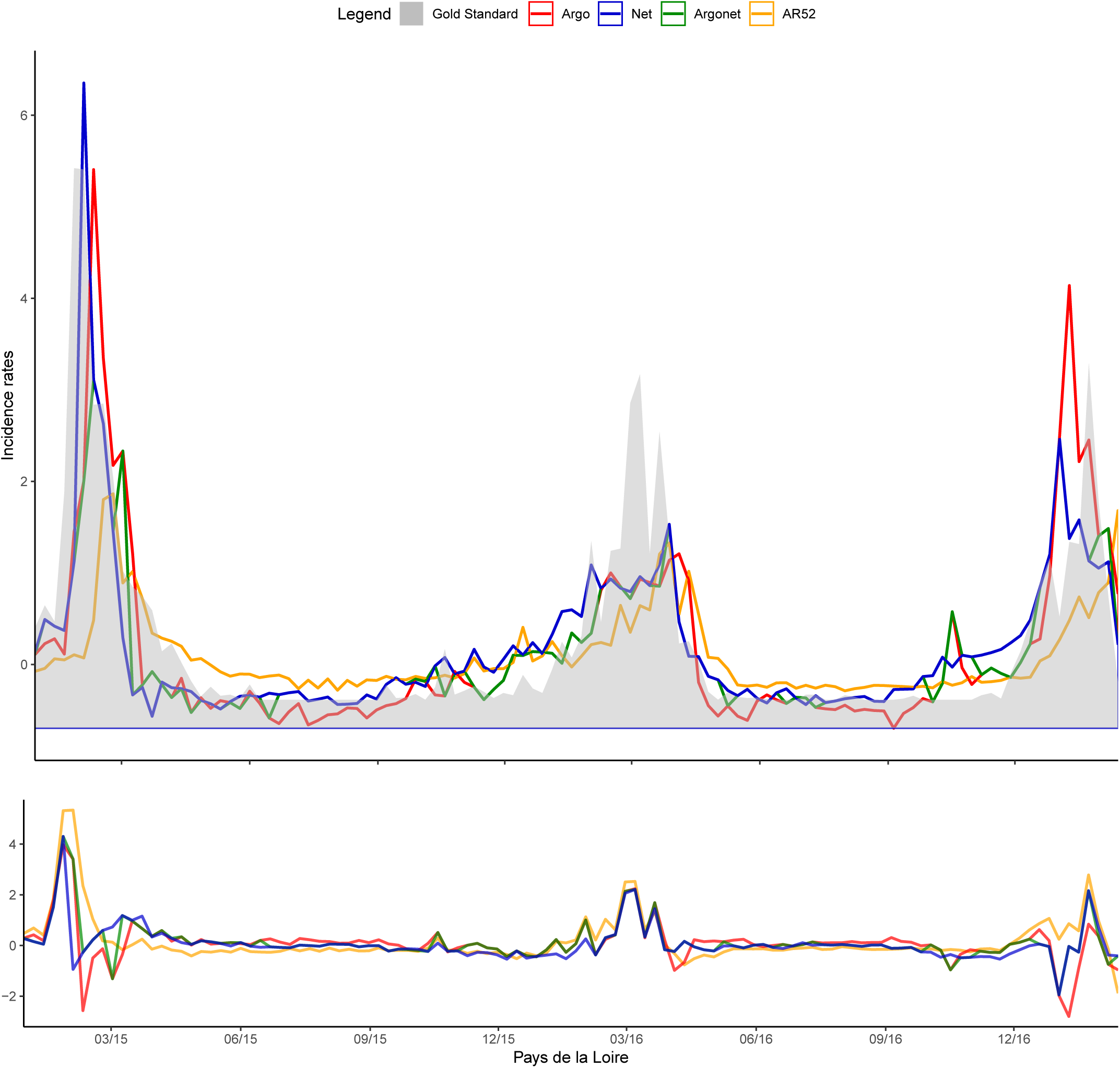
Pays de la Loire Two-week estimate.

**Fig S73.**
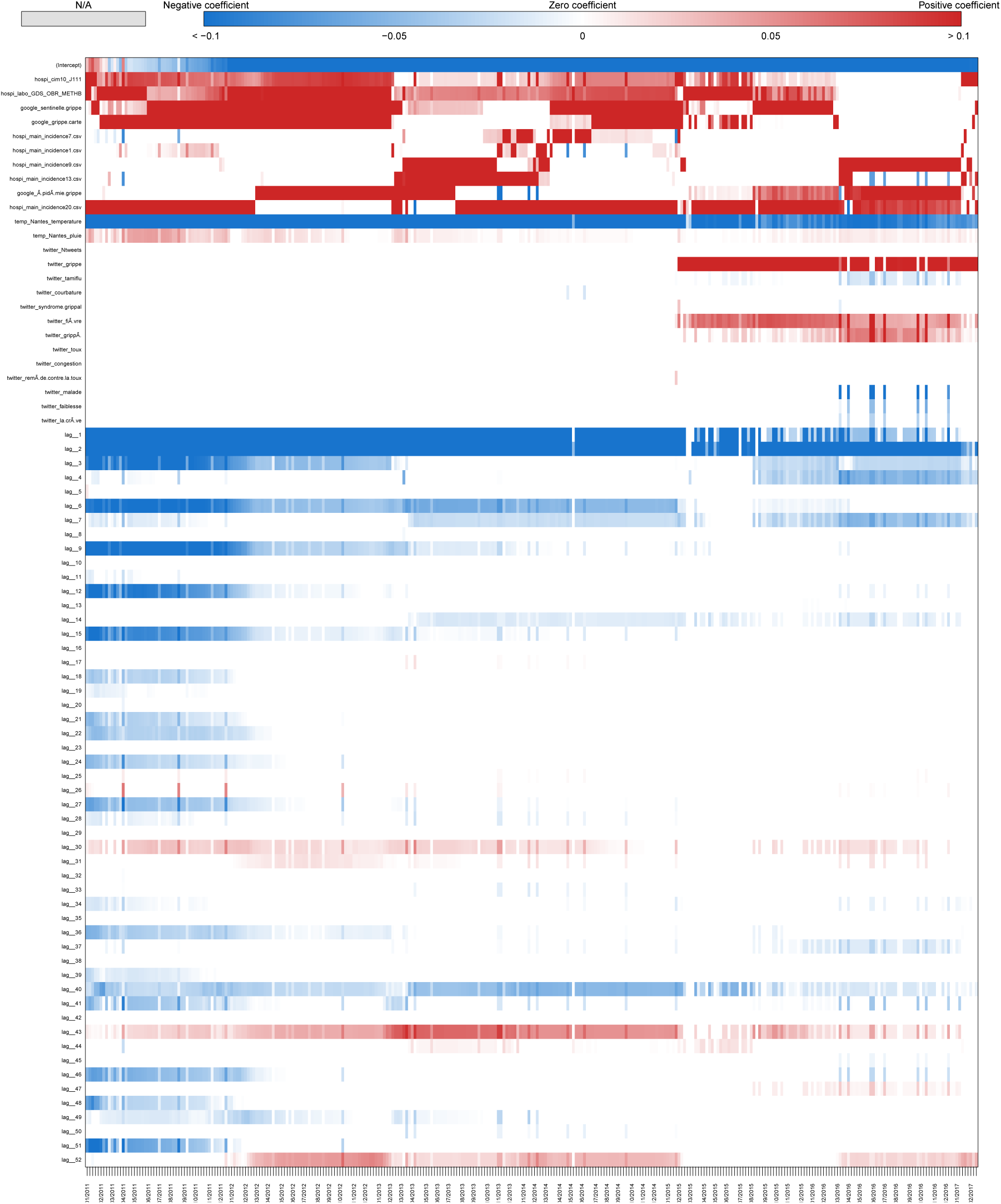
Coefficients Pays de la Loire Two-week estimate.

**Fig S74.**
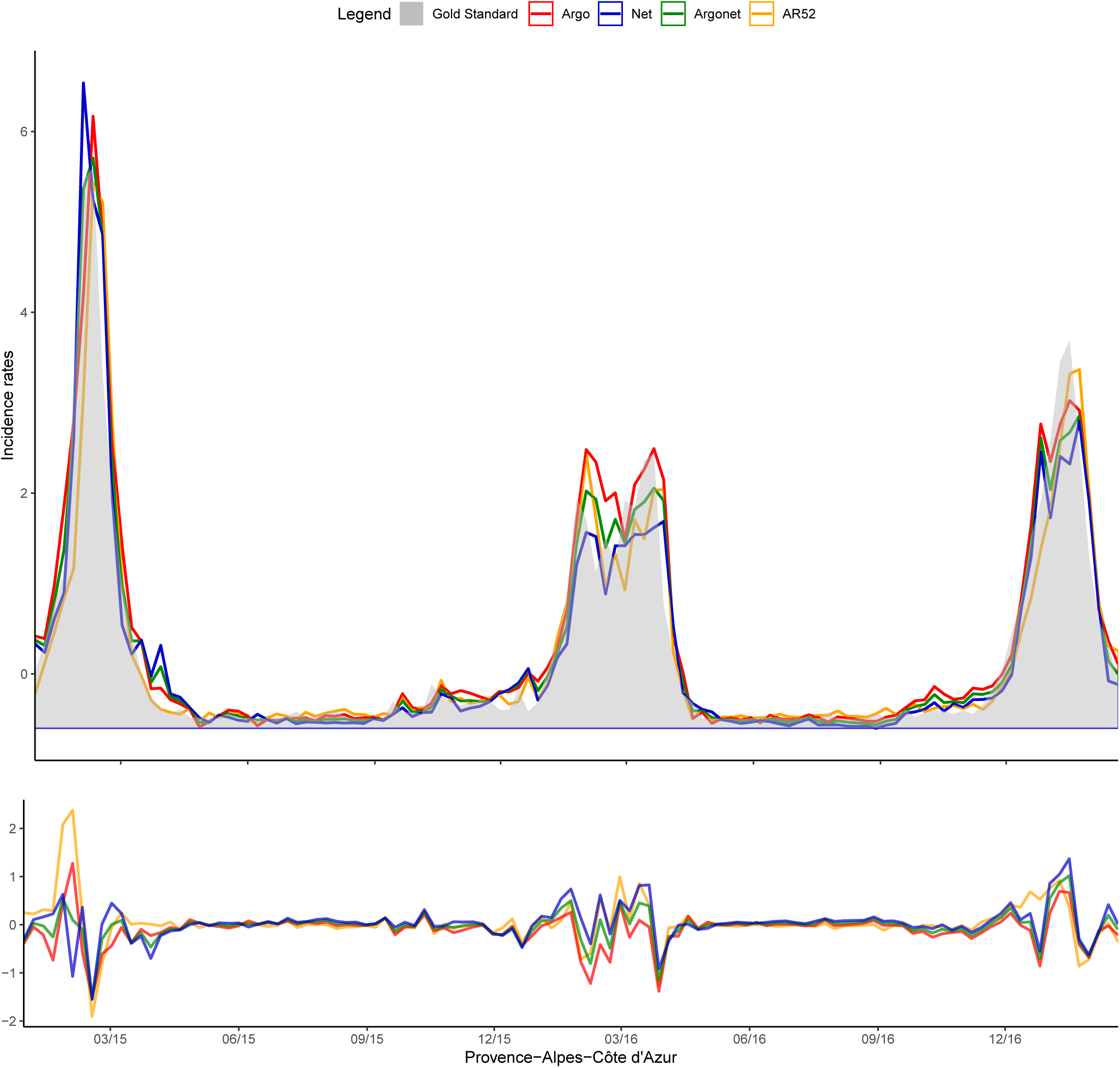
Provence Alpes Côte d’Azur Real-time estimate.

**Fig S75.**
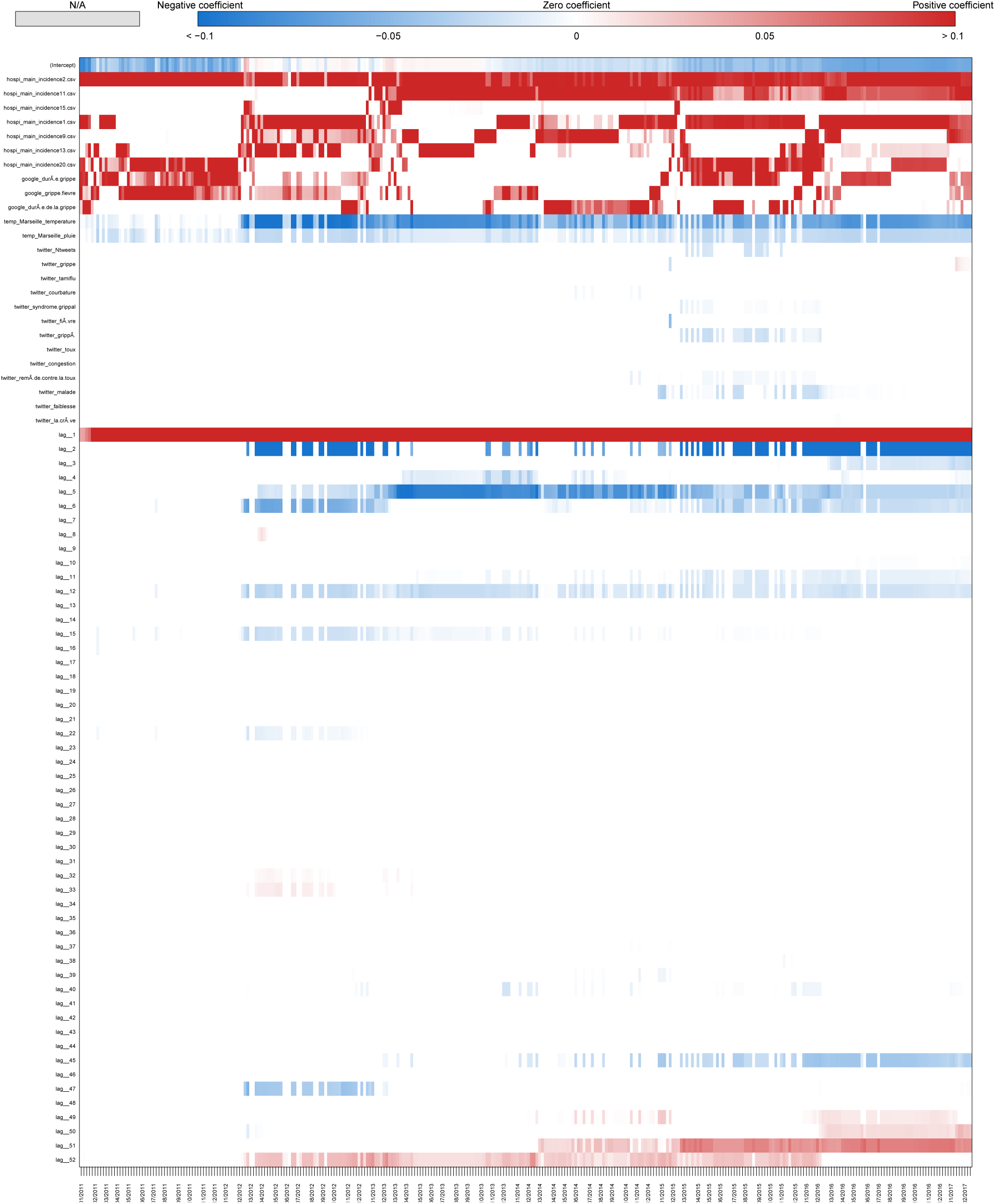
Coefficients Provence Alpes Côte d’Azur Real-time estimate.

**Fig S76.**
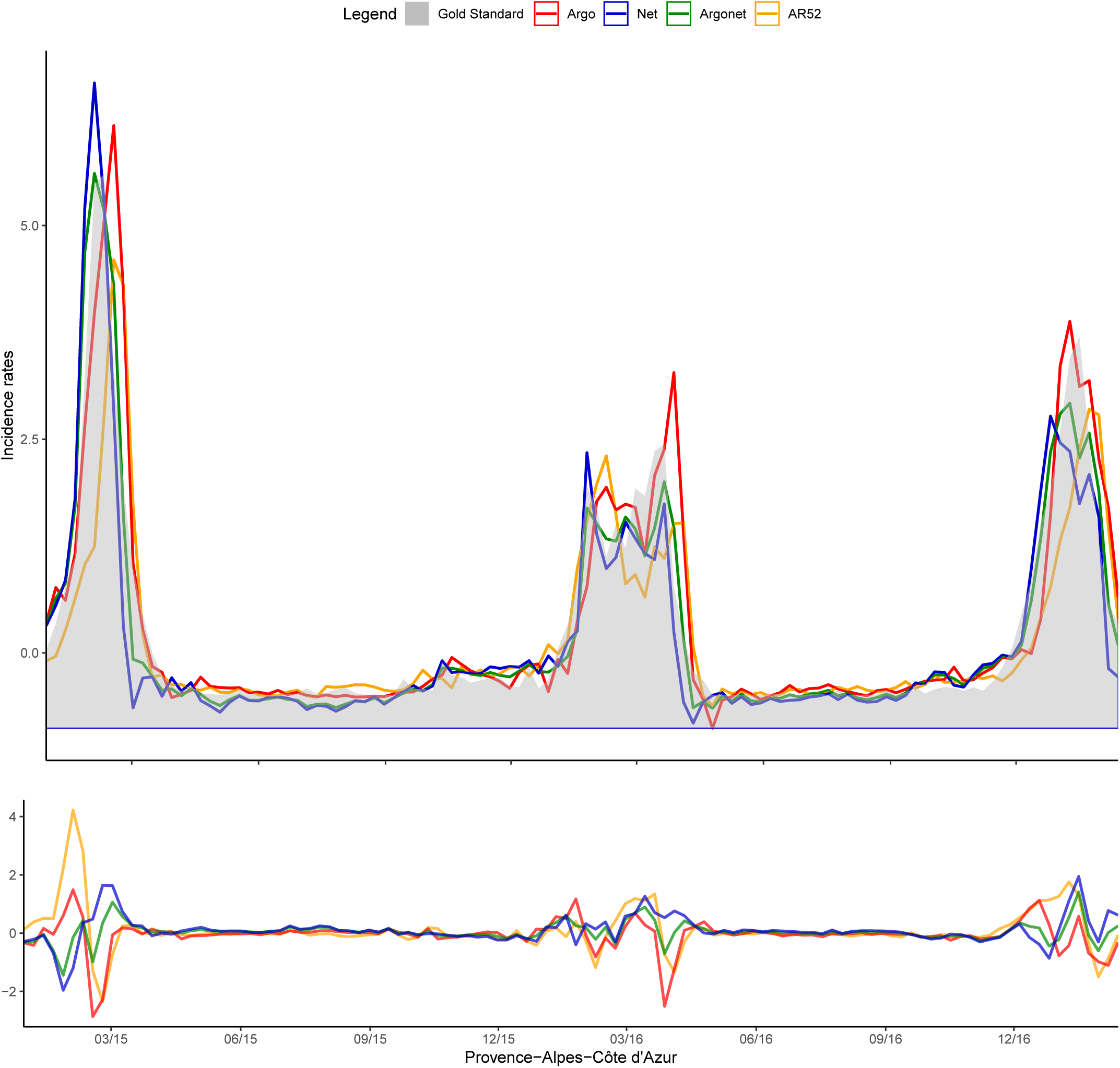
Provence Alpes Côte d’Azur One-week estimate.

**Fig S77.**
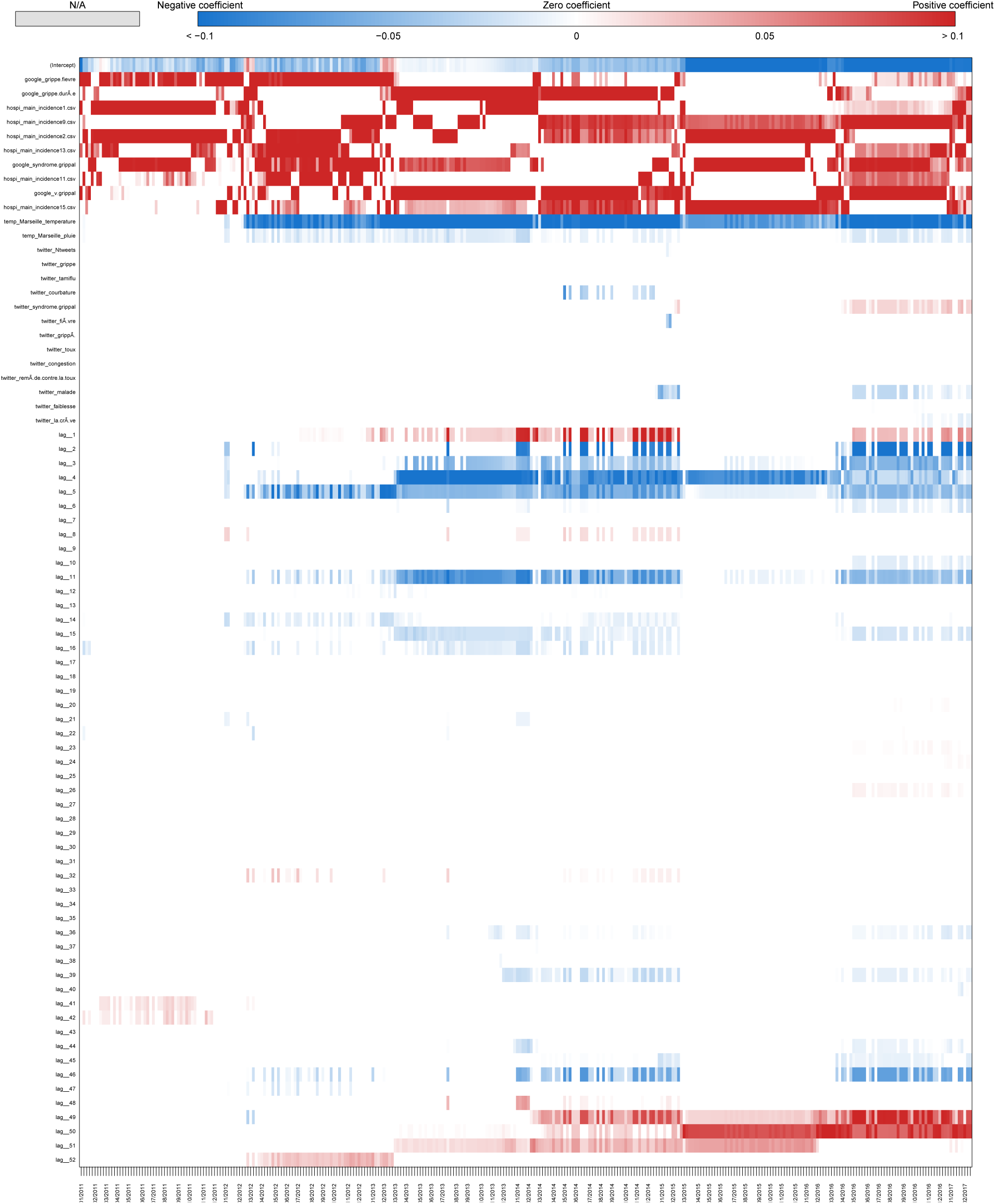
Coefficients Provence Alpes Côte d’Azur One-week estimate.

**Fig S78.**
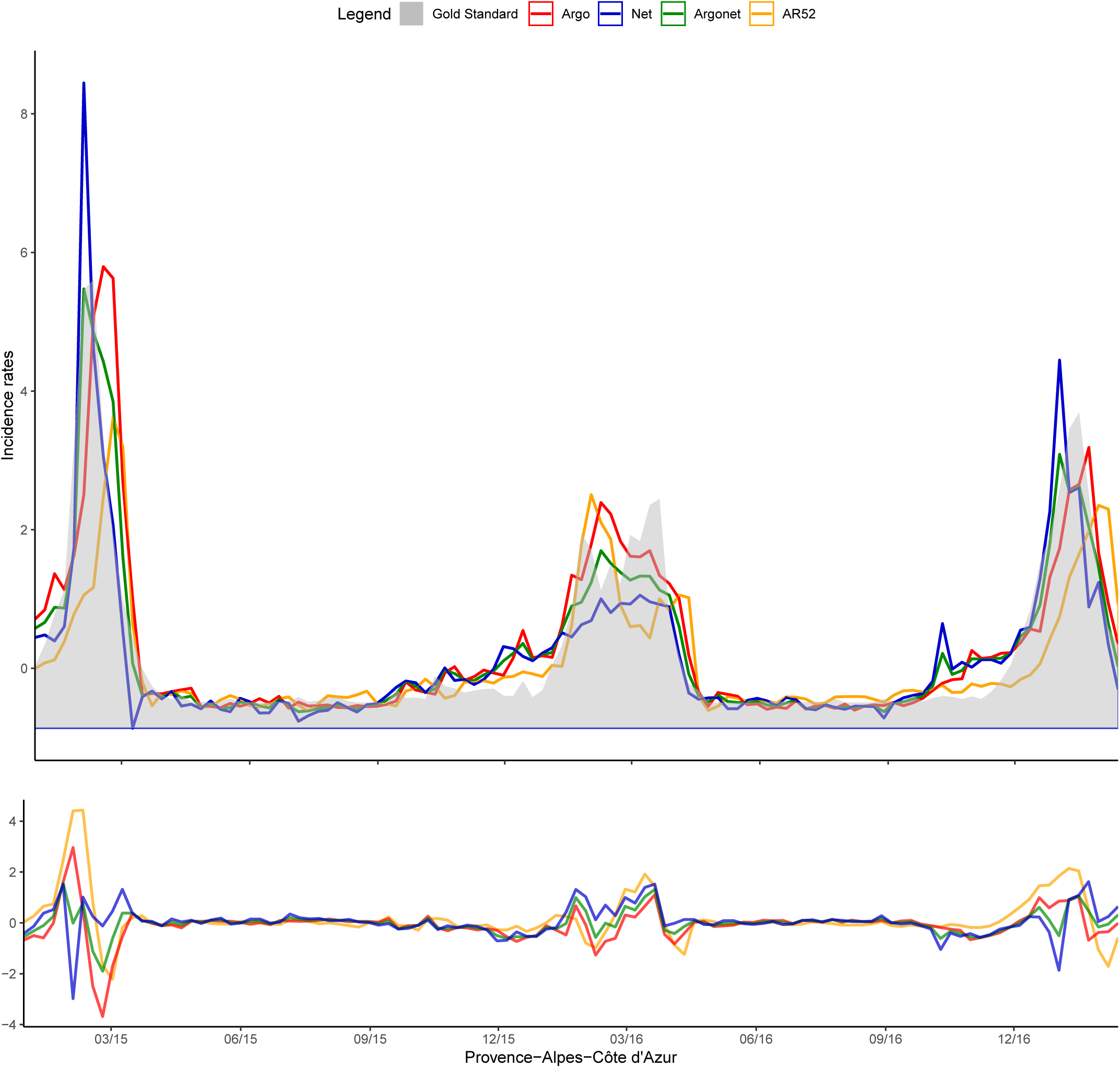
Provence Alpes Côte d’Azur Two-week estimate.

**Fig S79.**
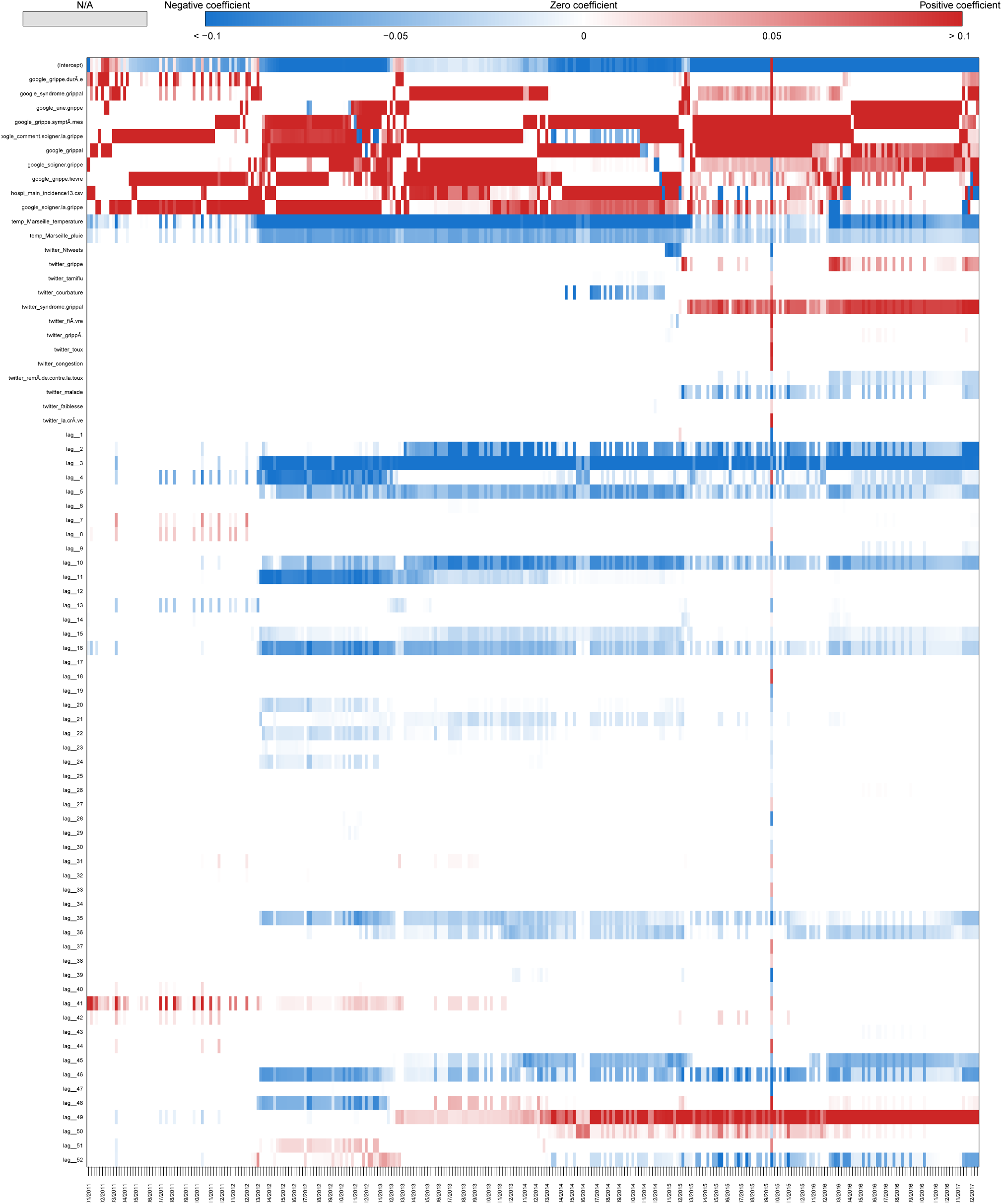
Coefficients Provence Alpes Côte d’Azur Two-week estimate.

